# Using Digital Surveillance Tools for Near Real-Time Mapping of the Risk of International Infectious Disease Spread: Ebola as a Case Study

**DOI:** 10.1101/19011940

**Authors:** Sangeeta Bhatia, Britta Lassmann, Emily Cohn, Malwina Carrion, Moritz U. G. Kraemer, Mark Herringer, John Brownstein, Larry Madoff, Anne Cori, Pierre Nouvellet

**Affiliations:** MRC Centre for Global Infectious Disease Analysis, School of Public Health, Imperial College London, Faculty of Medicine, Norfolk Place, London, W2 1PG, UK; ProMED, International Society for Infectious Diseases, Brookline, MA 02446 USA; Computational Epidemiology Group, Division of Emergency Medicine, Boston Children’s Hospital, Boston, MA USA; Department of Health Science, Sargent College, Boston University, Boston, MA USA; Department of Zoology, Tinbergen Building, Oxford University, Oxford, UK; Department of Pediatrics, Harvard Medical School, Boston, USA; healthsites.io; Evolution, Behaviour and Environment, University of Sussex, Brighton, UK

## Abstract

In our increasingly interconnected world, it is crucial to understand the risk of an outbreak originating in one country or region and spreading to the rest of the world. Digital disease surveillance tools such as ProMED and HealthMap have the potential to serve as important early warning systems as well as complement the field surveillance during an ongoing outbreak. Here we present a flexible statistical model that uses data produced from digital surveillance tools (ProMED and HealthMap) to forecast short term incidence trends in a spatially explicit manner. The model was applied to data collected by ProMED and HealthMap during the 2013-2016 West African Ebola epidemic. The model was able to predict each instance of international spread 1 to 4 weeks in advance. Our study highlights the potential and limitations of using publicly available digital surveillance data for assessing outbreak dynamics in real-time.

---

Increasing globalization of commerce, finance, production, and services has fostered rapid movement of people, animals, plants, and food [1]. With the transportation of people and goods comes the widespread dispersion of pathogens that cause infectious diseases and the vectors that may spread them [2]. Outbreaks that begin in the most remote parts of the world can now spread swiftly to urban centers and to countries far away with dangerous, global consequences [3]. For instance, population mobility across borders played a critical role in the spread of Ebola virus in West Africa during the 2013-2016 Ebola epidemic [4]. A more recent example is the 2016 yellow Fever outbreak in Angola [5]. Infected travellers from Angola reached China, representing the first ever reported cases of yellow fever in Asia [6].

Early detection and monitoring of infectious disease outbreaks through passive or active collection of surveillance data can help public health officials initiate interventions such as removing contaminated food sources, isolating affected individuals or launching vaccination campaigns. However, any data collection method involves trade-offs between speed, accuracy and costs. Data collected through traditional surveillance, for example via public health infrastructure, are generally reliable but are resource intensive and are therefore typically available for upstream analysis with an (understandable) delay [7].

In the past three decades, the internet has grown at a staggering pace, with approximately half of the world’s population accessing internet in 2017 [8]. The rapid growth of the internet has fostered a corresponding increase in tools for internet based disease detection and monitoring that lie at the other end of the spectrum. Digital disease surveillance consists of monitoring online information sources to collate relevant information about diseases. The sources of information can be formal such as advisories posted by a ministry of health, or informal such as news media items, blogs or tweets. Digital surveillance makes data collection less expensive and time consuming but the acquired data often contain more noise than those collected through traditional public health surveillance. While traditional surveillance systems report on select pathogens and depend on a well-functioning public health infrastructure, digital surveillance in contrast typically monitor a wide range of pathogens using little to no additional infrastructure. Thus, digital surveillance tools can play a significant role in the rapid recognition of public health emergencies [9, 10].

The Program for Monitoring Emerging Diseases (ProMED, www.promedmail.org) was one of the first entrants in the field of digital disease surveillance. ProMED was created in 1994 as a surveillance network to provide early warning of emerging and re-emerging infections [11]. ProMED collates information from various sources that include media reports, official reports, local observers, and a network of clinicians throughout the world. The reports generated through bottom-up surveillance are reviewed by a team of subject matter experts before being posted to the ProMED network. ProMED now provides free email based reports on outbreaks to over 70,000 subscribers in at least 185 countries.

HealthMap (www.healthmap.org) is another widely used tool for disease outbreak monitoring. In addition to ProMED alerts, HealthMap utilises online news aggregators, eyewitness reports and other formal and informal sources of information and allows for visualisation of alerts on a map [12]. The surveillance data collected by HealthMap has been incorporated into the Epidemic Intelligence from Open Sources (EIOS) surveillance system, developed by WHO. Both ProMED and HealthMap are used by key public health bodies, including the Centers for Disease Control and Prevention (CDC) and the World Health Organisation (WHO).

Some other examples of digital disease detection tools include MediSys (http://medisys.newsbrief.eu/), H5N1 Google Earth mashup (www.nature.com/avianflu/google-earth), and Emerging Infections Network (http://ein.idsociety.org). These tools provide a unique opportunity to rapidly detect new outbreaks and follow their evolution in near real-time.

Collating timely data, while critical, is only the first step in the disease surveillance process. Compiling the data, conducting analyses, and generating reports that are easily understood and actionable are equally important. For instance, one of the ProMED outbreak analysts reported in March 2014 on the likely spread of Zika to the Americas [13], well before the epidemic surfaced in South America in February 2015. However, lacking an easy to use and openly accessible tool to quantify and visualize the reported risk of disease spread, this report did not have any significant impact on public health resource allocation and decision making. While there has been a growing interest in using various internet data streams for epidemiological investigations [14, 15] and in using data from digital surveillance tools [16], there is as yet a dearth of a framework that can automatically combine such data with other streams of information, analyse them in a statistically robust manner, and produce actionable reports, particularly in real time during an outbreak where such analyses would be most useful.

In this study, we propose a new statistical framework to estimate and visualize risks posed by outbreak events using digital surveillance data. Our approach relies on a relatively simple statistical framework that integrates multiple data streams to predict the future incidence trajectory, and quantifies spatial heterogeneity in the risk of disease spread. In this paper, we present the model implemented in this framework and as a case study, we report the analysis of ProMED and HealthMap data for the 2013-16 West African Ebola epidemic. To assess the robustness of the digital surveillance data, we also applied the framework to the data collated by WHO and made available at the end of the epidemic. We present a comparison of the near real-time ProMED/HealthMap data with the retrospective WHO data and the results of the spatio-temporal analysis carried out on these three data sources.

Our analysis based solely on epidemic data available through ProMED/HealthMap provides a realistic appraisal of their strengths and weaknesses, especially if used in near real-time forecasting.

## Results

Incidence time series were computed from ProMED, HealthMap and WHO data (see Methods) for the three mainly affected countries, Guinea, Liberia and Sierra Leone, and are shown in Fig 1. We pre-processed the data from ProMED and HealthMap to remove inconsistencies and to fill gaps (see Methods for details). The raw and processed data from ProMED and HealthMap for all countries included in the feed are available in the Github repository for this article. There were substantial differences between the incidence time series derived from the three data sources, particularly at the peak of the epidemic. There may be multiple reasons underpinning such discrepancy, including potential variability in digital surveillance reporting during the course of the epidemic. It is also worth highlighting that the WHO data used here are an extensively cleaned version of the data collected during the epidemic [17,18], published more than a year after the epidemic was declared to be over. Moreover, while ProMED and HealthMap did not record probable cases for this epidemic, the WHO data contained confirmed, probable and suspected cases. However, despite these discrepancies, the weekly incidence derived from ProMED and HealthMap was moderately to highly correlated with that reported by WHO later (Pearson’s correlation coefficients aggregated across the three countries 0.44 and 0.74 respectively, p value *<* 0.001, also see Fig 2 for weekly trends).

**Figure 1:**
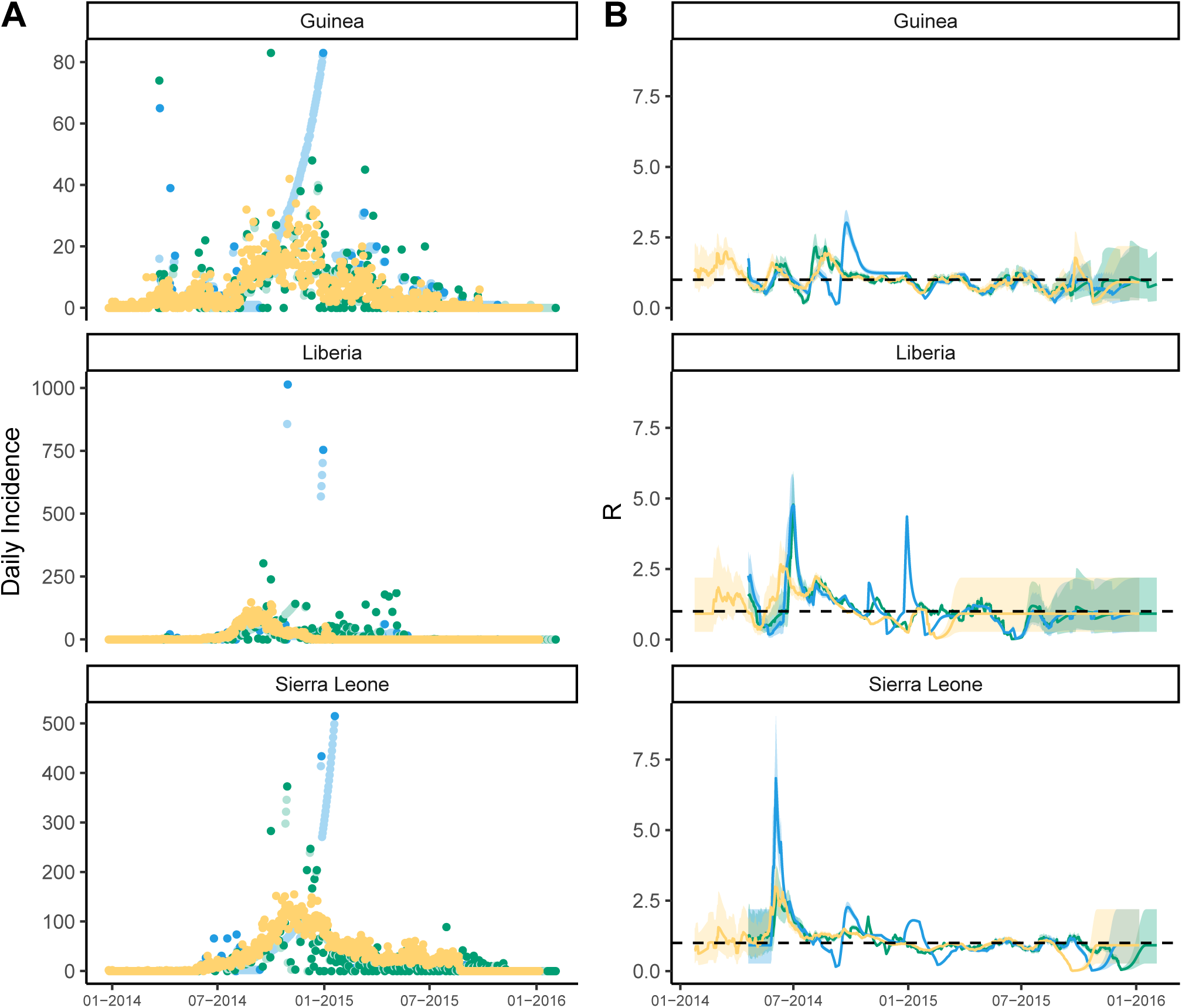
Comparison of national daily incidence trends and R estimates from ProMED, HealthMap and WHO data for Guinea, Liberia and Sierra Leone. (A) Daily incidence derived from ProMED (blue), HealthMap (green) and WHO data (orange). Daily incidence that were not directly available from ProMED and HealthMap data and which were therefore imputed (see Methods) are shown in lighter shade of blue and green respectively. WHO data were aggregated to country level. The y-axis differs for each plot. (B) The median time-varying reproduction number *R*^*t*^ estimated using the WHO data (orange), ProMED (blue) and HealthMap (green) data. The shaded regions depicts the 95% credible inte1r4vals (95% CrI) for the *R*^*t*^ estimates. The reproduction number was estimated on sliding windows of 28 days with a Gamma prior with mean 1 and variance 0.25, using the R package EpiEstim [20]. Estimates shown at time *t* are for the 28-day window finishing on day *t*.

**Figure 2:**
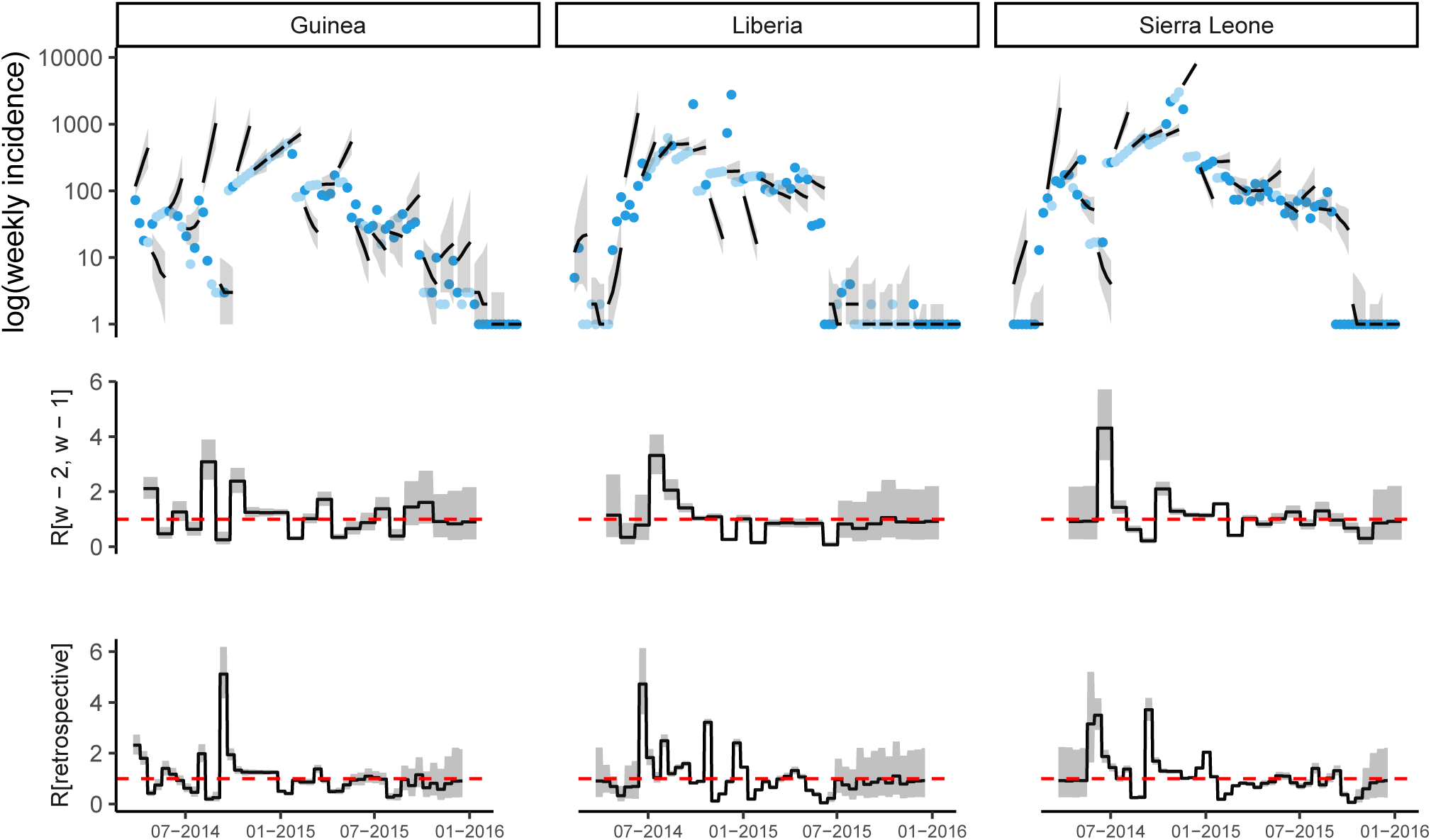
Observed and predicted incidence, and reproduction number estimates from ProMED data. The top panel shows the weekly incidence derived from ProMED data and the 4 weeks incidence forecast on log scale. The solid dots represent the observed weekly incidence where the light blue dots show weeks for which all data points were obtained using interpolation. The projections are made over 4 week windows, based on the reproduction number estimated in the previous 2 weeks. The middle figure in each panel shows the reproduction number used to make forecasts over each 4 week forecast horizon. The bottom figure shows the effective reproduction number estimated retrospectively using the full dataset up to the length of one calibration window before the end. In each case, the solid black line is the median estimate and the shaded region represents the 95% Credible Interval. The red horizontal dashed line indicates the *R*_*t*_ = 1 threshold. Results are shown for the three mainly affected countries although the analysis was done jointly using data for all countries in Africa.

To broadly assess the extent to which such discrepancies in incidence would impact the quantification of transmissibility throughout the epidemic, we estimated the time-varying transmissibility, measured by the reproduction number *R*^*t*^ (the average number of secondary cases at time *t* per infected individual [19]), using the incidence from each of the three data sources (Fig 1). *R*^*t*^ was estimated independently for each country using the R package EpiEstim [20] over a sliding time window of 4 weeks. There were substantial differences in the estimates of *R*^*t*^ according to the incidence data source used (Fig 1B). The correlation between the median *R*^*t*^ estimates on sliding 4-week windows from ProMED or HealthMap data with the estimates from WHO data varied from weak (0.30, between reproduction number from WHO and ProMED in Guinea) to very strong (0.72, between reproduction number from WHO and ProMED in Sierra Leone, Fig 3).

**Figure 3:**
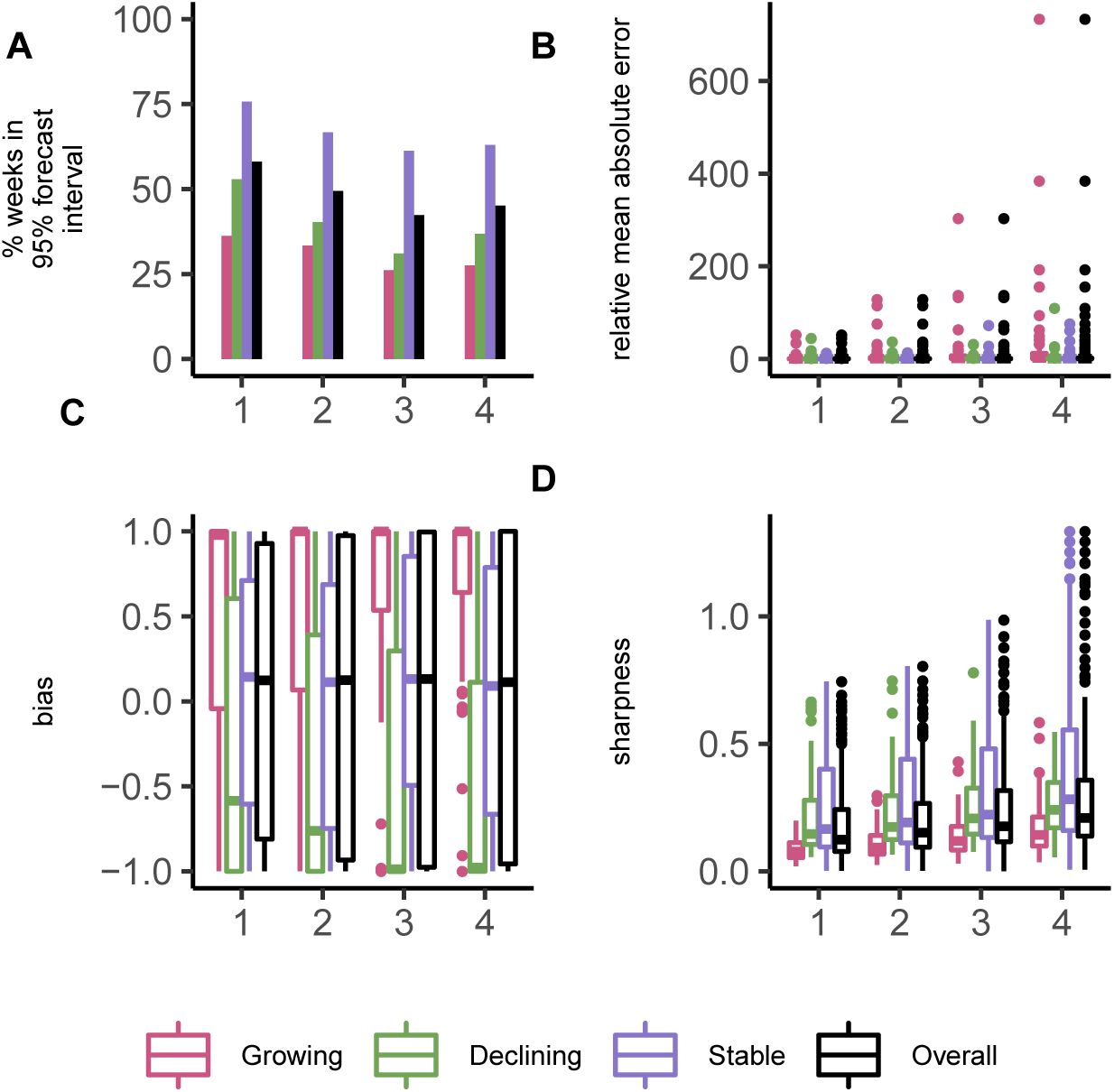
Model performance metrics overall and stratified by week of projection and epidemic phase. The performance metrics are (A) the percentage of weeks for which the 95% forecast interval contained the observed incidence, (B) relative mean absolute error, (C) bias, and (D) sharpness. In forecasting ahead, we assumed transmissibility to be constant over the forecast horizon. If the 97.5^th^ percentile of the R estimate used for forecasting was less than 1, we defined the epidemic to be in the declining phase during this period. Similarly, if the 2.5^th^ percentile of R was greater than 1, we defined the epidemic to be in a growing phase. The phase was set to stable where the 95% Credible Interval of the R estimates contained 1. See Methods for definitions of each metric.

Since HealthMap uses ProMED alerts in addition to other online data sources, ProMED represents the more conservative data source between the two. Therefore we present the results based on ProMED data in the main text, unless otherwise specified. The analysis based on HealthMap and WHO data are presented in the Supplementary Information (SI Sections 3 and 4).

### Short term forecasts

To forecast incidence at time *t* in location *j* 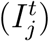, we fitted a spatially explicit branching process model to the daily incidence in all locations up to *t* − 1, using an extension of the renewal equation as the likelihood [19]:

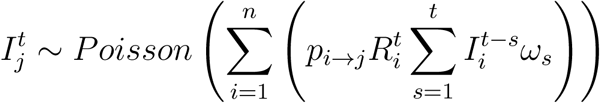

where the matrix *p*_*i*→*j*_ characterises the spatial spread between locations *i* and *j* based on a gravity model [21], 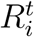 (the reproduction number) reflects transmissibility in location *i* at time *t*, and *ω* is the typical infectiousness profile of a case over time after infection (see Methods for details).

The ability of the model to robustly predict future outbreak trajectory was limited and depended on the data source (Fig 2) as well as on the time window used for inference (calibration window) and the forecast horizon. Results using a 2-week calibration window and a 4-week forecast horizon using ProMED data are presented in the main text (see Figs 6 and 7 for other forecast horizons and calibration windows). Overall, 48.7% of weekly observed incidence across all three countries were included in the 95% forecast interval (49.3% and 57.5% for HealthMap and WHO respectively, SI Table 1). Typically, model forecasts were 0.5 times lower or higher than the observed incidence (95% CrI 0.0 - 32.0) based on the median relative mean absolute error (Fig 3D), see Methods for details). We found no evidence of systematic bias in any week of the forecast horizon (median bias 0.12, Fig 3A).

Typically, individual forecasts were within 17.0% (95% CrI 0 - 80%) of the median forecast (based on the median and 95% CrI for relative sharpness, Fig 3B, see Methods for details).

As expected, the robustness of forecasts (both accuracy and precision) decreased over the forecast horizon (Fig 3C). In the first week of the forecast window, 58.0% of observed values (across the three countries) were within the 95% forecasts interval, reducing to 49.4%, 42.3% and 45.0% in the second, third and fourth weeks of the forecast horizon.

The model performance varied depending on the country and the phase of the epidemic, defined as “growing”, “declining”, and “stable” depending on *R*^*t*^ estimates (see Methods). In general, the model performance was best in the stable phase with 66.7% of the observations contained in the 95% forecasts interval (versus 40.2% and 30.8% in the declining and growing phases respectively, SI Table 1). However the forecast uncertainty was largest in the stable phase and smallest in the growing phase (Fig 3B). Importantly, in the growing phase the model tended to over-predict while under-predicting in the declining phase (Fig 3A).

Overall, the model performed moderately better using WHO data compared to ProMED and HealthMap data (Fig 33) and with shorter calibration windows (Fig 32).

### Risk of spatial spread

Although our model was not always able to robustly predict the future incidence in the three mainly affected countries, we found that it allowed to robustly predict the presence or absence of cases in all countries in Africa up to a week in advance. For each week and each country in Africa, our model generated an alert if the predicted incidence (using a predetermined percentile of the forecast interval) was greater than 0. We classified an alert for a given week as a true alert when the observed incidence was also greater than 0, as a false alert when no cases were observed, and as a missed alert if cases were observed but were not predicted by the model. Using different percentiles of the forecast (e.g., the median or the 95^th^ percentile) yielded different rates of true, false and missed alerts, which were summarised in a ROC curve (Fig 4A). Overall, our model achieved high sensitivity (i.e., true alert rate) but variable specificity (i.e., 1 - false alert rate). Maximising the average between sensitivity and specificity led to 93.7% sensitivity and 96.0% specificity at 42.5% threshold (Fig 4A). The sensitivity of the model remained high over longer forecast horizon while the specificity deteriorated with more false alerts being raised 4 week ahead (Fig 47). Both the sensitivity and the specificity of the model remained high when the analysis was restricted to all countries in Africa other than the three majorly affected countries (Guinea, Liberia and Sierra Leone). The average of sensitivity and specificity was maximum at 92.5% threshold in this case with 85.7% sensitivity and 81.7% specificity (Fig 48). The model exhibited high sensitivity (83.3%) and specificity (82.0%) in predicting presence of cases in weeks following a week with no observed cases in all countries in Africa (Fig 49) at 93% threshold (similarly chosen to maximise the average between sensitivity and specificity). Out of the 9 one-week ahead missed alerts in this case, 3 are in Liberia towards the end of the epidemic, after Liberia had been declared Ebola free on two separate occasions [22, 23]. The serial interval distribution that we have used does not account for very long intervals between infections such as that associated with sexual transmission. Using the latest available data on pairs of primary and secondary infections and models that allow for more heterogeneity in the distribution of cases e.g., using Negative Binomial distributions could potentially improve the assessment of risk of spread in such cases [24, 25].

**Figure 4:**
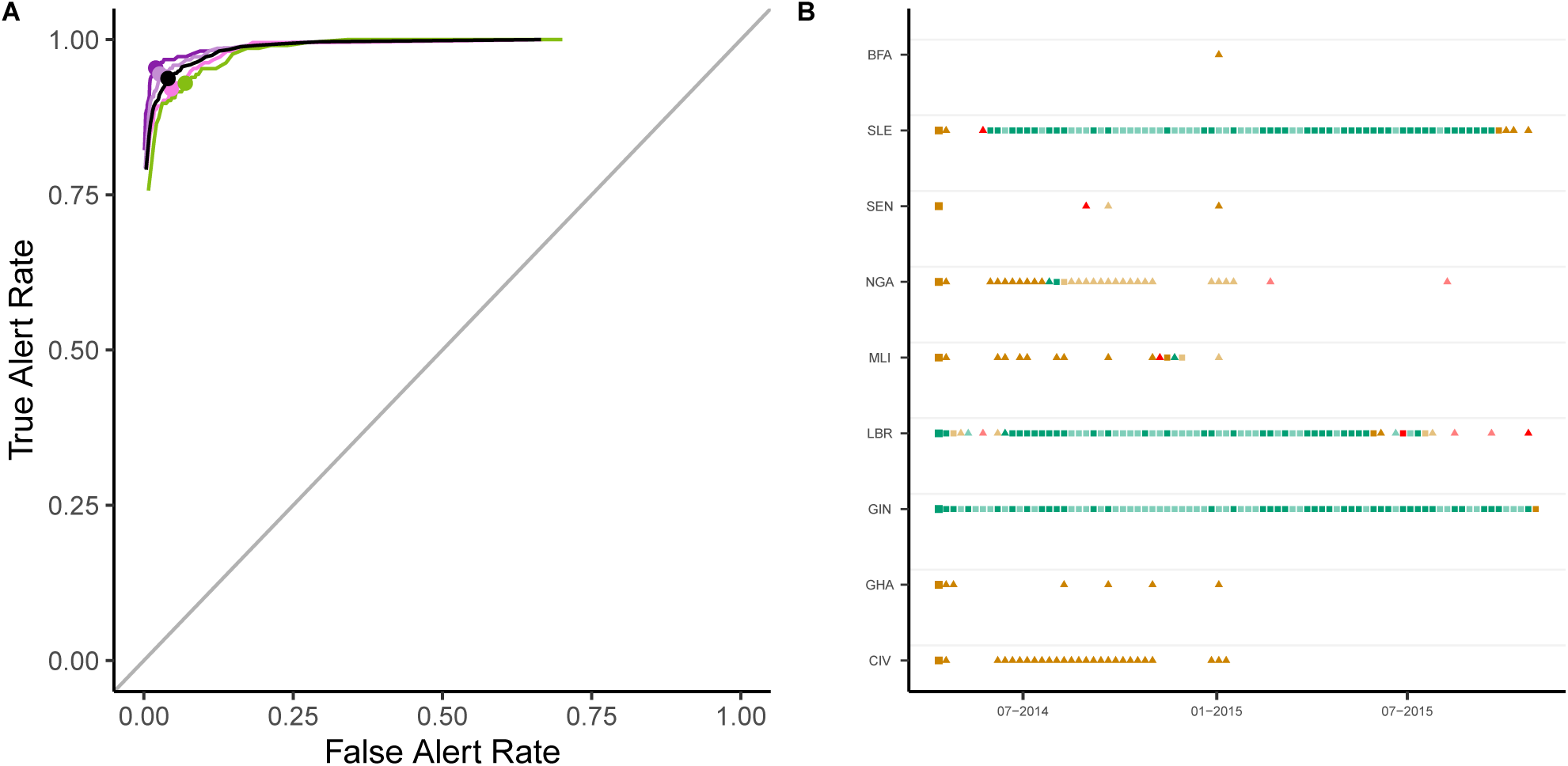
Predicted weekly presence of cases in each country. The left panel shows the True and False alert rates using different thresholds for classification for alerts raised 1 (violet), 2 (light violet), 3 (dark pink) and 4 (green) weeks ahead. The black curve depicts the overall True and False alert rates. On each curve, the dot shows the True and False Alert rates at 42.5% threshold. For a given threshold (*x*^*th*^ percentile of the forecast interval), we defined a True alert for a week where the *x*^*th*^ percentile of the forecast interval and the observed incidence for a country were both greater than 0; false alert for a week where the threshold for a country was greater than 0 but the observed incidence for that country was 0; and missed alert for a week where the threshold for a country was 0 but the observed incidence for that country was greater than 0. True alert rate is the ratio of correctly classified true alerts to the total number of true and missed alerts (i.e., (true alerts)/(true alerts + missed alerts)). False alert rate is similarly the ratio of false alerts to the total number of false alerts and weeks of no alert (where the observed and the threshold incidence are both 0). The right panel shows the True (green), False (orange) and Missed (red) 1 week ahead alerts using the 42.5^th^ percentile of the forecast interval as threshold. The figure only shows countries on the African continent for which either the 42.5^th^ percentile of the predicted incidence or the observed incidence was greater than 0 at least once. The first alert in each country is shown using larger squares. Alerts in a country in a week where there were no observed cases in the previous week are shown using triangles. In each case, weeks for which all observed points were imputed are shown in lighter shades. Country codes, shown on the y-axis, are as follows: CIV - Côte d’Ivoire, GHA - Ghana, GIN - Guinea, LBR - Liberia, MLI - Mali, NGA - Nigeria, SEN - Senegal, SLE - Sierra Leone, BFA - Burkina Faso. The alerts are based on forecasts using ProMED data, a 2-week calibration window and a 4 week forecast horizon.

The choice of a threshold at which to raise an alert is subjective and involves a trade-off between sensitivity and specificity. In general, using a high threshold to raise an alert leads to high sensitivity with a reasonably high specificity (Fig 50). The cut-off for raising an alert can be informed by the relative costs of missed and false alerts potentially using a higher cut-off when the observed incidence is low. It is also worth noting that where our model failed to raise an alert in a week, either a true or a false alert had been raised in the recent weeks in all but one instance, indicating a potential risk of spread of the epidemic to that country.

These results suggest that the model is able to capture and even anticipate the spatial spread of the epidemic. Importantly, as the model is fitted to data from any of the three data sources accumulating over the course of the epidemic, it is able to predict the presence of cases relatively early in the epidemic (Fig 51) when such inputs would be particularly useful.

Together with providing operational outputs such as the predicted short-term incidence in currently affected countries or the risk of spread to neighbouring countries, our method also provides near real-time estimates of parameters underlying the transmission model. First, the reproduction number *R*_*t*_ for each affected country is estimated over the time-window of inference, here over the most recent two weeks (Fig 2), second row). We found that these near real-time estimates of *R*_*t*_ were in good agreement with retrospective estimates obtained using the entire incidence time-series (Fig 2, bottom row, correlation coefficients varying between 0.58 and 0.90, Fig 5).

**Figure 5:**
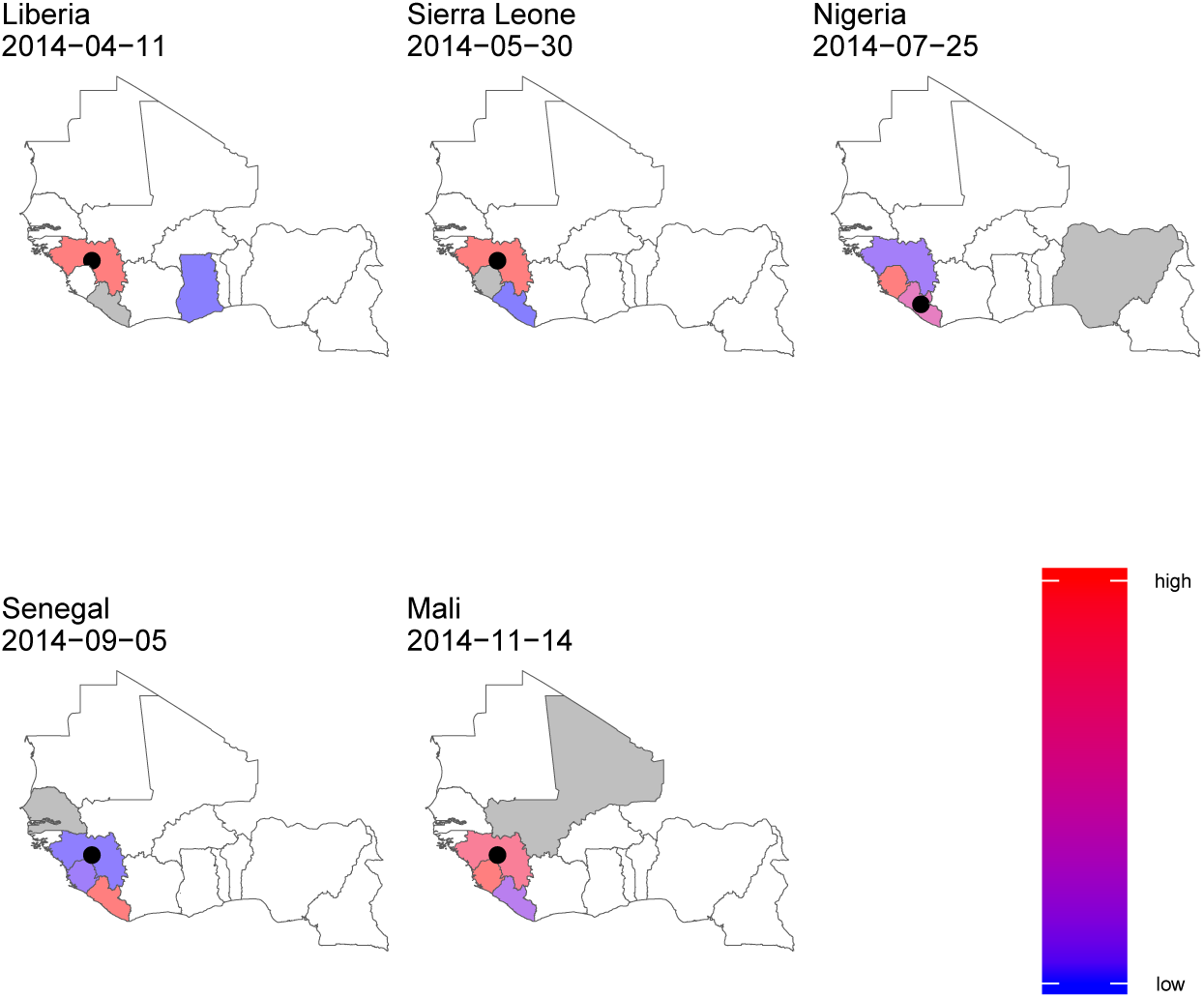
Relative risk of importation of the epidemic. For each country with non-zero incidence, the figure shows the relative importation risk (see Methods). Since we forecasted every 7 ^th^ day, the risk of importation was estimated using forecasts closest to and before the date of the first case in that country reported in the data used. The date on which risk was estimated for each country is shown in the figure. Blue indicates low relative risk while deeper shades of red represent higher relative risk of acting as a source of importation. White is used to denote no risk. The estimates presented here use ProMED data with a 2 week calibration window. The country for which risk is estimated is shown in gray. The black circle represents the actual source of importation as retrospectively identified through epidemiological and genomic investigations. For each country, the figure shows only the risk of importation from other countries and does not show the risk of transmission within the country.

The risk of spatial spread in our model relies on estimating movement patterns of infectious cases. Our method also provides estimates of the probability of a case staying within a country throughout their infectious period, and the extent to which distance between two locations affects the flow of infectious cases between them. The real-time estimates of these parameters over the course of the epidemic (Fig 34), suggest that while the relative flow of cases between locations did not vary substantially over time, the probability of travel across national borders may have decreased after the initial phase of the epidemic.

Finally, we quantify the relative risk of importation of a case into a country from any other currently affected country. The risk of importation is proportional to the population flow into a country from all other countries estimated using a mobility model (here, gravity model) weighted by the infectivity at each country (which depends on the number of cases and time at which they were infected, see Methods). Our estimates of the countries with higher risk of acting as source of importations are largely consistent with the reported source of cases (Fig 5). In 4 out of the 5 reported cases of international spread of the epidemic in West Africa, the model correctly attributed the highest relative risk of acting as a source of importation (relative risk *>* 0.9 for importation into Liberia, Nigeria and Sierra Leone and 0.49 in case of Mali) to the actual source identified through retrospective epidemiological and genomic investigations [26].

## Discussion

In the context of increasing potential for movement of diseases between various regions of the world due to increased global connectivity, innovative strategies for epidemic monitoring are urgently needed. In this study, we propose a statistical framework that relies on digital surveillance data from ProMED or HealthMap to 1) predict the short-term epidemic trajectory in currently affected countries, 2) quantify the short-term risk of spread to other countries and 3) for countries at risk of importation, quantify where the risk comes from. We apply our model to data collected during the West African Ebola epidemic of 2013-2016, curated by the outbreak analysts at ProMED/HealthMap, and we compare the model’s outputs to those obtained when using the data collated by the WHO and made available at the end of the epidemic.

In spite of the manual curation of the data carried out by outbreak analysts at ProMED and HealthMap, substantial issues remained in the quality and consistency of the data feeds. Dealing with issues such as missing data and inconsistent records will be a key challenge in using these data for prospective real-time analysis. Despite these challenges, we show the potential of digital surveillance tools to 1) inform early detection, 2) characterise the spread, and 3) forecast the future trajectory of outbreaks. Particularly in an evolving outbreak scenario, when information from traditional surveillance is limited and only available with a significant delay, digital surveillance data can be used to complement the information gap. For instance, during the West African Ebola epidemic, the first situation report by the WHO was published only at the end of August 2014 [27], reporting on cases between January and August 2014. On the other hand, the first post on ProMED on Ebola cases in Guinea appeared in March 2014 [28]. Development of tools that can directly be plugged into the current digital surveillance ecosystem should therefore be a growing area of focus.

In general, we found that, after systematic processing to remove inconsistencies, data from ProMED and HealthMap were in reasonably good agreement with the data collected through traditional surveillance methods and collated by WHO. In particular, both the incidence time-series and retrospective national estimates of transmissibility over time were well correlated across the three data sources. This suggests that digital surveillance data are a promising avenue for quantitative assessment of outbreak dynamics in real-time.

We used the ProMED/HealthMap data to perform a spatio-temporal analysis of the spread of the West African Ebola epidemic. We fitted a spatially explicit branching process model to the daily incidence data derived from ProMED/HealthMap feeds. The estimated model parameters were used to simulate the future outbreak trajectory over 4, 6 or 8 weeks. The model performs relatively well at short forecast horizon, i.e. up to two weeks. At a longer time scale, the model performance starts to deteriorate. A likely explanation for this is that our model assumes that transmissibility remains constant over the entire projection window. This assumption may not hold as we project over longer horizons, for example due to interventions being implemented. Model performance was also highly dependent on the phase of the epidemic in which projections were made. During the growing phase, the model tended to over-predict. This is likely due to interventions implemented throughout the growing phase to reduce transmissibility, leading to a reduced observed incidence compared to our model’s expectation. In the declining phase on the other hand, our model tended to under-predict the observed incidence. This could be due to control efforts being relaxed too early as case numbers decline. Another contributing factor could be super-spreading, a phenomenon observed in many epidemics including Ebola epidemics [29,30] whereby a small number of cases generate a large number of secondary infections, implying that when case numbers are small, epidemic trajectory may be difficult to predict and not well described by Poisson likelihood. Models using more complex likelihoods, e.g. using Negative Binomial distributions, should be explored in future work, but will present additional challenges as analytical results underpinning the estimation of the reproduction number will no longer hold [20].

Such variability in model performance throughout an epidemic could have important implications if the model predictions are used to inform resource allocation. Model estimates should therefore be interpreted with caution, particularly as an outbreak is observed to be declining, and if the forecast horizon is long.

We have shown that our model would have been able to accurately predict in real-time the international spread of Ebola in West Africa. Importantly, our model has very high sensitivity, predicting all instances of observed international spread 1 to 4 weeks in advance. Choosing a cut-off to maximise sensitivity led to low model specificity. On occasions the model predicted cases in countries, such as Côte d’Ivoire, where neither WHO nor ProMED reported any case. However this may also be due to imperfect case reporting. Thus our method could be used with a high cut-off as a highly sensitive surveillance system with an alert triggering further epidemiological investigations and implementation of epidemic preparedness measures.

A key feature of our model is that it provides, for each country identified as at risk, a map of where the risk comes from. Out of 5 observed instances of international spread of Ebola in West Africa, our model correctly identified the source of importation in 4 cases while in the remaining case, the model highlighted the source of importation while assigning it low relative risk. This could help translating data collected through digital surveillance into concrete operational outputs in real-time that could assist in epidemic management and control.

The systematic collection, storage, organization and communication of disease surveillance data were especially challenging during the West African Ebola epidemic as the deficiencies in transportation and communication resources, surveillance data quality and management, human resources and management structures posed unique challenges in this context [31]. The collection of case incidence data and rapid dissemination through digital surveillance systems was further hampered by the limited information technology and internet service in the countries most affected. Thus, for the West African Ebola epidemic, ProMED and HealthMap data were available at a coarse spatial scale with the sub-national level information for cases missing in most of the records. This limited our analysis to the spread of the outbreak across national borders only, although in principle our model could deal with data at any spatial scale. Both ProMED and HealthMap collect more granular data for most outbreaks. Utilizing these data to investigate outbreaks and regions would provide further evidence of the ability of digital surveillance data to usefully complement data collected from traditional surveillance. Another interesting research avenue would be to explore ways of integrating timely data from ProMED and HealthMap with delayed data from ground surveillance to generate timely insights into the spatial spread of an outbreak.

The framework presented in this paper was developed as a proof-of-concept to use digital surveillance data for near real-time forecasting of the spatio-temporal spread of an outbreak. It has been implemented as a web-based tool called “Mapping the Risk of International Infectious Disease Spread” (MRIIDS) (see [32] for more information). To further develop such approaches, it is important to establish an automated pipeline from data collection to curation to analysis, which currently requires manual intervention at each of these steps. Another factor that could enhance the usability of our model in near real time is to improve the running time of the fitting and forward simulation. In the current implementation, the running time varies from approximately 0.5 CPU hours when 100 days of incidence data are being used to approximately 335 CPU hours using 462 days of incidence data using a 3.3 GHz Intel Xeon X5680 processor. Although the West African Ebola epidemic was of unusual scale and duration, there is a scope for optimising the model implementation.

Importantly, many other open data sources could be included in our framework to improve model performance. For example, data on human mobility could be used to further inform the parametric form and parameter values of our mobility model. We have incorporated a simple and well-characterised model of population movement in the current work. In addition to utilising other possible data sources, future work could consider other models of human population movement [33]. When relevant, spatially-explicit data on population-level immunity to the circulating pathogen (e.g. following previous epidemics and/or due to vaccination) could also be used to refine our transmission model. Finally, the impact of the health capacity of a region to respond to a public health emergency could also be accounted for in future iterations of the model. Ongoing efforts to collate quantitative information on the performance of health systems and the ability of regions or countries to respond to an epidemic [34], [35] can potentially provide valuable data for future work. Here using a relatively simple modelling approach we provide one of the first pieces of evidence of the potential value of digital surveillance for real-time quantitative analysis of epidemic data, with important operational and actionable outputs.

## Figures

### Supporting Information (SI)

The following files are provided as Supporting Information: (i) SI 1. Details of ProMED/HealthMap data cleaning and processing, model outputs for other data sources, model parameters and sensitivity analysis. (ii) Convergence report produced by the R package ggmcmc.

## Materials and Methods

### Processing ProMED/HealthMap data feed

We used a set of curated ProMED and HealthMap records on the human cases of Ebola in West Africa. The dates in the feeds ranged from March 2014 to October 2016. Each dataset recorded the cumulative number of suspected/confirmed cases and suspected/confirmed deaths by country at various dates in this period. We derived the country specific daily and weekly incidence time series from these data after the following data cleaning and pre-processing:

- We first extracted the total case counts as a sum of suspected and confirmed cases (ProMED and HealthMap data did not record probable cases for the West African Ebola epidemic).
- Each unique record in ProMED is associated with a unique alert id. An alert id can correspond to various reports from different sources (news, social media etc.) which might report different case numbers. In such cases, we assigned the median of the case numbers to the record.
- In some instances, cumulative case count was inconsistent in that it failed to be monotonically non-decreasing. We identified consecutive dates (*t*_*k*_ and *t*_*k*+1_) where the cumulative case count was not increasing. If removing either *t*_*k*_ or *t*_*k*+1_ made the cumulative case count increasing, we adopted this option. If however removing both or none of them resulted in a increasing series, we removed both *t*_*k*_ and *t*_*k*+1_. These rules were applied iteratively until the cumulative case count was consistent.
- If an inconsistent point was at the end of the the cumulative case series, applying the above rules led to the removal of a large number of points. Hence, to remove outliers at the end, we used Chebyshev inequality with sample mean [36]. Given a set of observations *X*_1_, *X*_2_ … *X*_*n*_, the formulation of Chebyshev inequality by Saw et al. gives the probability that the observation *X*_*n*+1_ is within given sample standard deviations of the sample mean. We defined *X*_*n*+1_ to be an outlier if the probability of observing this point given observations *X*_1_, *X*_2_ … *X*_*n*_ is less than 0.5. Fixing this probability allowed us to determine *k* such that *Pr*(*µ* − *kσ* ≤ *X*_−_ {*n* + 1} ≤ *µ* + *kσ*) ≥ 0.5, where *µ* and *σ* are the sample mean and sample standard deviation respectively. We deleted an observation *X*_*n*+1_ as an outlier if it did lie in this interval.
- Finally, cumulative incidence on days with no records was filled in using log-linear interpolation.

An example of the pre-processing of ProMED feed is presented in the supplementary text (SI Fig 1).

### Data collated by WHO

We used the West African Ebola incidence data collated by the WHO during the epidemic which was made available approximately an year after the end of the epidemic [37]. This data set is referred to as “WHO data” in the interest of brevity. The cleaned version of the WHO data consisted of cases reported between December 2013 and October 2015 in the three most affected countries - Guinea, Sierra Leone and Liberia. The location of residence of cases was geo-coded to the second administrative level. We aggregated the WHO data to national level to match the spatial resolution of ProMED and HealthMap that were only available at the country level.

Since the data collected by ProMED and HealthMap are manually curated by outbreak analysts, we have used the word “curate” in referring to their data collection process. Similarly, we refer to the data “collated” by WHO as WHO coordinated the international response to the outbreak and in this role, they collated the information from Ministries of Health, situation reports from NGOs, and local and international medical teams.

### Demographic Data

We used LandScan™ 2015 dataset grid [38] for population estimates and centroids for all countries on African mainland.

### Statistical Model

Our model relies on a well-established statistical framework that assumes the daily incidence, *I*_*t*_, can be approximated with a Poisson process following the renewal equation [19]:

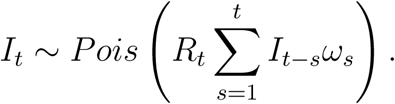

Here *R*_*t*_ is the reproduction number at time *t* (the average number of secondary cases per infected individual) and *ω* is the distribution of the serial interval (the time between onset of symptoms in a case and their infector).

We extend this model to incorporate the spatial spread of the outbreak between *n* different locations. The number of incident cases at a location *j* at time *t* is given by the equation

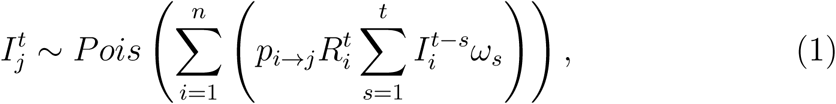

where 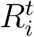 is the reproduction number at location *i* at time *t* and *p*_*i*→*j*_ is the probability of a case moving from location *i* to location *j* while they are infectious. 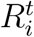 is affected by a number of factors e.g., the intrinsic transmissibility of a pathogen or the health care capacity at location *i* (which could influence for example the capacity to isolate cases). The model could be easily extended to explicitly incorporate the dependence of the reproduction number on such factors.

The probability of moving between locations is derived from the relative flow of populations between locations. This latter quantity can be estimated using a population flow model; here we used a gravity model [39], [21]. Under a gravity model, the flow of individuals from area *i* to area *j, ϕ*_*i*→*j*_, is proportional to the product of the populations of the two locations, *N*_*i*_ and *N*_*j*_ and inversely proportional to the distance between them *d*_*i,j*_ raised to a power *γ*:

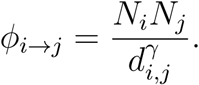

The exponent *γ*, which modulates the effect of distance on the flow of populations, was estimated. A large value of *γ* indicates that the distances travelled by populations tend to be short.

The relative risk of spread at a location *j* from a location *i* is thus proportional to the population flow into location *j* from location *i*.

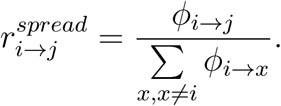

The probability of movement from location *i* to location *j* where (*j* ≠ *i*)is then given by

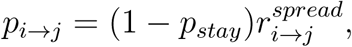

where *p*_*stay*_ is the probability that a case remains in a location *i* throughout their infectious period i.e. *p*_*stay*_ is *p*_*i*→*i*_. *p*_*stay*_ is assumed to be the same across all locations.

The parameters of the full model are:

1. 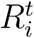, the reproduction number at time *t* in location *i*,
2. *p*_*stay*_, the probability of staying in a location, and
3. *γ*, the exponent of the distance in the gravity model.

The likelihood of the incidence at time *t* in location *j* given past incidence across all locations and model parameters is:

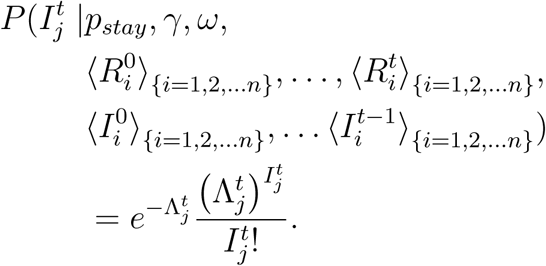

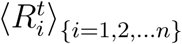 is the set of reproduction numbers at time *t* in locations 1, 2, … *n*. 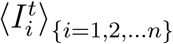 is similarly the incidence at time *t* in locations 1, 2, … *n*. 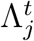 is given by

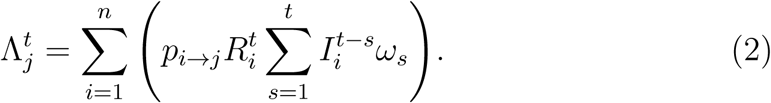

The likelihood of the parameters for data up to time *T*, the duration of the outbreak so far, is

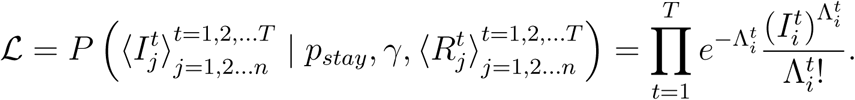

Therefore the joint posterior distribution of the model parameters given the observed data is:

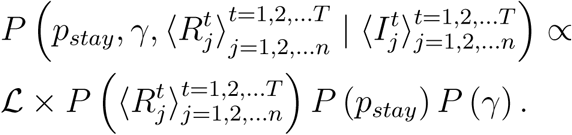

*P* (*p*_*stay*_), *P* (*γ*), and 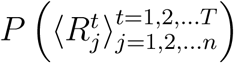 represent the prior distributions on the parameters.

Here, we have assumed a single *p*_*stay*_ for all countries on African mainland. For estimating the time-varying reproduction number for each country, we split the duration of the total outbreak into intervals of equal width. We assume that transmissibility in each location stays constant within each time window and thus, within a time window, we estimated a single reproduction number for each location. We varied the length of the time window to obtain different models, with short time windows increasing the number of parameters in the model. To reduce the number of parameters in the model, we divided the 55 countries on African continent into 5 groups and forced each country in a group to have the same reproduction number in each time window. The first three groups correspond of the three mainly affected countries - Sierra Leone, Guinea and Liberia. The countries that shared a border with these three countries were grouped together. These were Mali, Côte d’Ivoire, Guinea-Bissau and Gambia. The rest of the countries were assigned to the fifth group. A comparison of the performance of different models is presented in the Supplementary Material (Fig 32).

We assumed a Gamma distributed serial interval with mean 14.2 days and standard deviation 9.6 days [18]. For the reproduction number, we used a Gamma prior with mean 3.3 and variance 1.5. This was informed by a review of estimates of the reproduction number for Ebola Zaire in outbreaks preceding the West African Ebola outbreak which reported estimates ranging from 1.4 to 4.7 [40]. The mean prior 3.3 was chosen as the midpoint of this interval, and the variance 1.5, was chosen so the 95% prior probability interval contains the extremes of this interval.

For the gravity model parameters *p*_*stay*_ and *γ*, we chose uninformative uniform priors. Since *p*_*stay*_ is a probability, the prior was a uniform distribution on the interval [0, 1]. *γ* was allowed to vary between 1 and 2 in the results presented in the main text. We performed a sensitivity analysis where *γ* has a uniform prior between 1 and 10. The results of this analysis are presented in the SI (SI Section 8).

Model fitting was done in a Bayesian framework using a Markov Chain Monte Carlo (MCMC) as implemented in the software Stan [41] and its R interface rstan [42]. We ran 2 MCMC chains with 3000 iterations and burn-in of 1000 iterations. Convergence of MCMC chains was confirmed using visual inspections of the diagnostics (Potential Scale Reduction Factor [43] and Geweke Diagnostics [44]) reported by R package ggmcmc [45]. An example report produced by ggmcmc is included in the Supplementary Material.

For each model (i.e., for each choice of the time window), we made forward projections every 7^th^ day, over a 2 week, 4 week and 6 week horizon. To forecast incidence from day *t* onwards, we fitted the model to the daily incidence series up to day *t* − 1. We then sampled 1000 parameter sets (reproduction numbers for each location in each time window and parameters of the gravity model) from the joint posterior distribution, and for each parameter set, simulated one future epidemic trajectory according to equation (1), assuming that future *R*_*t*_ is equal to the last estimated *R*_*t*_ value in each location.

## Model Validation

Given the retrospective nature of our analysis, we validated the incidence projected using our model against observed incidence. In addition to the accuracy of the forecasted incidence, the uncertainty associated with the forecasts (e.g., measured by the width of the prediction interval) is an important indicator of model performance. A narrow prediction interval that contains the observed values is preferable over wide prediction intervals. To assess the performance of the model along both these dimensions, we used four different metrics drawn from the literature.

In the remainder of this paper, we use the following notation. For a location *j*, let 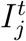 be the observed incidence at time *t* and let 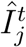 be the set of predictions of the model at time *t*. That is, 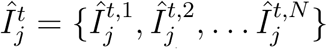 is the set of *N* draws from the Poisson distribution with mean 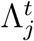 (Equation 1) (here N = 1000).

### Relative mean absolute error

The relative mean absolute error (rmae) is a widely used measure of model accuracy [46]. The relative mean absolute error for the forecasts at a location *j* at time *t* is defined as:

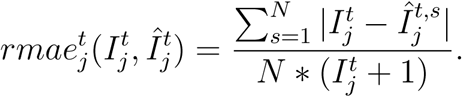

That is the mean absolute error at time *t* is averaged across all simulated incidence trajectories and normalised by the observed incidence. We add 1 to the observed value to prevent division by 0. A rmae value of *k* means that the average error is *k* times the observed value.

### Sharpness

Sharpness is a measure of the spread (or uncertainty) of the forecasts. Adapting the definition proposed by [47], we used the relative mean deviation about the median to evaluate sharpness. The sharpness 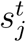 of forecasts at at time *t* at location *j* is

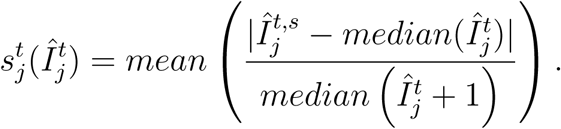

We add 1 to 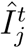 to prevent division by 0. A sharpness score of *k* indicates that the average deviation of the predicted incidence trajectories is *k* times their median. Low values of sharpness therefore suggest that the predicted trajectories are clustered around the median.

### Bias

The bias of forecasts is a measure of the tendency of a model to systematically under- or over-predict [47]. The bias of a set of predictions 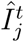 at time *t* at location *j* is defined as

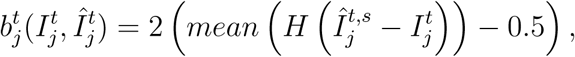

where the mean is taken across the *N* draws. *H*(*x*) is the Heaviside step function defined as

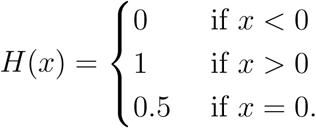

The above formulation can better be understood by considering the following extreme scenarios. If every projected value 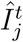 is greater than the observed value 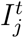, then the Heaviside function is 1 for all *i* = 1, 2, … *N*, and

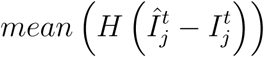

is 1. The bias for a model that always over-predicts is therefore 1. On the other hand, if the model systematically under-predicts, then

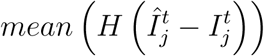

is 0 and the bias is −1. For a model for which all predictions match the observed values exactly, the bias is 0.

## Epidemic Phase

We defined the epidemic to be in a “growing” phase at time *t* if the 2.5^th^ percentile of the distribution of the reproduction number at this time was greater than 1. Similarly, the epidemic was defined to be in “declining” phase if the 97.5^th^ percentile of the distribution of the reproduction number was below 1. In all other cases, the epidemic was defined to be in a “stable” phase.

## Code and data availability

All analysis was carried out in the statistical software R version 3.5.3. The code for analysis of ProMED and HealthMap data and an implementation of the model is available at https://github.com/annecori/mRIIDSprocessData. The code for producing the text and figures for this manuscript is available at https://github.com/sangeetabhatia03/mriids_manuscript/tree/v1.0. This repository also contains the raw and processed data. Implementation of model performance metrics is at https://github.com/sangeetabhatia03/assessR.

## Relative risk of importation

The total infectivity 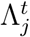 at a location *j* at time *t* is the sum of infectivity at all locations weighted by the relative flow of cases into *j* from each location *i* (Eq 2). We define the risk of importation of case from *i* into *j* as the proportion of infectivity at *j* due to infectivity at *i*. That is,

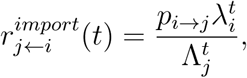

where 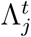 is as in Eq 2, and 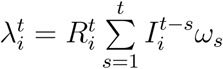.

## Data Availability

All data used in this study are available online.

https://github.com/sangeetabhatia03/mriids_manuscript/tree/v1.0

## Acknowledgement

The work in this manuscript was funded through the USAID (United States Agency of International Development) Center for Accelerating Innovation and Impact - “Combating Zika and Future Threats: A Grand Challenge for Development” Program (Award No. AID-0AA-F-16-00115). The contents of the manuscript are solely the responsibility of the authors and do not necessarily represent the official views of the USAID. SB, AC and PN acknowledge joint Centre funding from the UK Medical Research Council and Department for International Development (MR/R015600/1), funding from the UK National Institute for Health Research Health Protection Research Unit (NIHR HPRU) in Modelling Methodology at Imperial College London in partnership with Public Health England (PHE) (HPRU-2012-10080), and funding from the Bill and Melinda Gates Foundation (OPP1092240).

## Supplementary Information

### Overview

In this supplement, we present the details of the pre-processing of ProMED and HealthMap feeds (Section 1), the model results using ProMED, HealthMap and WHO data and the impact of the datasources and model parameters on the performance of the model. We varied the length of the time window used for model fitting (see Methods for details). SI Sections 2, SI Section 3 and SI Section 4 present the forecasts using ProMED, HealthMap and WHO data respectively using different calibration windows (2, 4 and 6 weeks) and forecast horizons (4, 6 and 8 weeks). The model performance was moderately better with shorter calibration windows (Fig 32) and with WHO data (Fig 33). We explored the impact of alternate priors for the gravity model parameter on the results from the model. The results of this sensitivity analysis are presented in Section 8. We present additional analysis on the predicting the spatial spread of the epidemic in Section 9.

### 1 Processing ProMED and HealthMap data

#### 1.1 Data cleaning

The data from ProMED and HealthMap was pre-processed to remove inconsistencies and address missing data (see Methods). The data on cumulative number of confirmed and suspected Ebola cases in Guinea, Liberia, Sierra Leone, Senegal, Mali, Nigeria and Ghana were extracted from ProMED and HealthMap feeds. The pre-processing workflow is illustrated using data for Sierra Leone from ProMED.

**Fig SI 1:**
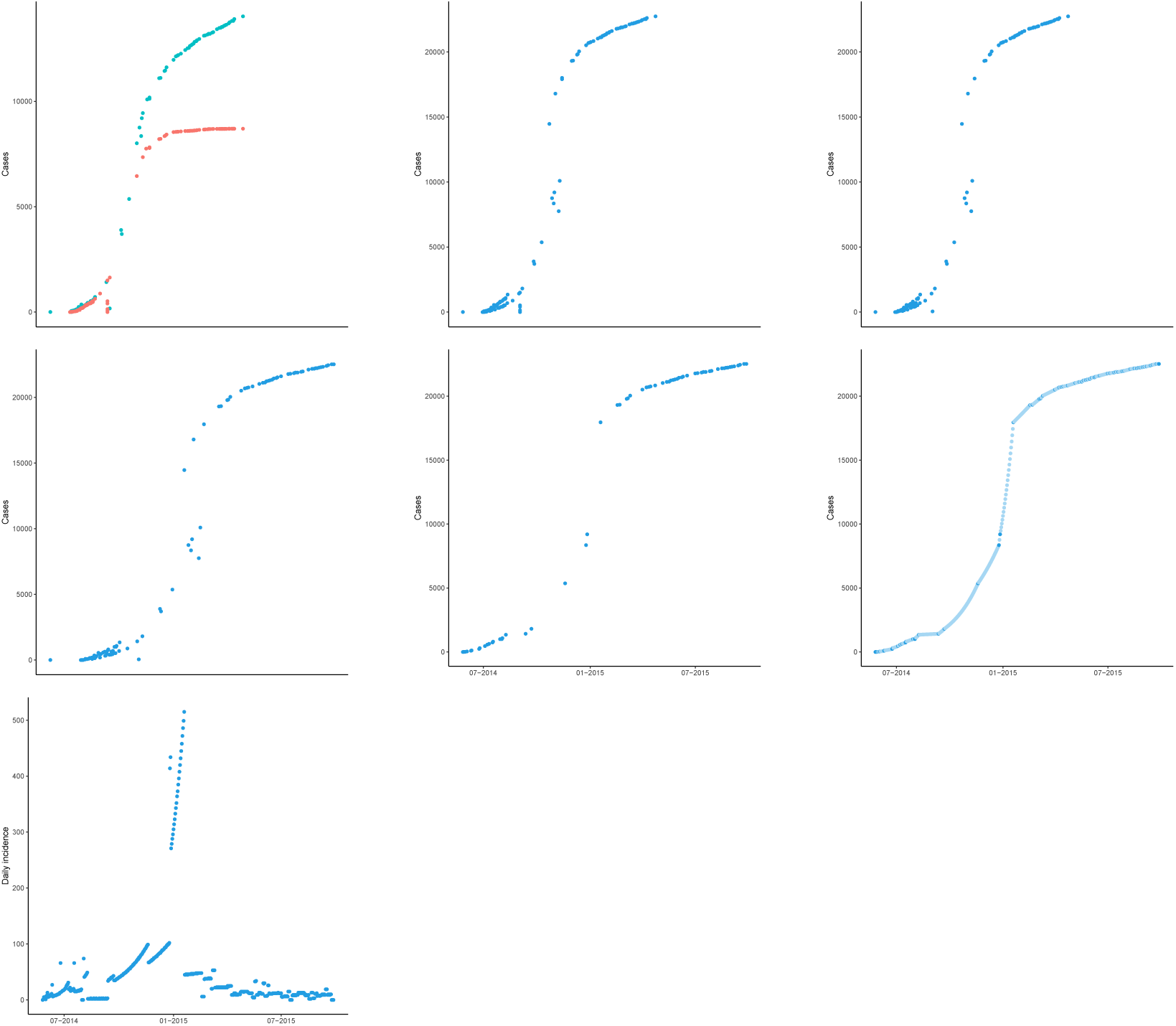
Illustration of workflow for Processing ProMED feed. Raw ProMED feed consisted of suspected and confirmed cases and suspected and confirmed deaths. The top left figure (a) shows the suspected (teal) and confirmed (red) cases in Sierra Leone in the raw feed. We used the suspected and confirmed cases to derive cumulative incidence data (b). If there were more than one alert on a day, these were then removed by assigning the median of the cases reported to this day. (c). If there were outliers in the data, we removed them in the next step (d). We then made the cumulative case count monotonically increasing (e) by removing inconsistent records. Finally, missing data were imputed using log-linear interpolation (f). Imputed points are show in light blue. See Methods for details.

#### 1.2 Weekly incidence from ProMED, HealthMap and WHO data

We processed the data from ProMED and HealthMap and derived daily and weekly incidence. The weekly incidence series derived from the three data sources were highly correlated (Fig 2).

**Fig SI 2:**
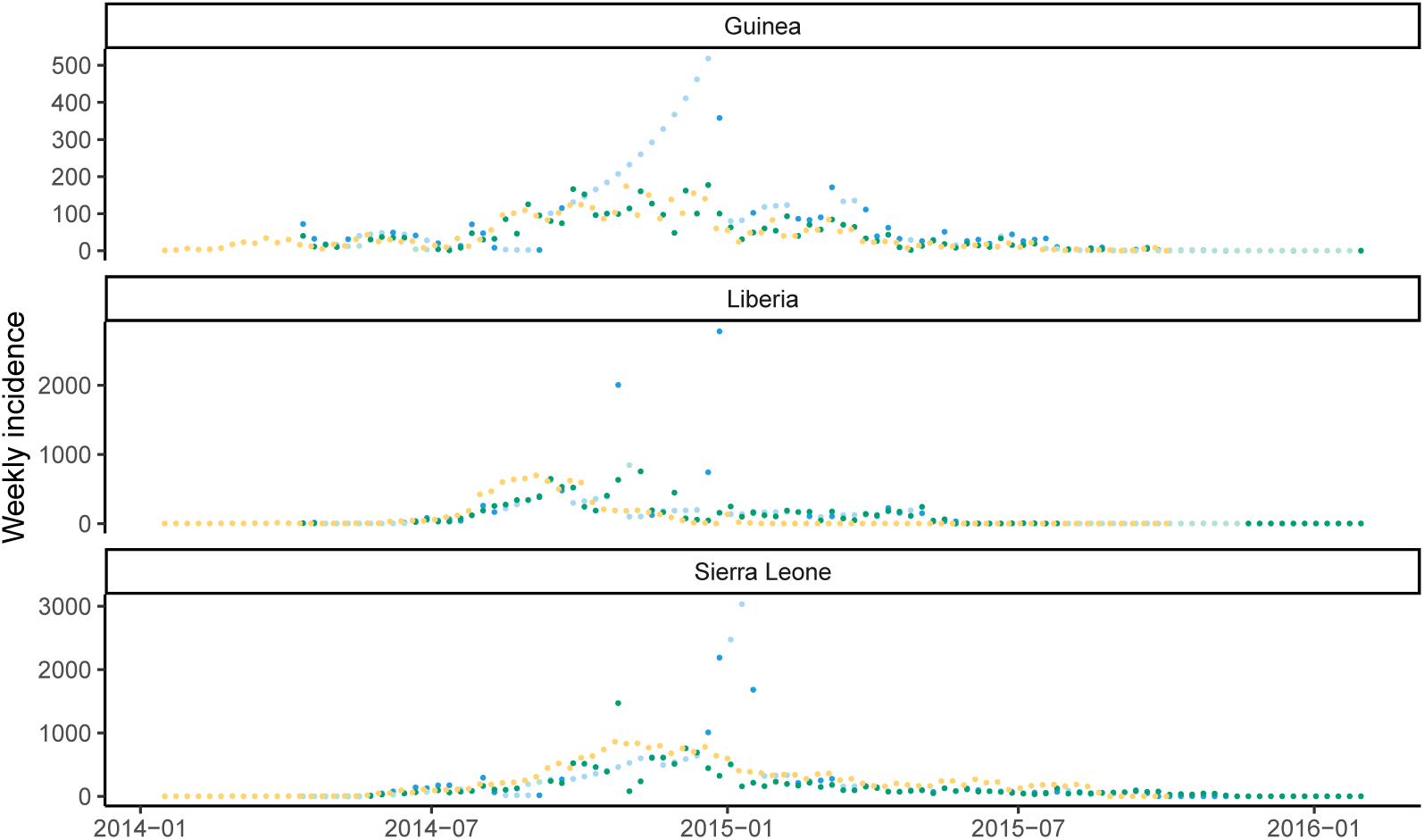
Comparison of national weekly incidence trends from ProMED (blue), HealthMap (green) and WHO (yellow) data for Guinea, Liberia and Sierra Leone. Weeks for which all daily data points were imputed are shown in lighter shades of blue and green respectively. The y-axis differs for each plot.

#### 1.3 Correlation between R estimates

The correlation between estimates of time-varying reproduction number estimated using ProMED, HealthMap and WHO data depended on the time window used for estimation and the country (Fig 3). Restricting the analysis was robust to using reproduction number estimates with lower uncertainty (coefficient of variation less than 0.25, Fig 4) Similarly, for each data source, the retrospective estimates of reproduction number in the Bayesian framework using the full incidence curve were in reasonably good agreement with the real-time estimates. The strength of the correlation varied depending on the country and the length of the calibration window (Fig 5).

**Fig SI 3:**
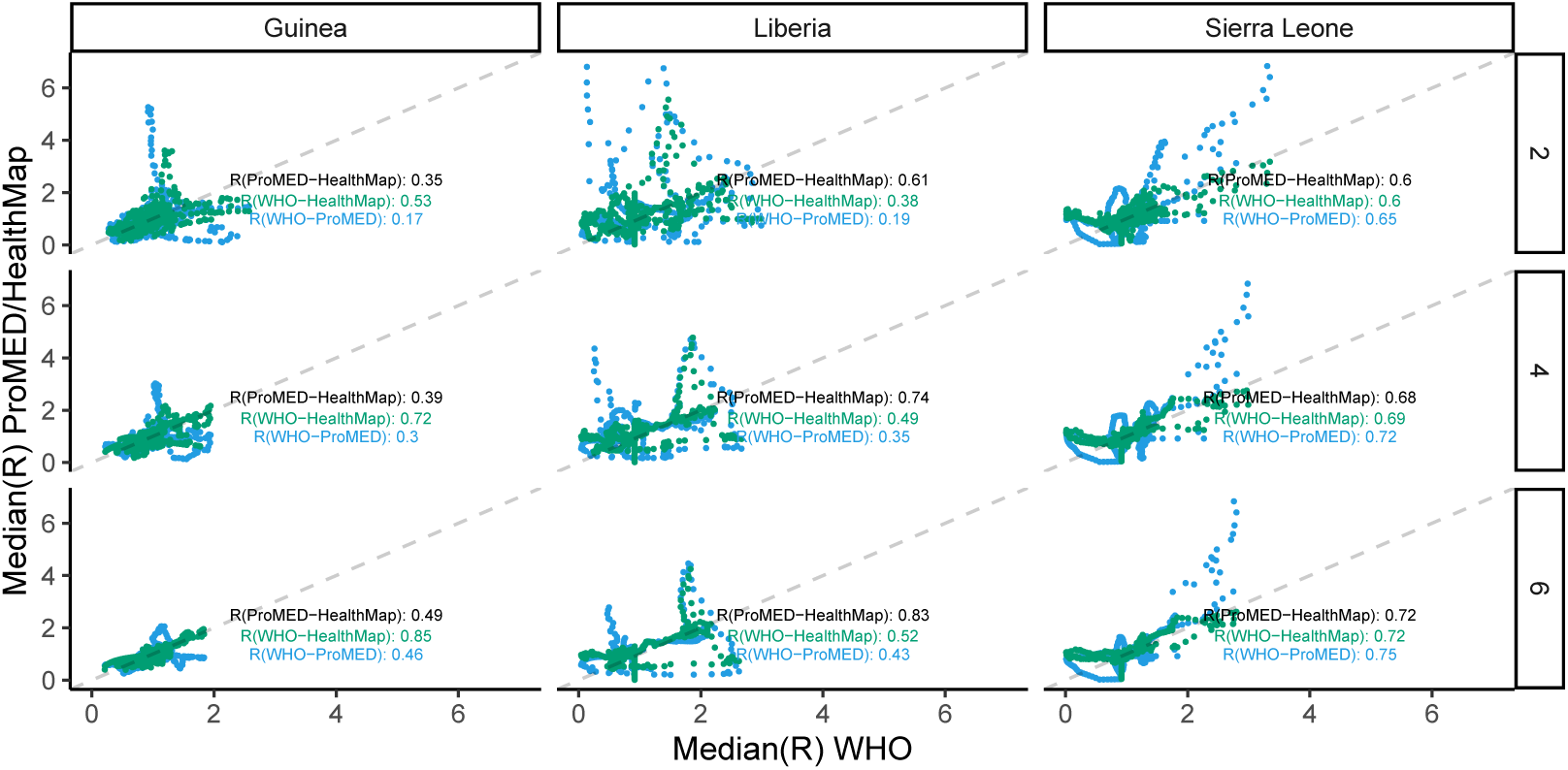
Correlation between time-varying reproduction number estimated from ProMED, HealthMap and WHO data. The reproduction numbers were estimated using R package EpiEstim over a 4 week sliding window. Median estimates from WHO data are on the x-axis and the median estimates using ProMED (blue) and HealthMap (green) data are on the y-axis. All correlation coefficients were statistically significant.

**Fig SI 4:**
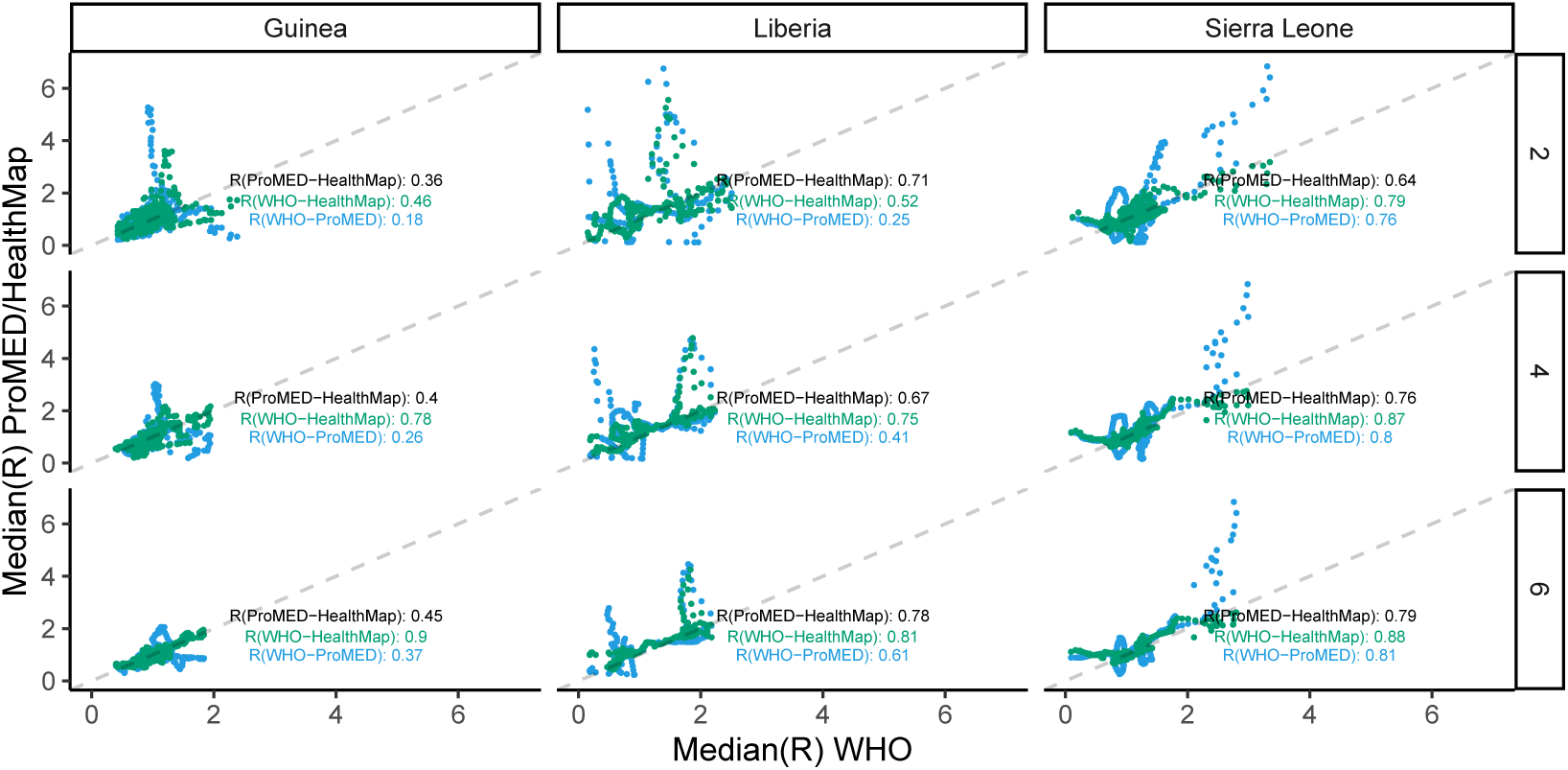
Correlation between time-varying reproduction number estimated from ProMED, HealthMap and WHO data. Only estimates with a coefficient of variation less than 0.25 were included in this analysis. The reproduction numbers were estimated using R package EpiEstim over a 4 week sliding window. Median estimates from WHO data are on the x-axis and the median estimates using ProMED (blue) and HealthMap (green) data are on the y-axis. All correlation coefficients were statistically significant.

**Fig SI 5:**
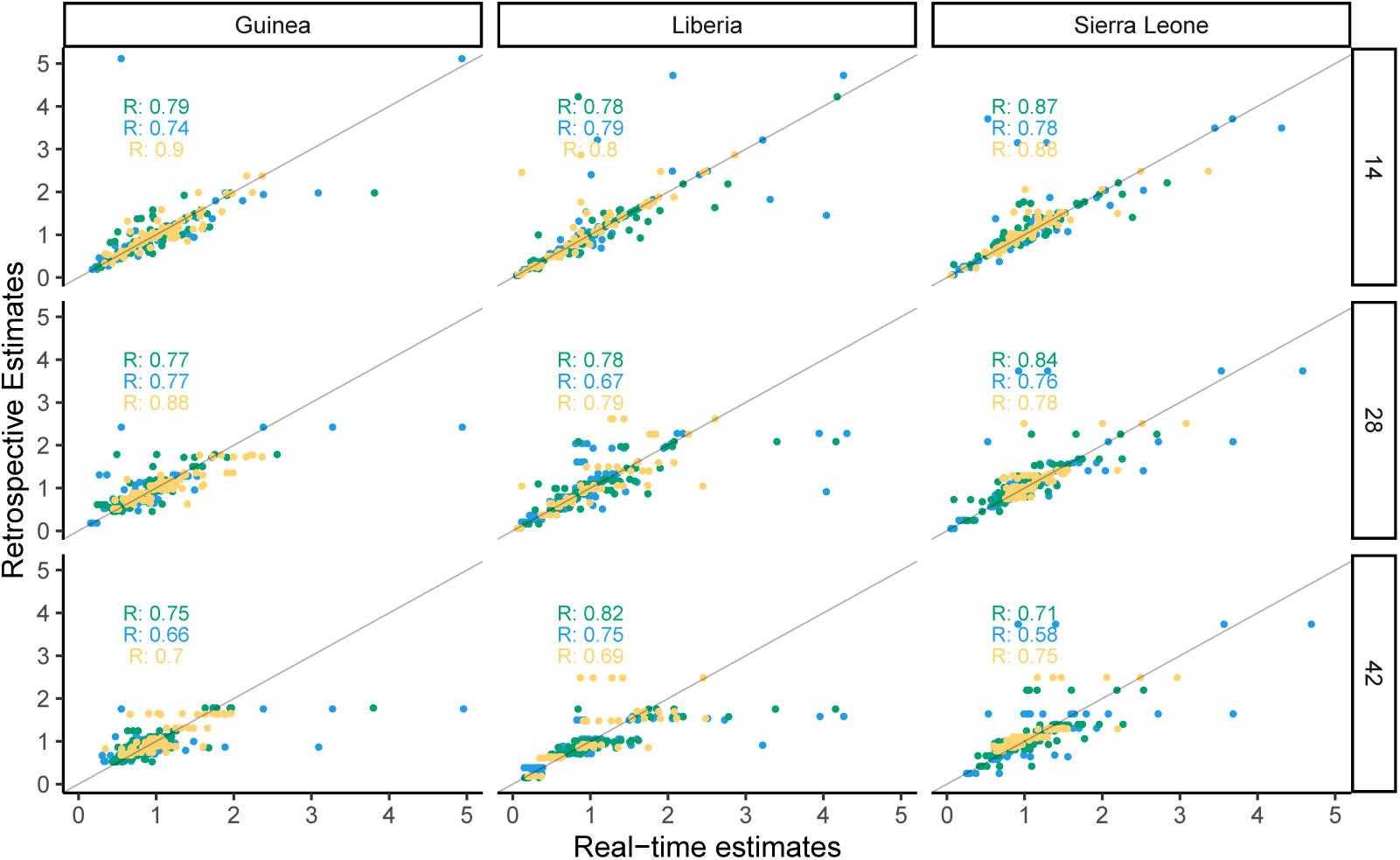
Correlation between real-time and retrospective estimates of time-varying reproduction number from ProMED (blue), HealthMap (green) and WHO (yellow) data. Median real-time estimates are on the x-axis and the median retrospective estimates are on the y-axis. All correlation coefficients were statistically significant.

### 2 Forecasts using ProMED data

This section presents the forecast produced using ProMED data with calibration window of 2 (Section 2.1), 4 (Section 2.2) and 6 (Section 2.3) weeks over a forecast horizon of 4, 6 and 8 weeks. Results using calibration window of 2 weeks and forecast horizon of 4 weeks are presented in the main text.

#### 2.1 Calibration window of 2 weeks

##### 2.1.1 Forecast horizon of 4 weeks

##### 2.1.2 Forecast horizon of 6 weeks

**Fig SI 6:**
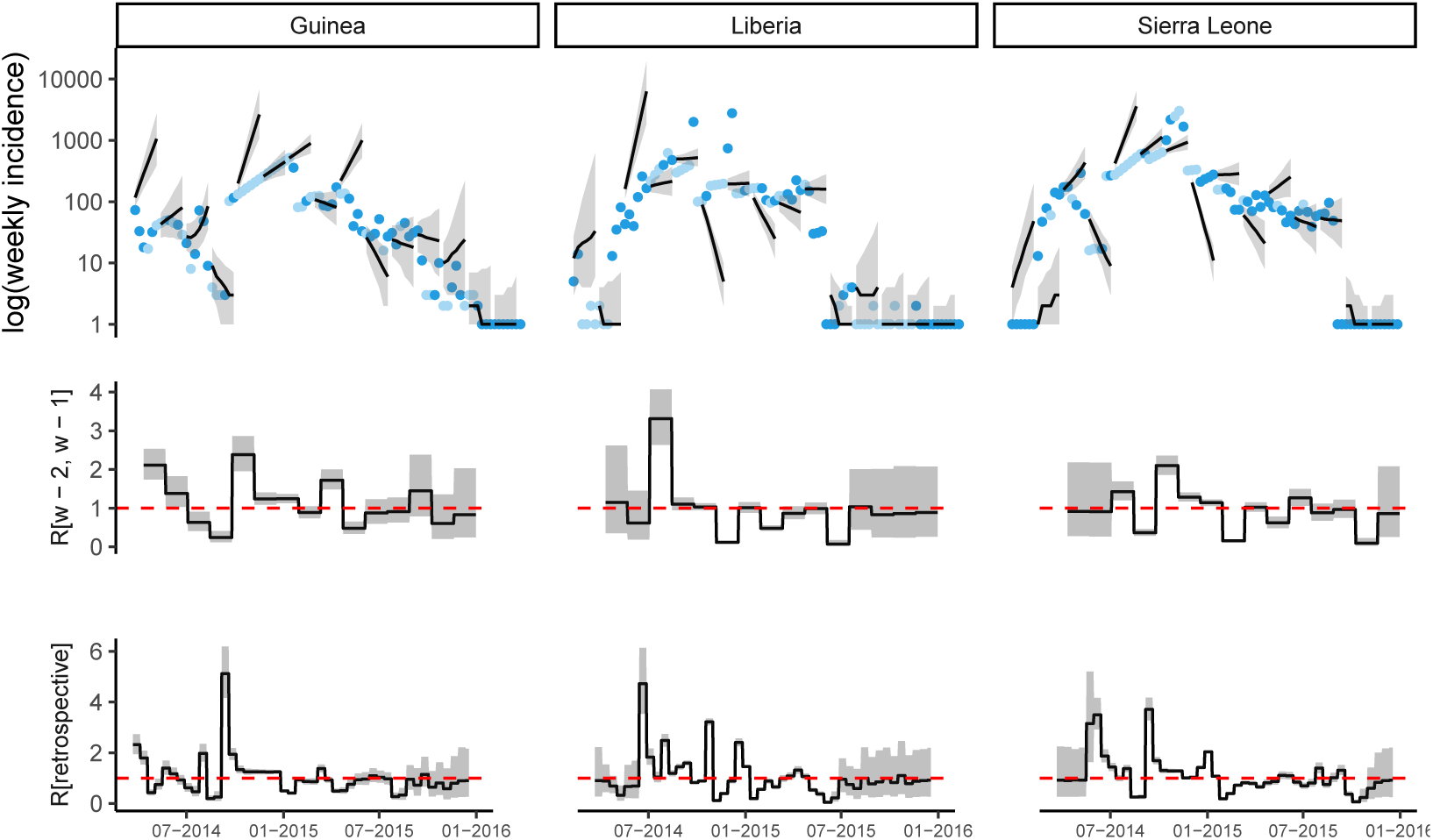
Observed and predicted incidence, and reproduction number estimates from ProMED data. The top panel shows the weekly incidence derived from ProMED data and the 6 weeks incidence forecast on log scale. The solid dots represent the observed weekly incidence where the light blue dots show weeks for which all data points were obtained using interpolation. The projections are made over 6 week windows, based on the reproduction number estimated in the previous 2 weeks. The middle figure in each panel shows the reproduction number used to make forecasts over each 6 week forecast horizon. The bottom figure shows the effective reproduction number estimated retrospectively using the full dataset up to the length of one calibration window before the end. In each case, the solid black line is the median estimate and the shaded region represents the 95% Credible Interval. The red horizontal dashed line indicates the *R*_*t*_ = 1 threshold. Results are shown for the three mainly affected countries although the analysis was done jointly using data for all countries in Africa.

**Table 1:** Percentage of weeks with observed incidence in 95% forecast interval. In forecasting ahead, we assumed transmissibility to be constant over the forecast horizon. If the 97.5th percentile of the R estimate used for forecasting was less than 1, we defined the epidemic to be in the declining phase during this period. Similarly, if the 2.5th percentile of R was greater than 1, we defined the epidemic to be in a growing phase. The phase was set to stable where the 95% Credible Interval of the R estimates contained 1.

| Country | Growing | Declining | Stable | Overall |
| --- | --- | --- | --- | --- |
| Guinea | 44.0% | 42.2% | 71.9% | 54.4% |
| Liberia | 18.3% | 31.5% | 69.5% | 49.3% |
| Sierra Leone | 25.9% | 46.0% | 55.7% | 42.5% |
| All | 30.8% | 40.2% | 66.7% | 48.7% |

##### 2.1.3 Forecast horizon of 8 weeks

**Fig SI 7:**
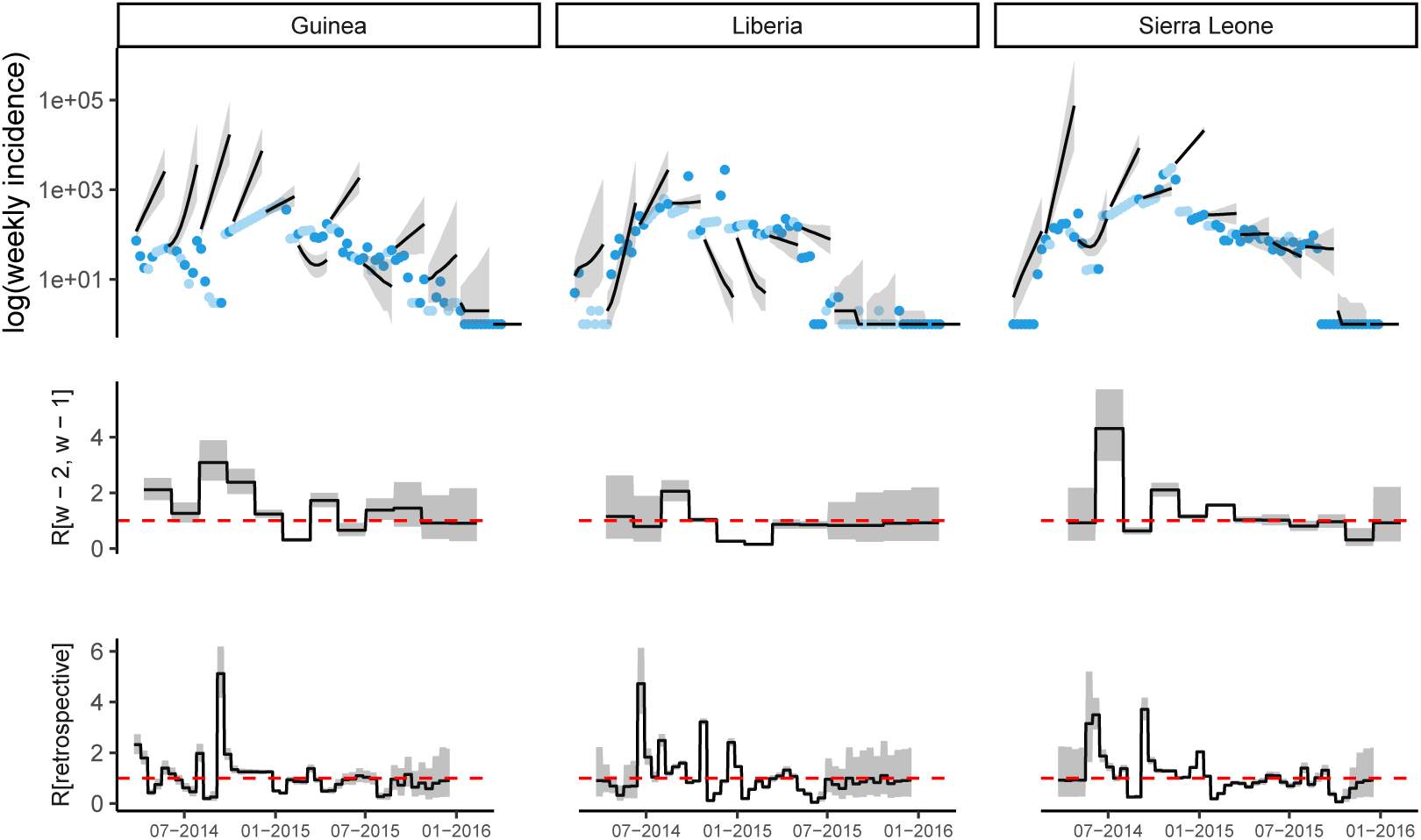
Observed and predicted incidence, and reproduction number estimates from ProMED data. The top panel shows the weekly incidence derived from ProMED data and the 8 weeks incidence forecast on log scale. The solid dots represent the observed weekly incidence where the light blue dots show weeks for which all data points were obtained using interpolation. The projections are made over 8 week windows, based on the reproduction number estimated in the previous 2 weeks. The middle figure in each panel shows the reproduction number used to make forecasts over each 8 week forecast horizon. The bottom figure shows the effective reproduction number estimated retrospectively using the full dataset up to the length of one calibration window before the end. In each case, the solid black line is the median estimate and the shaded region represents the 95% Credible Interval. The red horizontal dashed line indicates the *R*_*t*_ = 1 threshold. Results are shown for the three mainly affected countries although the analysis was done jointly using data for all countries in Africa.

#### 2.2 Calibration window of 4 weeks

##### 2.2.1 Forecast horizon of 4 weeks

**Fig SI 8:**
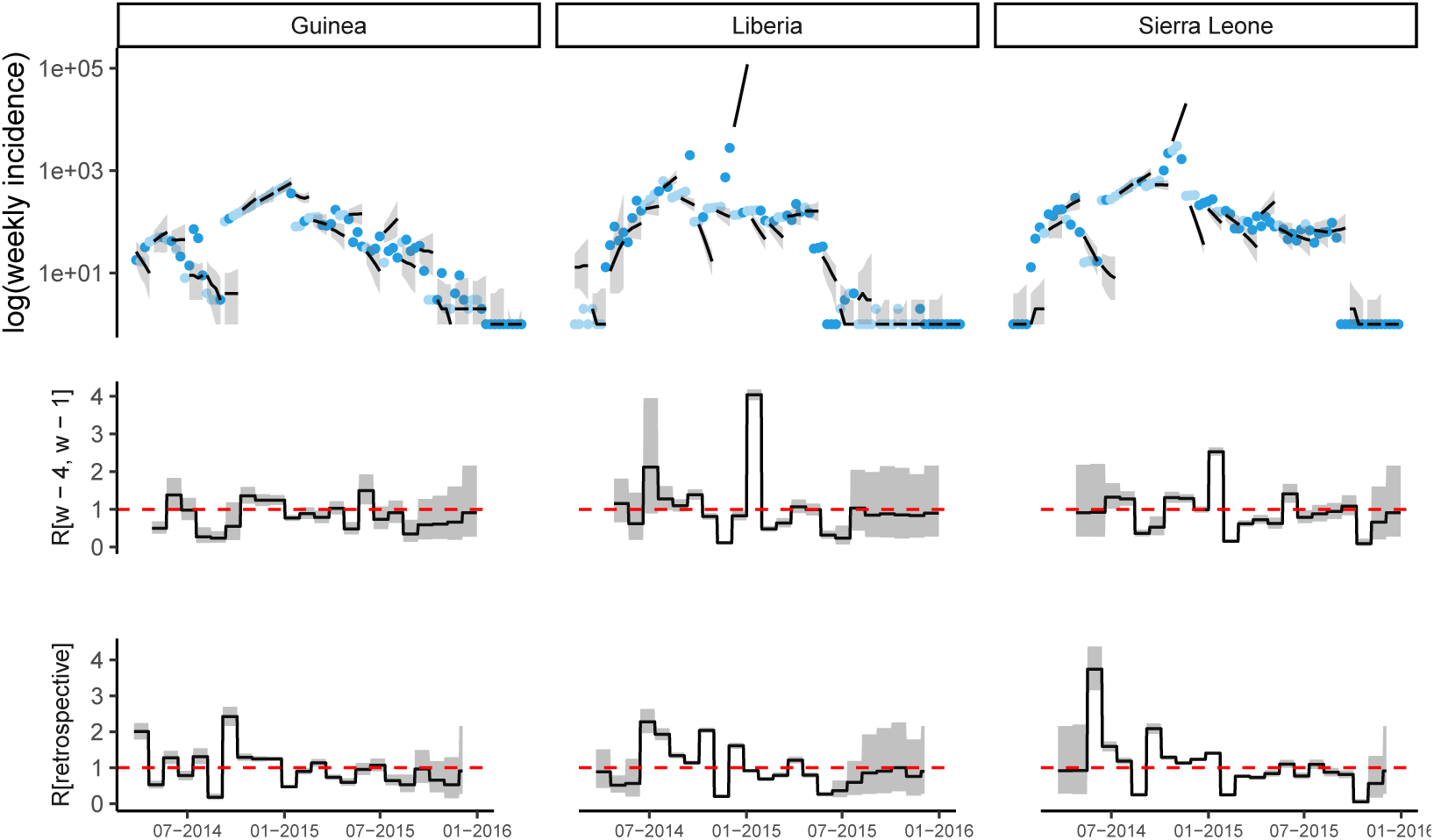
Observed and predicted incidence, and reproduction number estimates from ProMED data. The top panel shows the weekly incidence derived from ProMED data and the 4 weeks incidence forecast on log scale. The solid dots represent the observed weekly incidence where the light blue dots show weeks for which all data points were obtained using interpolation. The projections are made over 4 week windows, based on the reproduction number estimated in the previous 4 weeks. The middle figure in each panel shows the reproduction number used to make forecasts over each 4 week forecast horizon. The bottom figure shows the effective reproduction number estimated retrospectively using the full dataset up to the length of one calibration window before the end. In each case, the solid black line is the median estimate and the shaded region represents the 95% Credible Interval. The red horizontal dashed line indicates the *R*_*t*_ = 1 threshold. Results are shown for the three mainly affected countries although the analysis was done jointly using data for all countries in Africa.

##### 2.2.2 Forecast horizon of 6 weeks

**Fig SI 9:**
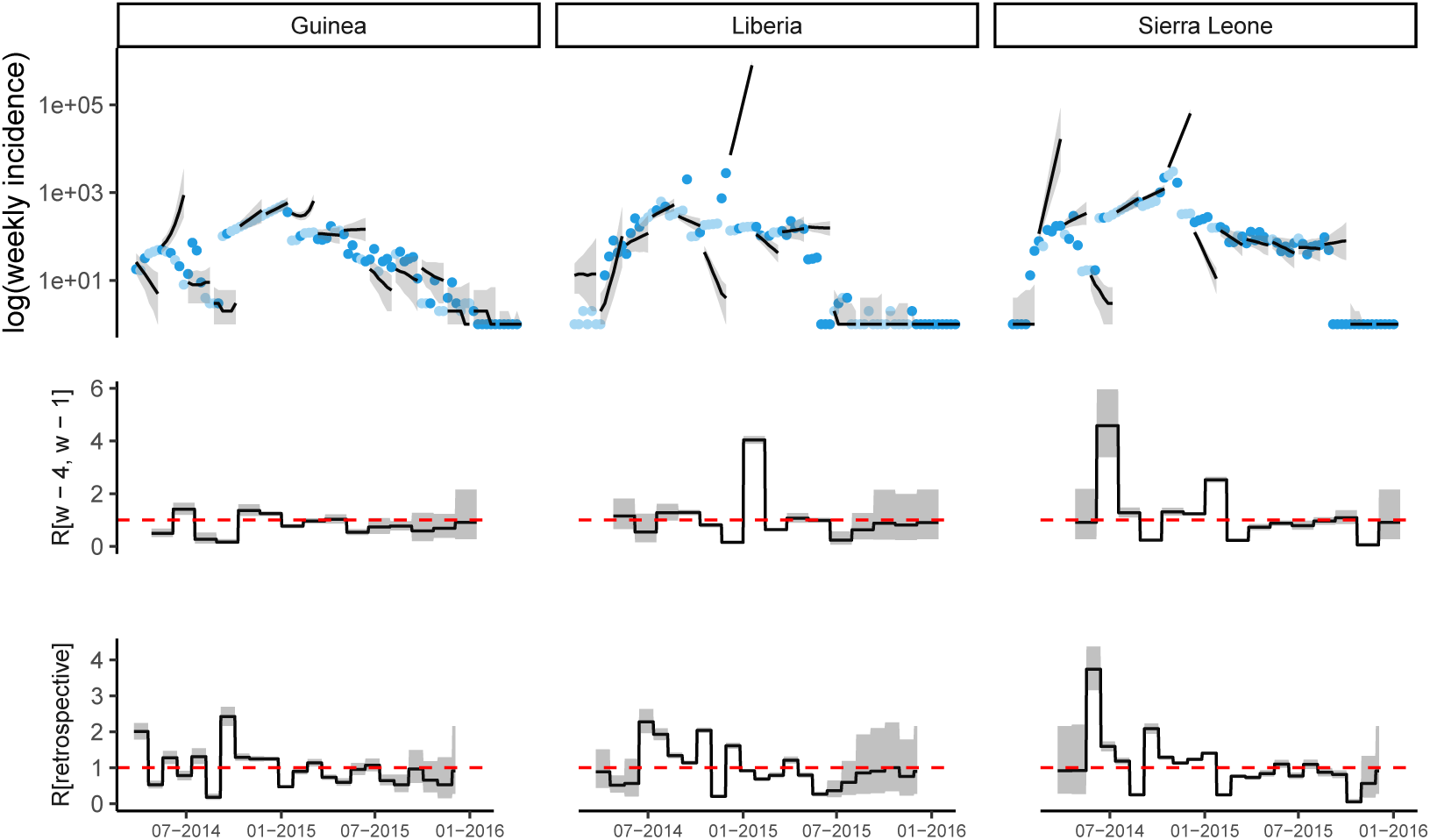
Observed and predicted incidence, and reproduction number estimates from ProMED data. The top panel shows the weekly incidence derived from ProMED data and the 6 weeks incidence forecast on log scale. The solid dots represent the observed weekly incidence where the light blue dots show weeks for which all data points were obtained using interpolation. The projections are made over 6 week windows, based on the reproduction number estimated in the previous 4 weeks. The middle figure in each panel shows the reproduction number used to make forecasts over each 6 week forecast horizon. The bottom figure shows the effective reproduction number estimated retrospectively using the full dataset up to the length of one calibration window before the end. In each case, the solid black line is the median estimate and the shaded region represents the 95% Credible Interval. The red horizontal dashed line indicates the *R*_*t*_ = 1 threshold. Results are shown for the three mainly affected countries although the analysis was done jointly using data for all countries in Africa.

##### 2.2.3 Forecast horizon of 8 weeks

**Fig SI 10:**
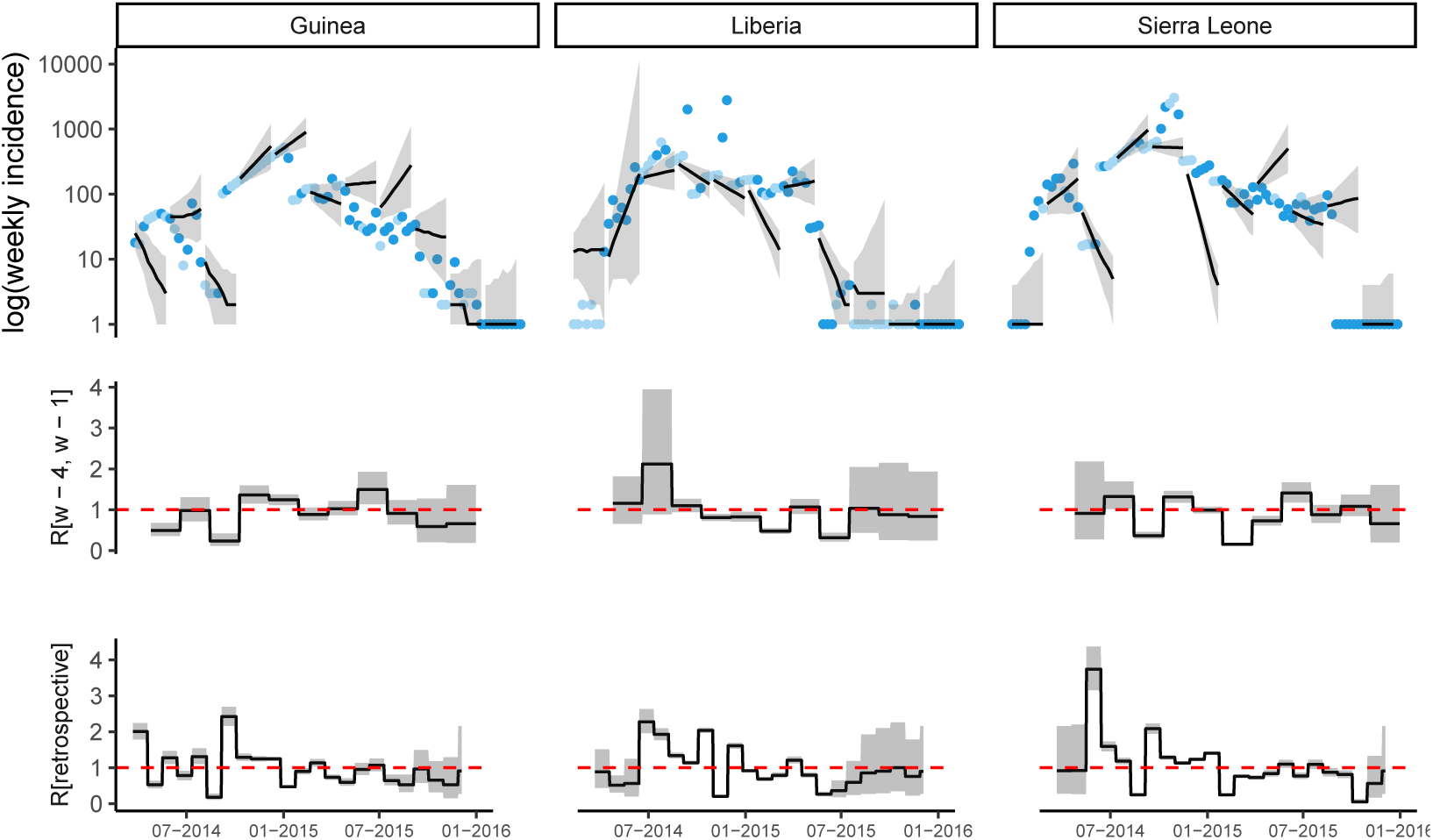
Observed and predicted incidence, and reproduction number estimates from ProMED data. The top panel shows the weekly incidence derived from ProMED data and the 8 weeks incidence forecast on log scale. The solid dots represent the observed weekly incidence where the light blue dots show weeks for which all data points were obtained using interpolation. The projections are made over 8 week windows, based on the reproduction number estimated in the previous 4 weeks. The middle figure in each panel shows the reproduction number used to make forecasts over each 8 week forecast horizon. The bottom figure shows the effective reproduction number estimated retrospectively using the full dataset up to the length of one calibration window before the end. In each case, the solid black line is the median estimate and the shaded region represents the 95% Credible Interval. The red horizontal dashed line indicates the *R*_*t*_ = 1 threshold. Results are shown for the three mainly affected countries although the analysis was done jointly using data for all countries in Africa.

#### 2.3 Calibration window of 6 weeks

##### 2.3.1 Forecast horizon of 4 weeks

**Fig SI 11:**
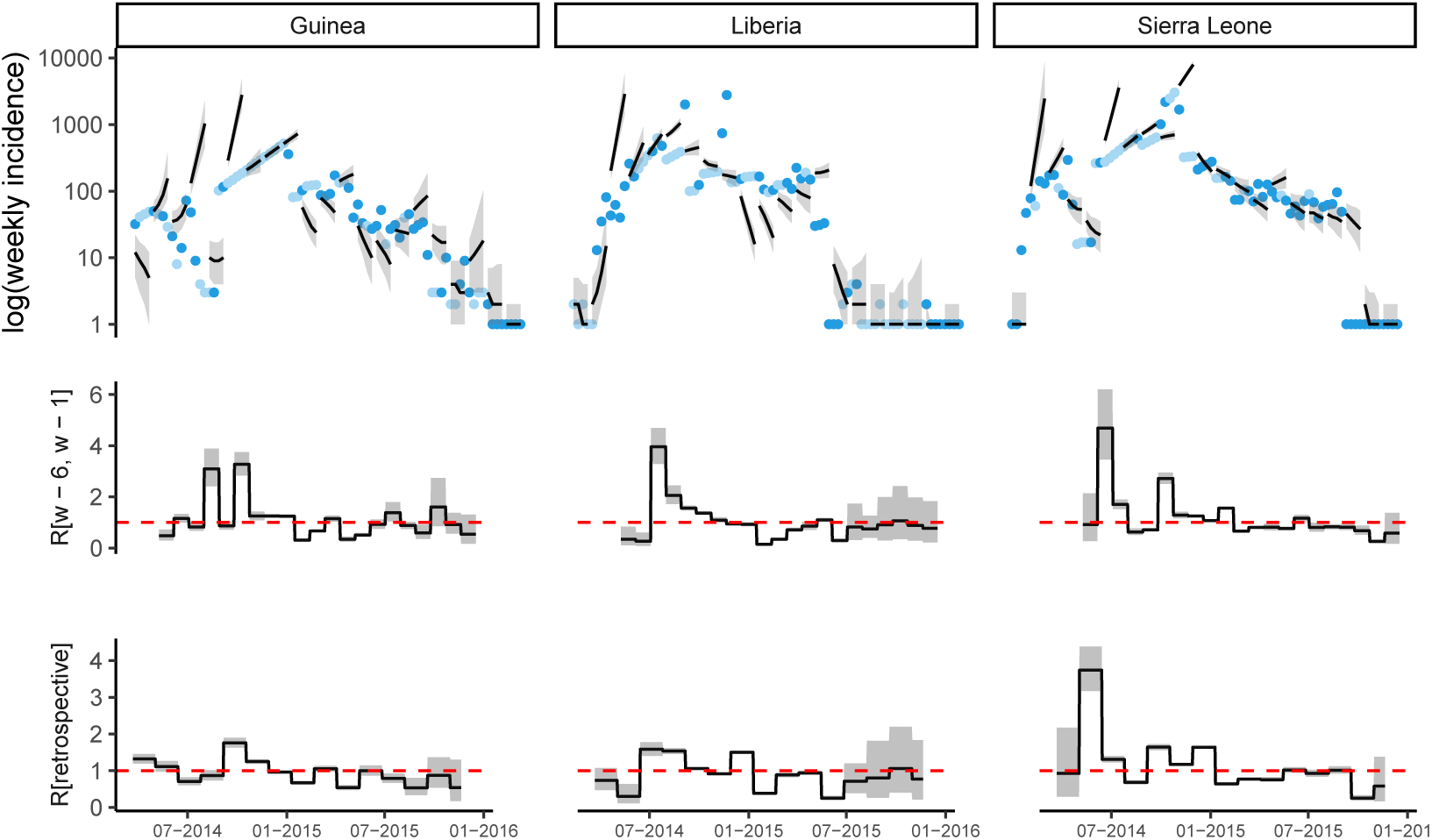
Observed and predicted incidence, and reproduction number estimates from ProMED data. The top panel shows the weekly incidence derived from ProMED data and the 4 weeks incidence forecast on log scale. The solid dots represent the observed weekly incidence where the light blue dots show weeks for which all data points were obtained using interpolation. The projections are made over 4 week windows, based on the reproduction number estimated in the previous 6 weeks. The middle figure in each panel shows the reproduction number used to make forecasts over each 4 week forecast horizon. The bottom figure shows the effective reproduction number estimated retrospectively using the full dataset up to the length of one calibration window before the end. In each case, the solid black line is the median estimate and the shaded region represents the 95% Credible Interval. The red horizontal dashed line indicates the *R*_*t*_ = 1 threshold. Results are shown for the three mainly affected countries although the analysis was done jointly using data for all countries in Africa.

##### 2.3.2 Forecast horizon of 6 weeks

**Fig SI 12:**
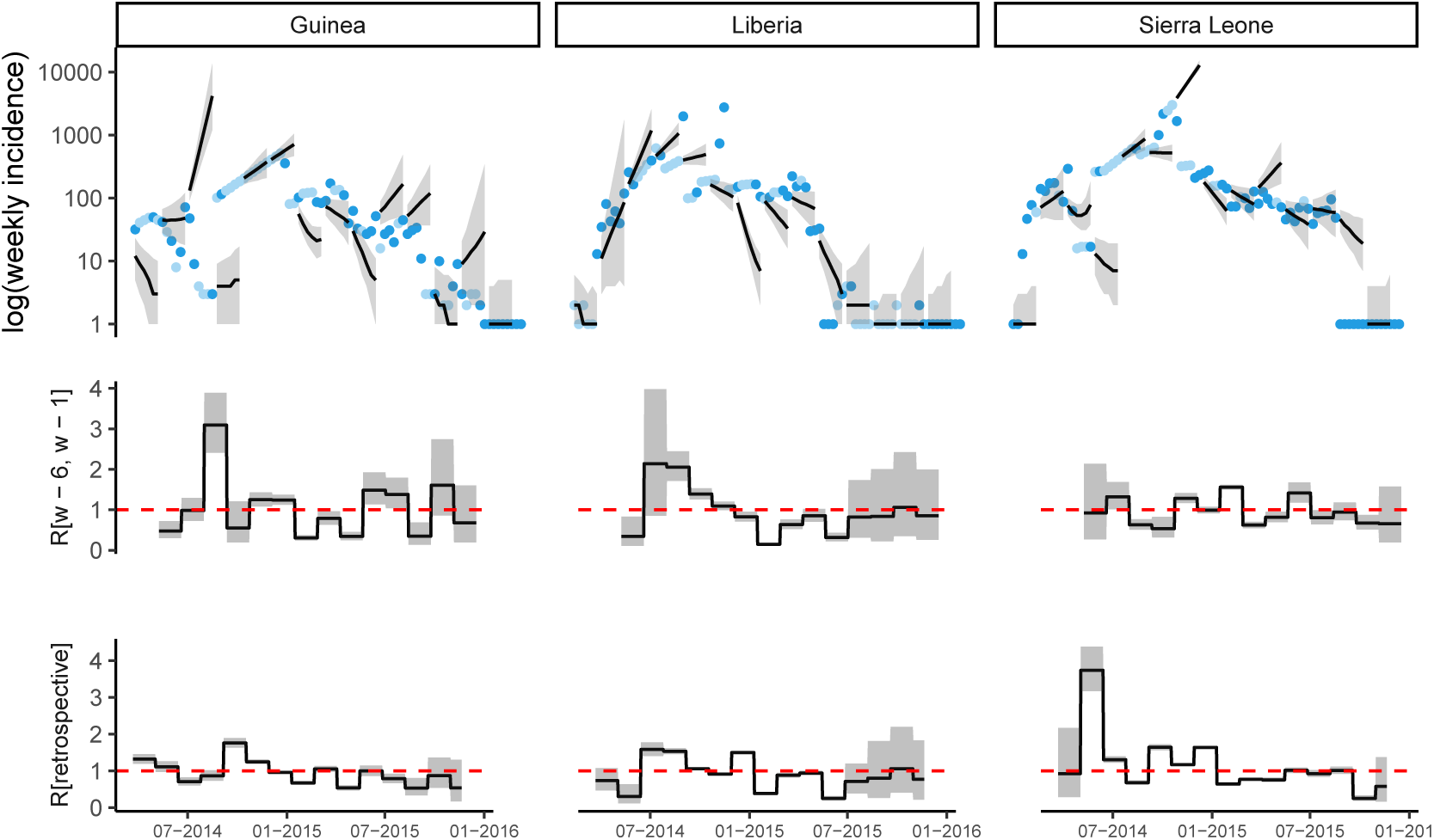
Observed and predicted incidence, and reproduction number estimates from ProMED data. The top panel shows the weekly incidence derived from ProMED data and the 6 weeks incidence forecast on log scale. The solid dots represent the observed weekly incidence where the light blue dots show weeks for which all data points were obtained using interpolation. The projections are made over 6 week windows, based on the reproduction number estimated in the previous 6 weeks. The middle figure in each panel shows the reproduction number used to make forecasts over each 6 week forecast horizon. The bottom figure shows the effective reproduction number estimated retrospectively using the full dataset up to the length of one calibration window before the end. In each case, the solid black line is the median estimate and the shaded region represents the 95% Credible Interval. The red horizontal dashed line indicates the *R*_*t*_ = 1 threshold. Results are shown for the three mainly affected countries although the analysis was done jointly using data for all countries in Africa.

##### 2.3.3 Forecast horizon of 8 weeks

**Fig SI 13:**
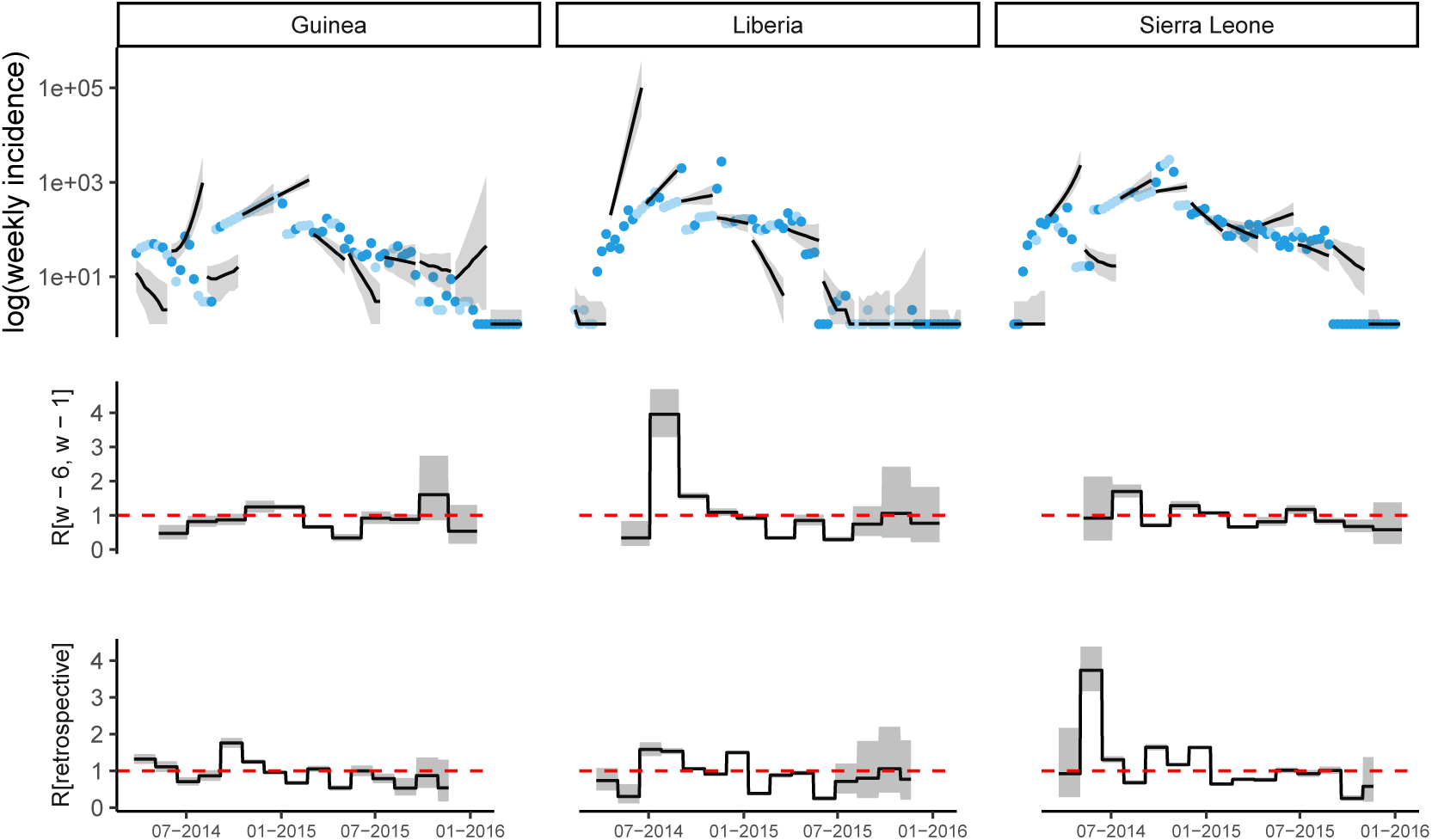
Observed and predicted incidence, and reproduction number estimates from ProMED data. The top panel shows the weekly incidence derived from ProMED data and the 8 weeks incidence forecast on log scale. The solid dots represent the observed weekly incidence where the light blue dots show weeks for which all data points were obtained using interpolation. The projections are made over 8 week windows, based on the reproduction number estimated in the previous 6 weeks. The middle figure in each panel shows the reproduction number used to make forecasts over each 8 week forecast horizon. The bottom figure shows the effective reproduction number estimated retrospectively using the full dataset up to the length of one calibration window before the end. In each case, the solid black line is the median estimate and the shaded region represents the 95% Credible Interval. The red horizontal dashed line indicates the *R*_*t*_ = 1 threshold. Results are shown for the three mainly affected countries although the analysis was done jointly using data for all countries in Africa.

### 3 Forecasts using HealthMap data

This section presents the forecasts over 4, 6 and 8 weeks produced using HealthMap data and calibration window of 2, 4 and 6 weeks.

#### 3.1 Calibration window of 2 weeks

##### 3.1.1 Forecast horizon of 4 weeks

**Fig SI 14:**
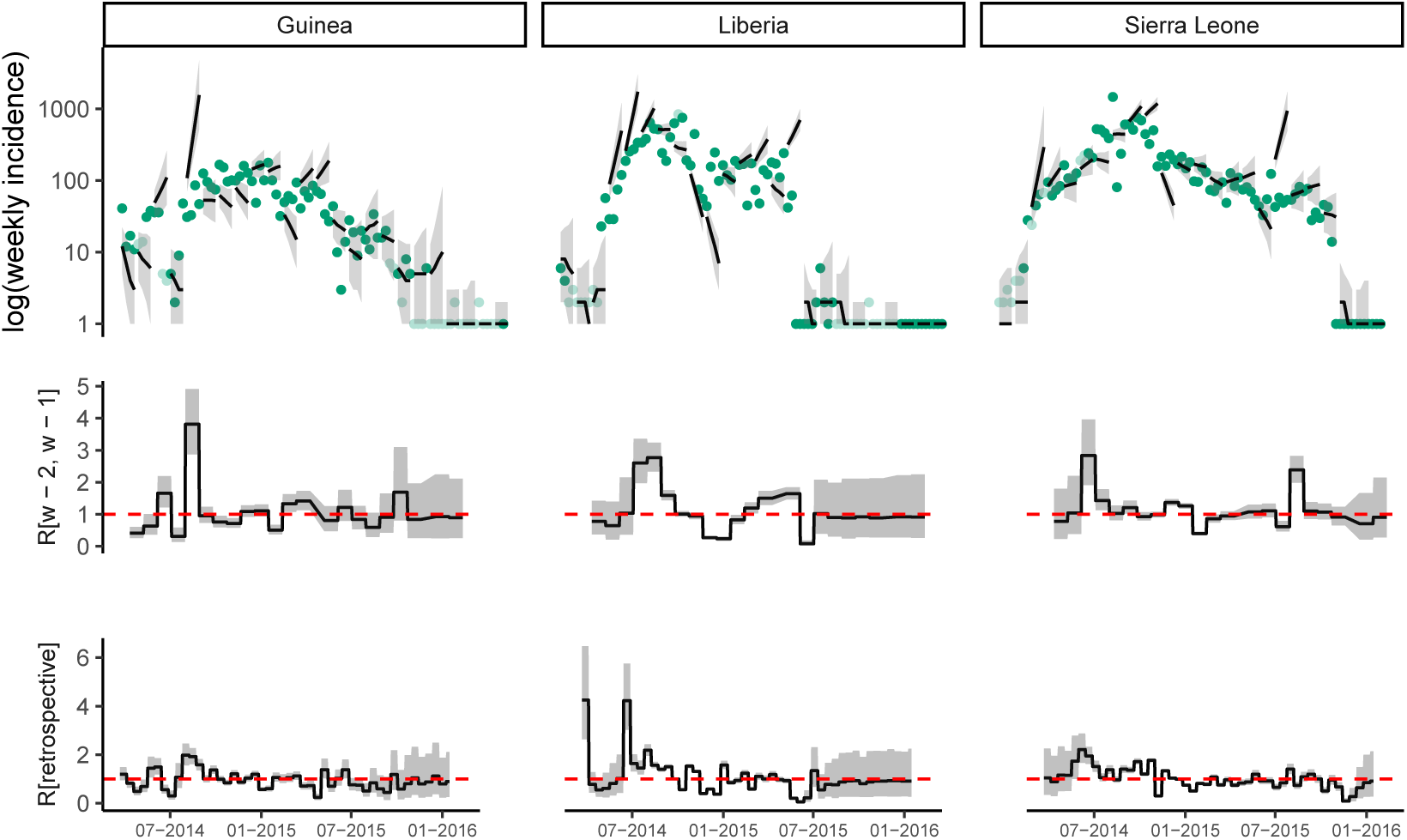
Observed and predicted incidence, and reproduction number estimates from HealthMap data. The top panel shows the weekly incidence derived from HealthMap data and the 4 weeks incidence forecast on log scale. The solid dots represent the observed weekly incidence where the light green dots show weeks for which all data points were obtained using interpolation. The projections are made over 4 week windows, based on the reproduction number estimated in the previous 2 weeks. The middle figure in each panel shows the reproduction number used to make forecasts over each 4 week forecast horizon. The bottom figure shows the effective reproduction number estimated retrospectively using the full dataset up to the length of one calibration window before the end. In each case, the solid black line is the median estimate and the shaded region represents the 95% Credible Interval. The red horizontal dashed line indicates the *R*_*t*_ = 1 threshold. Results are shown for the three mainly affected countries although the analysis was done jointly using data for all countries in Africa.

##### 3.1.2 Forecast horizon of 6 weeks

**Fig SI 15:**
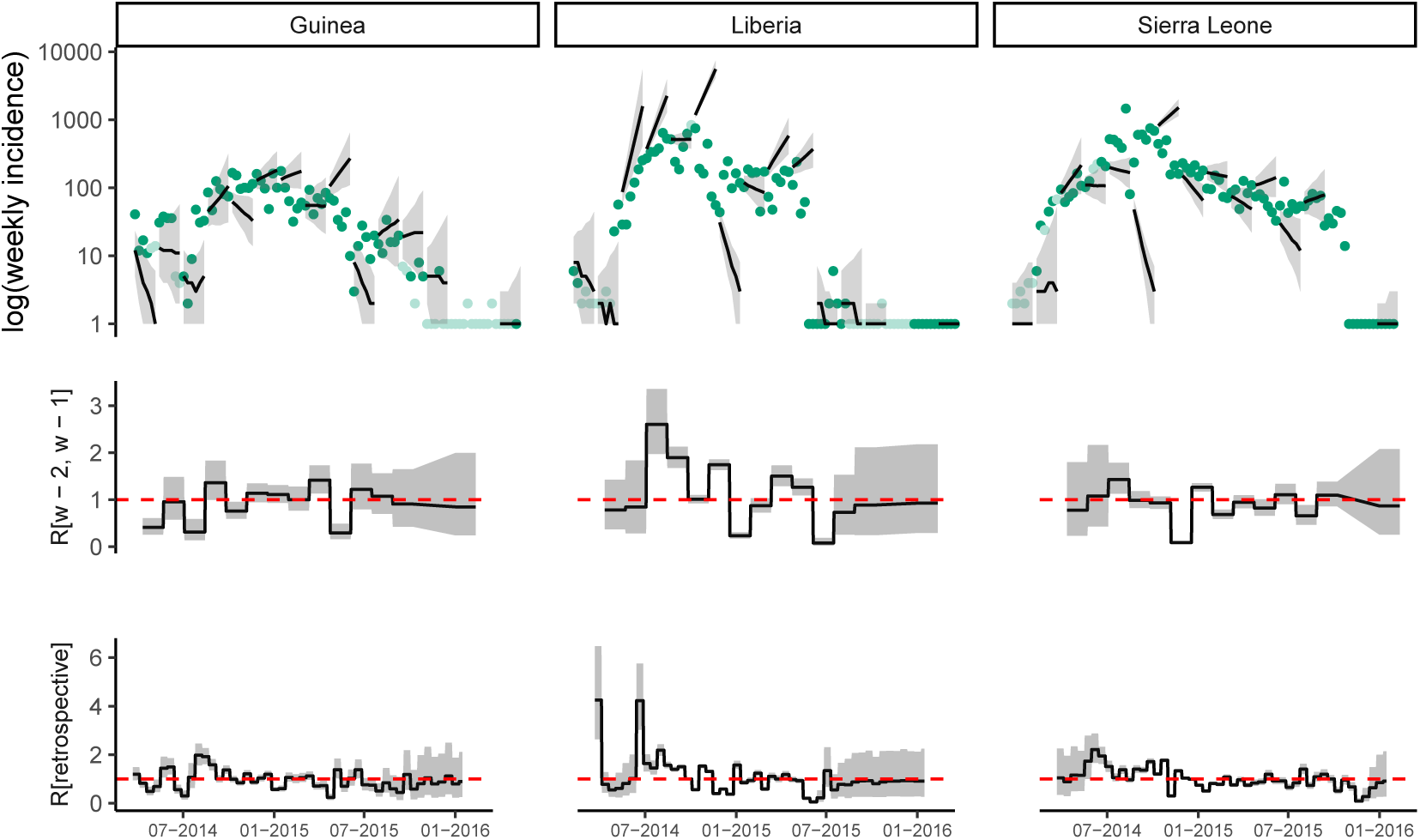
Observed and predicted incidence, and reproduction number estimates from HealthMap data. The top panel shows the weekly incidence derived from HealthMap data and the 6 weeks incidence forecast on log scale. The solid dots represent the observed weekly incidence where the light green dots show weeks for which all data points were obtained using interpolation. The projections are made over 6 week windows, based on the reproduction number estimated in the previous 2 weeks. The middle figure in each panel shows the reproduction number used to make forecasts over each 6 week forecast horizon. The bottom figure shows the effective reproduction number estimated retrospectively using the full dataset up to the length of one calibration window before the end. In each case, the solid black line is the median estimate and the shaded region represents the 95% Credible Interval. The red horizontal dashed line indicates the *R*_*t*_ = 1 threshold. Results are shown for the three mainly affected countries although the analysis was done jointly using data for all countries in Africa.

##### 3.1.3 Forecast horizon of 8 weeks

**Fig SI 16:**
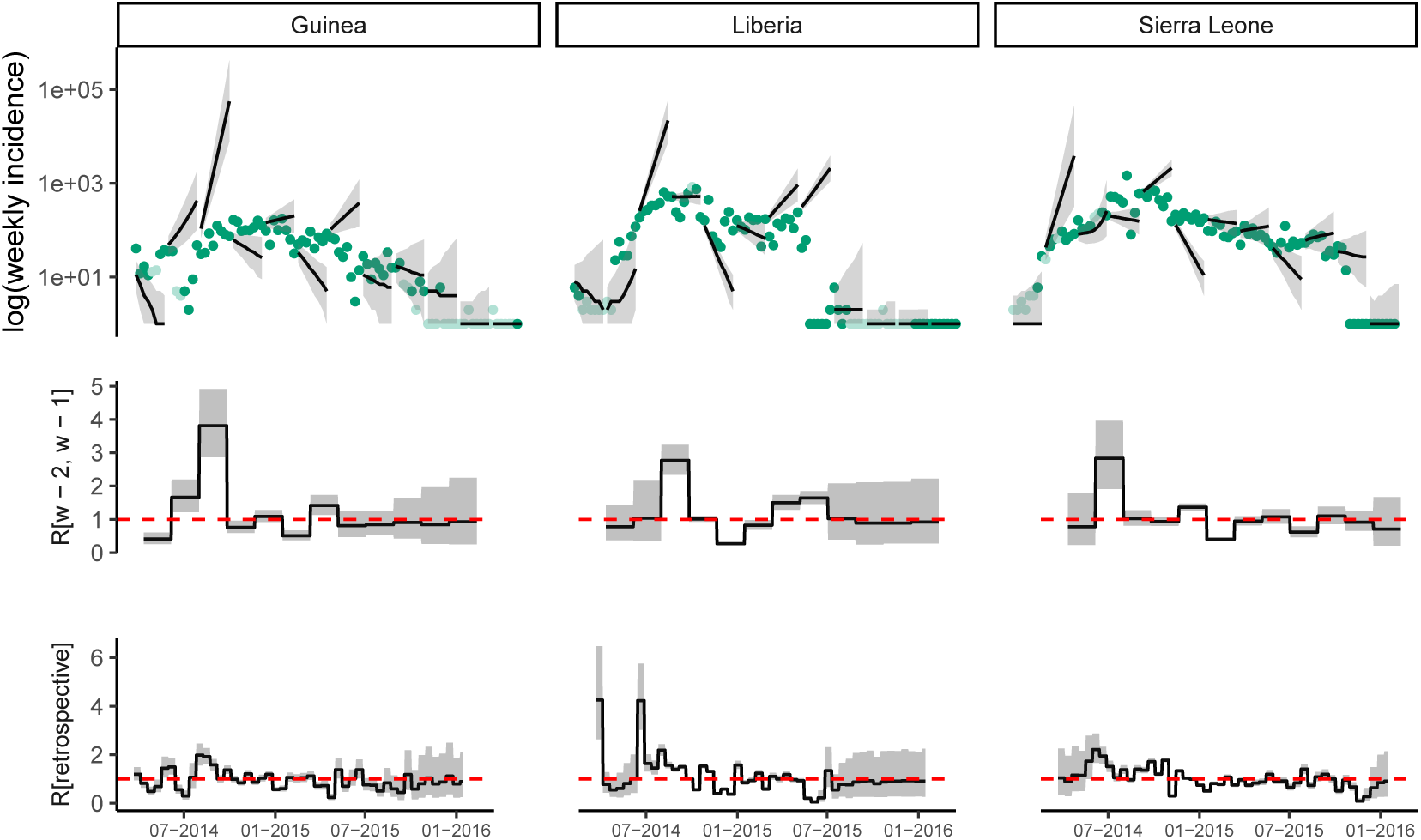
Observed and predicted incidence, and reproduction number estimates from HealthMap data. The top panel shows the weekly incidence derived from HealthMap data and the 8 weeks incidence forecast on log scale. The solid dots represent the observed weekly incidence where the light green dots show weeks for which all data points were obtained using interpolation. The projections are made over 8 week windows, based on the reproduction number estimated in the previous 2 weeks. The middle figure in each panel shows the reproduction number used to make forecasts over each 8 week forecast horizon. The bottom figure shows the effective reproduction number estimated retrospectively using the full dataset up to the length of one calibration window before the end. In each case, the solid black line is the median estimate and the shaded region represents the 95% Credible Interval. The red horizontal dashed line indicates the *R*_*t*_ = 1 threshold. Results are shown for the three mainly affected countries although the analysis was done jointly using data for all countries in Africa.

#### 3.2 Calibration window of 4 weeks

##### 3.2.1 Forecast horizon of 4 weeks

**Fig SI 17:**
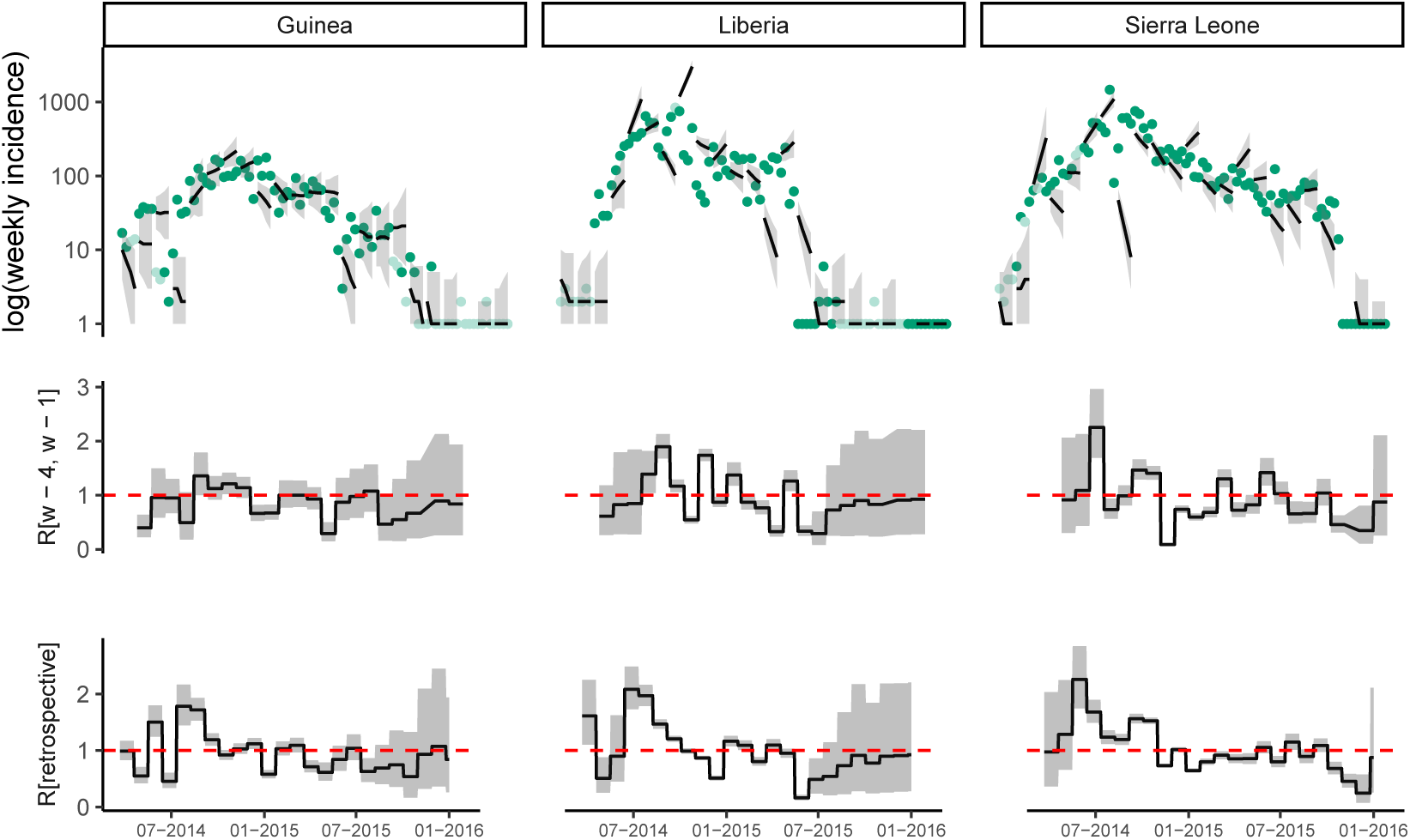
Observed and predicted incidence, and reproduction number estimates from HealthMap data. The top panel shows the weekly incidence derived from HealthMap data and the 4 weeks incidence forecast on log scale. The solid dots represent the observed weekly incidence where the light green dots show weeks for which all data points were obtained using interpolation. The projections are made over 4 week windows, based on the reproduction number estimated in the previous 4 weeks. The middle figure in each panel shows the reproduction number used to make forecasts over each 4 week forecast horizon. The bottom figure shows the effective reproduction number estimated retrospectively using the full dataset up to the length of one calibration window before the end. In each case, the solid black line is the median estimate and the shaded region represents the 95% Credible Interval. The red horizontal dashed line indicates the *R*_*t*_ = 1 threshold. Results are shown for the three mainly affected countries although the analysis was done jointly using data for all countries in Africa.

##### 3.2.2 Forecast horizon of 6 weeks

**Fig SI 18:**
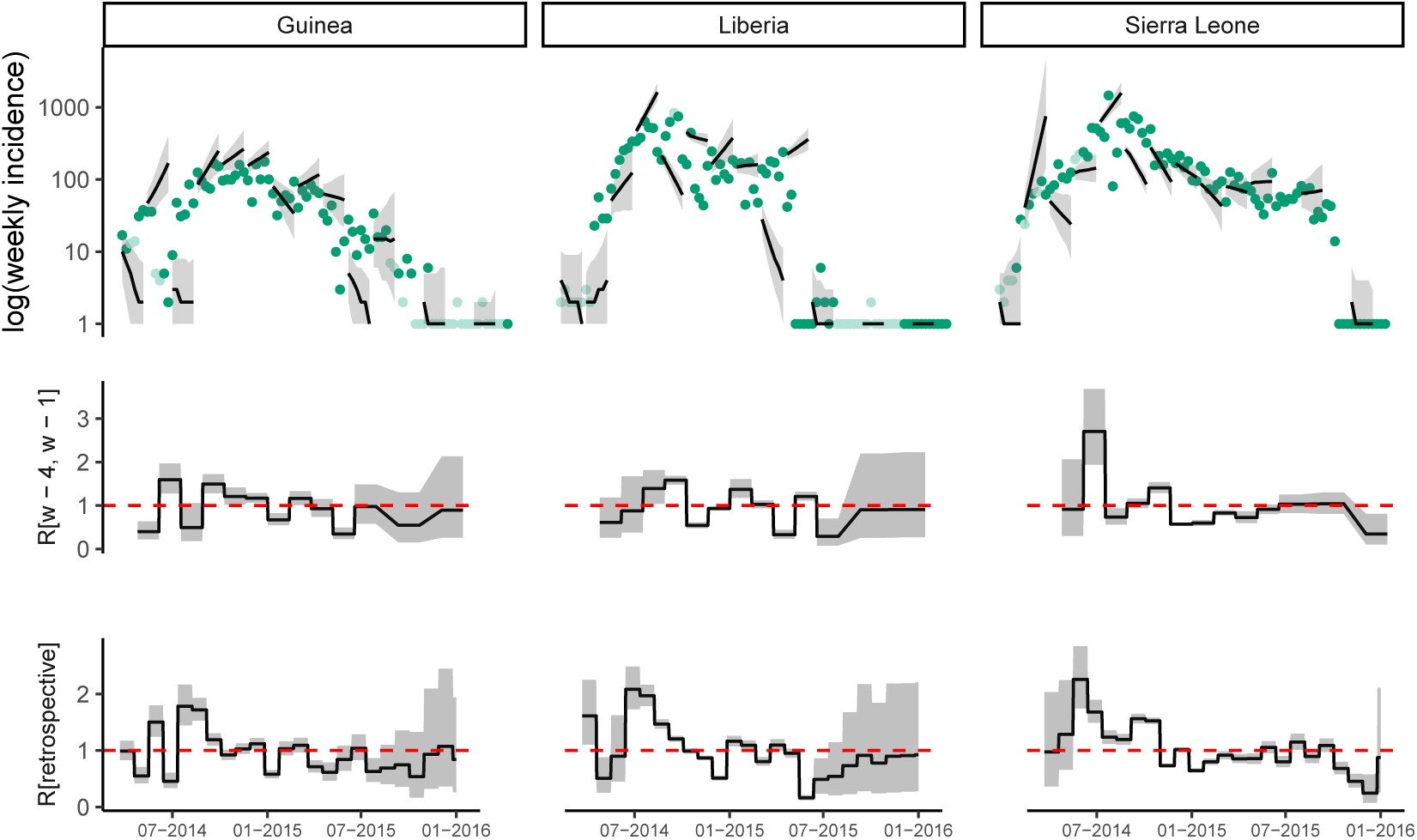
Observed and predicted incidence, and reproduction number estimates from HealthMap data. The top panel shows the weekly incidence derived from HealthMap data and the 6 weeks incidence forecast on log scale. The solid dots represent the observed weekly incidence where the light green dots show weeks for which all data points were obtained using interpolation. The projections are made over 6 week windows, based on the reproduction number estimated in the previous 4 weeks. The middle figure in each panel shows the reproduction number used to make forecasts over each 6 week forecast horizon. The bottom figure shows the effective reproduction number estimated retrospectively using the full dataset up to the length of one calibration window before the end. In each case, the solid black line is the median estimate and the shaded region represents the 95% Credible Interval. The red horizontal dashed line indicates the *R*_*t*_ = 1 threshold. Results are shown for the three mainly affected countries although the analysis was done jointly using data for all countries in Africa.

##### 3.2.3 Forecast horizon of 8 weeks

**Fig SI 19:**
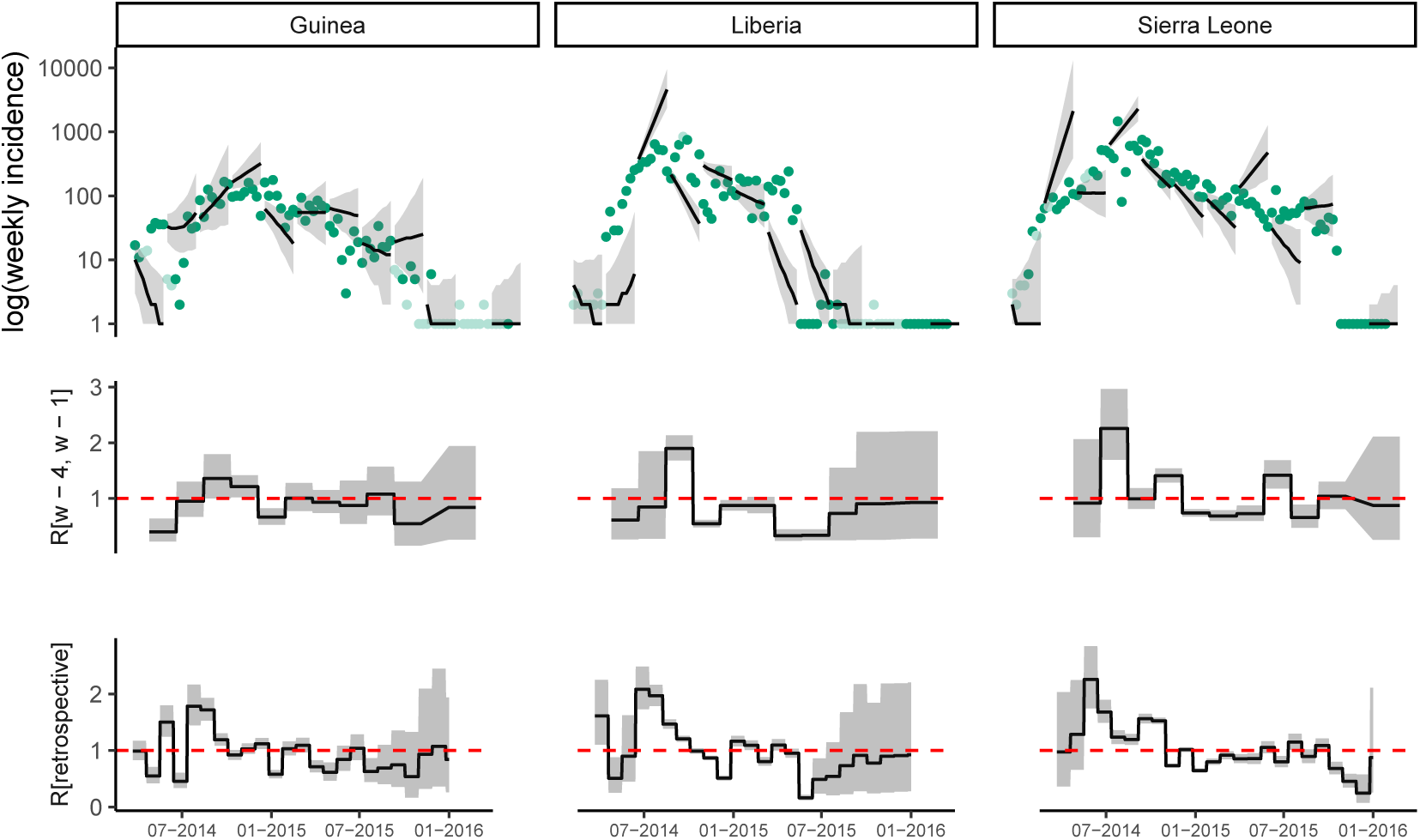
Observed and predicted incidence, and reproduction number estimates from HealthMap data. The top panel shows the weekly incidence derived from HealthMap data and the 8 weeks incidence forecast on log scale. The solid dots represent the observed weekly incidence where the light green dots show weeks for which all data points were obtained using interpolation. The projections are made over 8 week windows, based on the reproduction number estimated in the previous 4 weeks. The middle figure in each panel shows the reproduction number used to make forecasts over each 8 week forecast horizon. The bottom figure shows the effective reproduction number estimated retrospectively using the full dataset up to the length of one calibration window before the end. In each case, the solid black line is the median estimate and the shaded region represents the 95% Credible Interval. The red horizontal dashed line indicates the *R*_*t*_ = 1 threshold. Results are shown for the three mainly affected countries although the analysis was done jointly using data for all countries in Africa.

#### 3.3 Calibration window of 6 weeks

##### 3.3.1 Forecast horizon of 4 weeks

**Fig SI 20:**
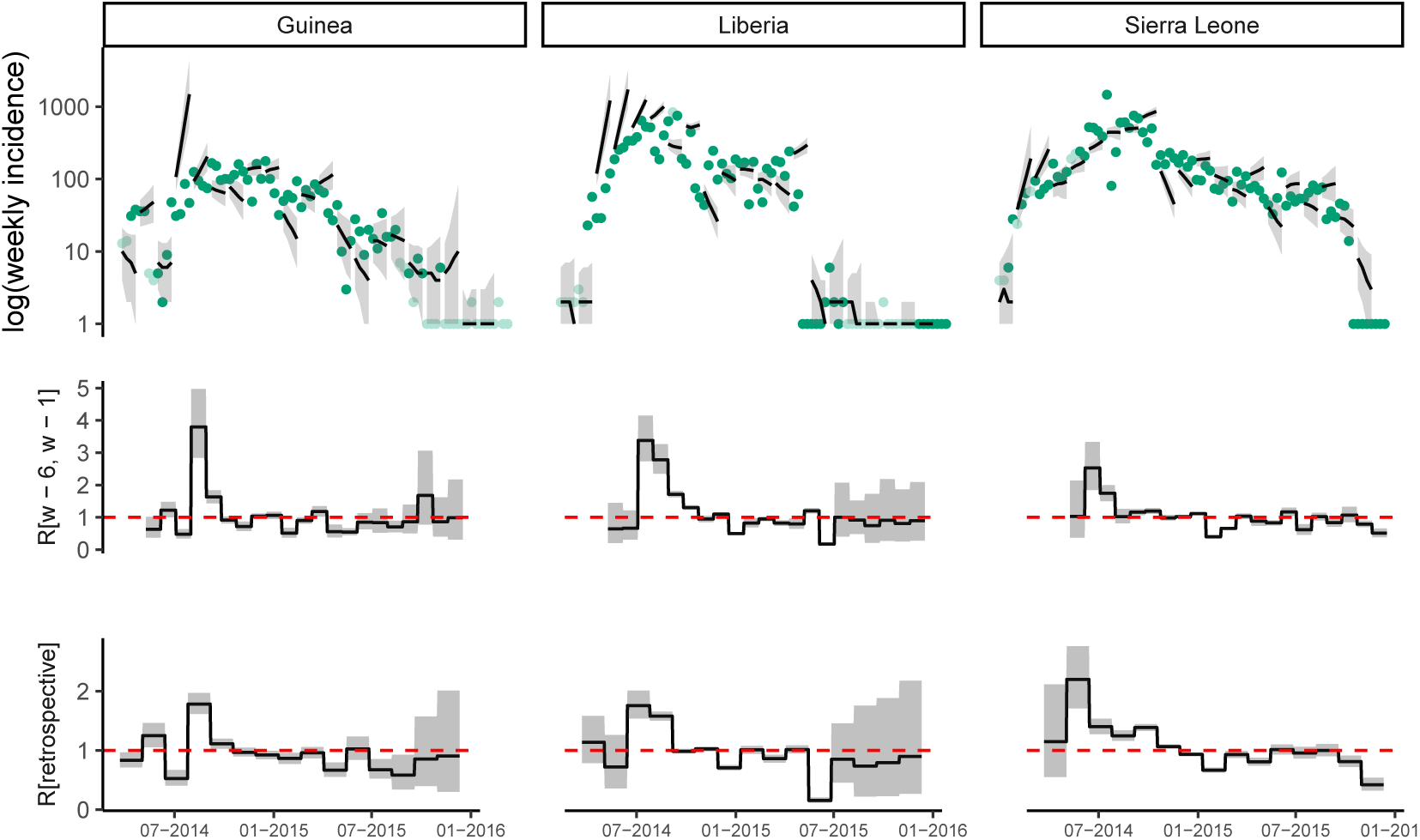
Observed and predicted incidence, and reproduction number estimates from HealthMap data. The top panel shows the weekly incidence derived from HealthMap data and the 4 weeks incidence forecast on log scale. The solid dots represent the observed weekly incidence where the light green dots show weeks for which all data points were obtained using interpolation. The projections are made over 4 week windows, based on the reproduction number estimated in the previous 6 weeks. The middle figure in each panel shows the reproduction number used to make forecasts over each 4 week forecast horizon. The bottom figure shows the effective reproduction number estimated retrospectively using the full dataset up to the length of one calibration window before the end. In each case, the solid black line is the median estimate and the shaded region represents the 95% Credible Interval. The red horizontal dashed line indicates the *R*_*t*_ = 1 threshold. Results are shown for the three mainly affected countries although the analysis was done jointly using data for all countries in Africa.

##### 3.3.2 Forecast horizon of 6 weeks

**Fig SI 21:**
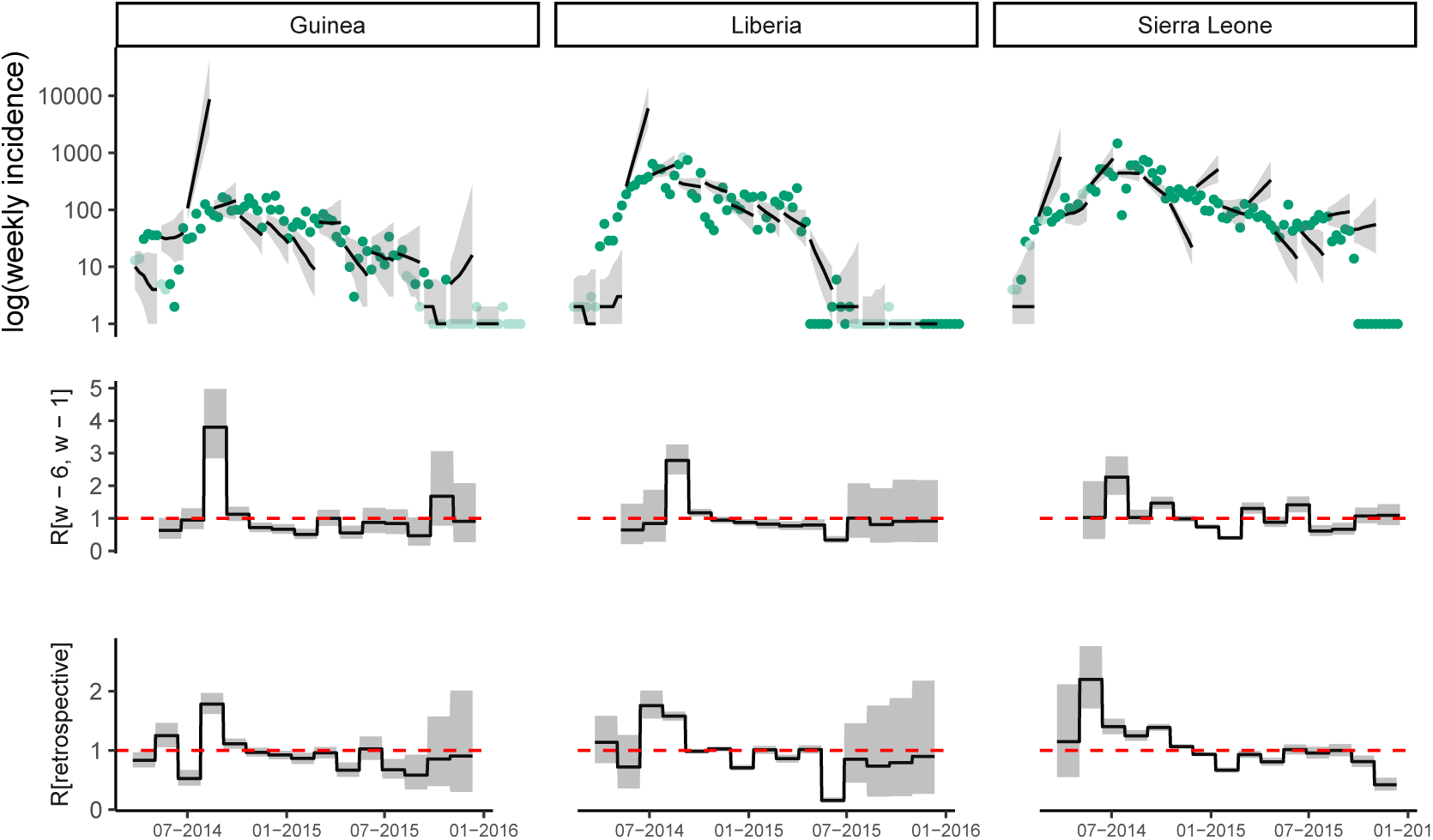
Observed and predicted incidence, and reproduction number estimates from HealthMap data. The top panel shows the weekly incidence derived from HealthMap data and the 6 weeks incidence forecast on log scale. The solid dots represent the observed weekly incidence where the light green dots show weeks for which all data points were obtained using interpolation. The projections are made over 6 week windows, based on the reproduction number estimated in the previous 6 weeks. The middle figure in each panel shows the reproduction number used to make forecasts over each 6 week forecast horizon. The bottom figure shows the effective reproduction number estimated retrospectively using the full dataset up to the length of one calibration window before the end. In each case, the solid black line is the median estimate and the shaded region represents the 95% Credible Interval. The red horizontal dashed line indicates the *R*_*t*_ = 1 threshold. Results are shown for the three mainly affected countries although the analysis was done jointly using data for all countries in Africa.

##### 3.3.3 Forecast horizon of 8 weeks

**Fig SI 22:**
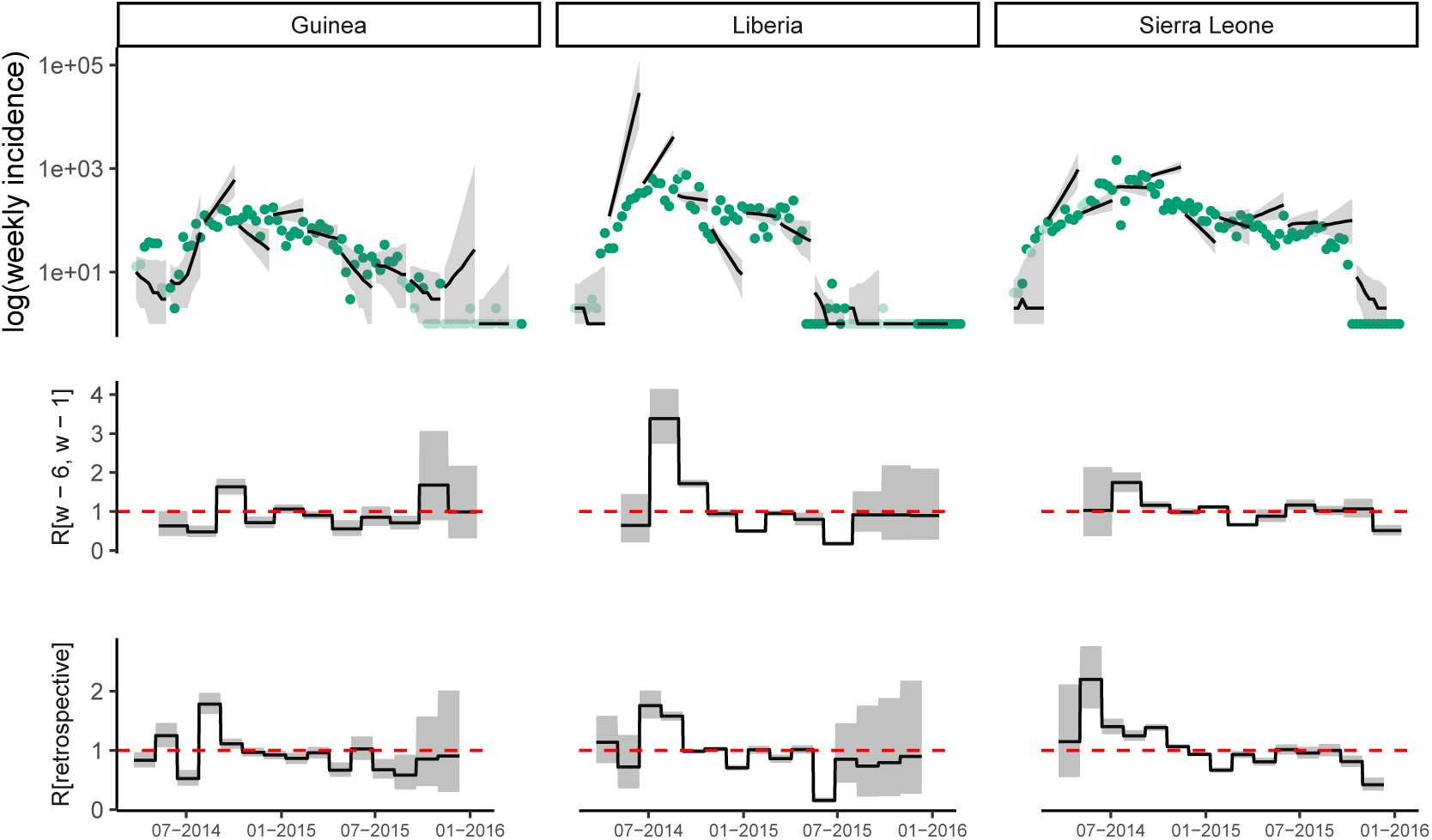
Observed and predicted incidence, and reproduction number estimates from HealthMap data. The top panel shows the weekly incidence derived from HealthMap data and the 8 weeks incidence forecast on log scale. The solid dots represent the observed weekly incidence where the light green dots show weeks for which all data points were obtained using interpolation. The projections are made over 8 week windows, based on the reproduction number estimated in the previous 6 weeks. The middle figure in each panel shows the reproduction number used to make forecasts over each 8 week forecast horizon. The bottom figure shows the effective reproduction number estimated retrospectively using the full dataset up to the length of one calibration window before the end. In each case, the solid black line is the median estimate and the shaded region represents the 95% Credible Interval. The red horizontal dashed line indicates the *R*_*t*_ = 1 threshold. Results are shown for the three mainly affected countries although the analysis was done jointly using data for all countries in Africa.

### 4 Forecasts using WHO data

This section presents the model forecasts over 4 6 and 6 weeks horizon produced using WHO data and calibration window of 2, 4 or 6 weeks.

#### 4.1 Calibration window of 2 weeks

##### 4.1.1 Forecast horizon of 4 weeks

**Fig SI 23:**
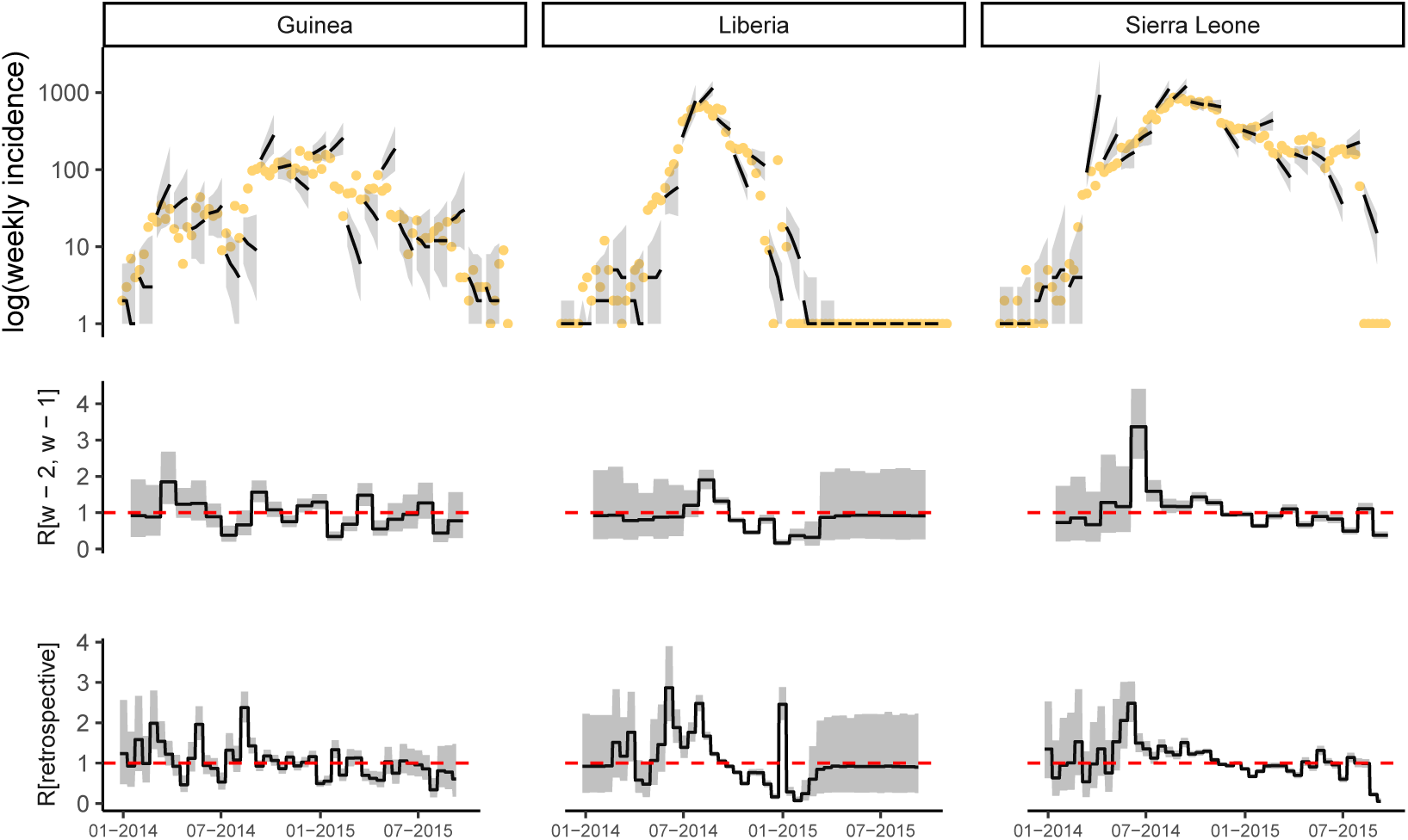
Observed and predicted incidence, and reproduction number estimates from WHO data. The top panel shows the weekly incidence derived from WHO data and the 4 weeks incidence forecast on log scale. The solid dots represent the observed weekly incidence. The projections are made over 4 week windows, based on the reproduction number estimated in the previous 2 weeks. The middle figure in each panel shows the reproduction number used to make forecasts over each 4 week forecast horizon. The bottom figure shows the effective reproduction number estimated retrospectively using the full dataset up to the length of one calibration window before the end. In each case, the solid black line is the median estimate and the shaded region represents the 95% Credible Interval. The red horizontal dashed line indicates the *R*_*t*_ = 1 threshold. Results are shown for the three mainly affected countries although the analysis was done jointly using data for all countries in Africa.

##### 4.1.2 Forecast horizon of 6 weeks

**Fig SI 24:**
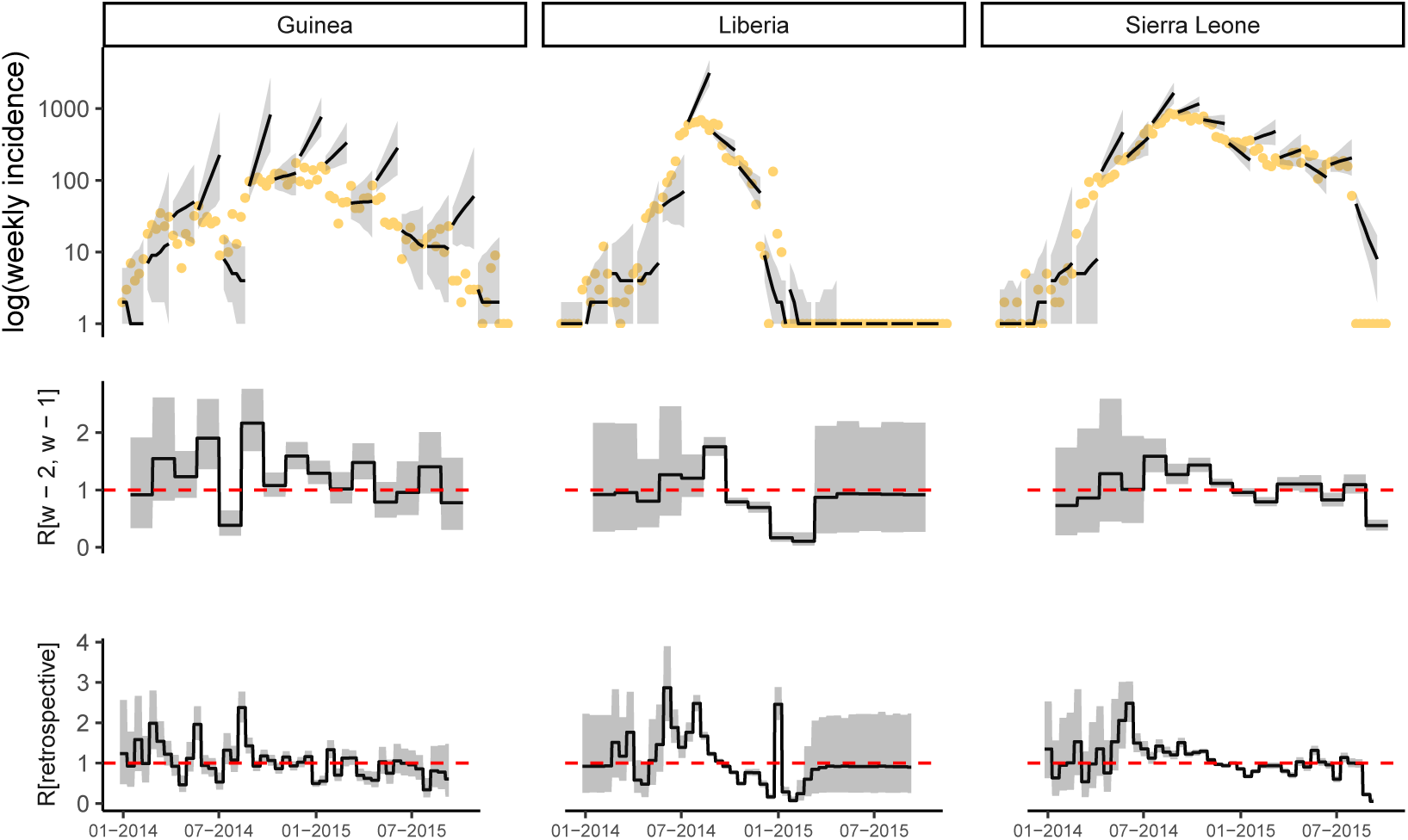
Observed and predicted incidence, and reproduction number estimates from WHO data. The top panel shows the weekly incidence derived from WHO data and the 6 weeks incidence forecast on log scale. The solid dots represent the observed weekly incidence. The projections are made over 6 week windows, based on the reproduction number estimated in the previous 2 weeks. The middle figure in each panel shows the reproduction number used to make forecasts over each 6 week forecast horizon. The bottom figure shows the effective reproduction number estimated retrospectively using the full dataset up to the length of one calibration window before the end. In each case, the solid black line is the median estimate and the shaded region represents the 95% Credible Interval. The red horizontal dashed line indicates the *R*_*t*_ = 1 threshold. Results are shown for the three mainly affected countries although the analysis was done jointly using data for all countries in Africa.

##### 4.1.3 Forecast horizon of 8 weeks

**Fig SI 25:**
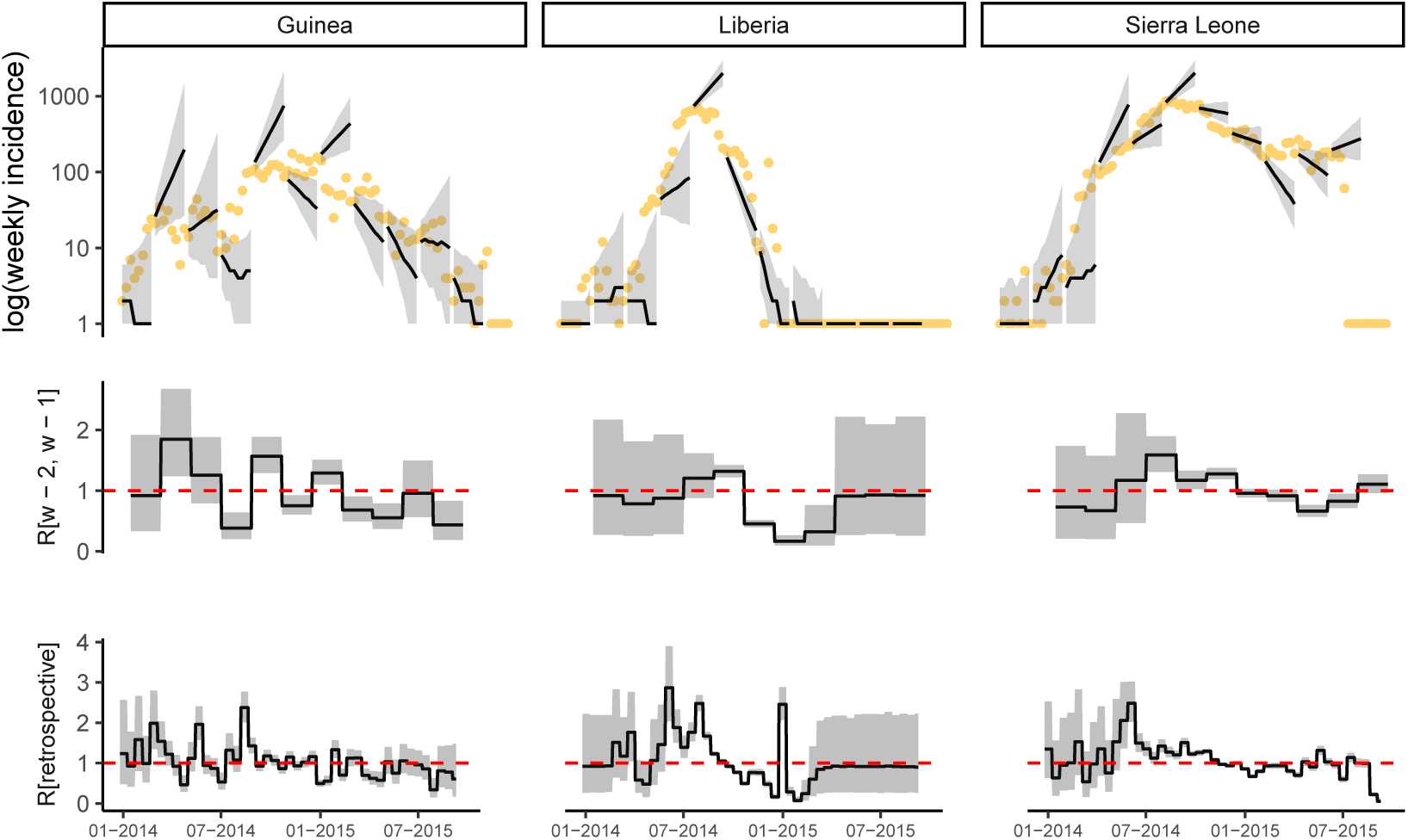
Observed and predicted incidence, and reproduction number estimates from WHO data. The top panel shows the weekly incidence derived from WHO data and the 8 weeks incidence forecast on log scale. The solid dots represent the observed weekly incidence. The projections are made over 8 week windows, based on the reproduction number estimated in the previous 2 weeks. The middle figure in each panel shows the reproduction number used to make forecasts over each 8 week forecast horizon. The bottom figure shows the effective reproduction number estimated retrospectively using the full dataset up to the length of one calibration window before the end. In each case, the solid black line is the median estimate and the shaded region represents the 95% Credible Interval. The red horizontal dashed line indicates the *R*_*t*_ = 1 threshold. Results are shown for the three mainly affected countries although the analysis was done jointly using data for all countries in Africa.

#### 4.2 Calibration window of 4 weeks

##### 4.2.1 Forecast horizon of 4 weeks

**Fig SI 26:**
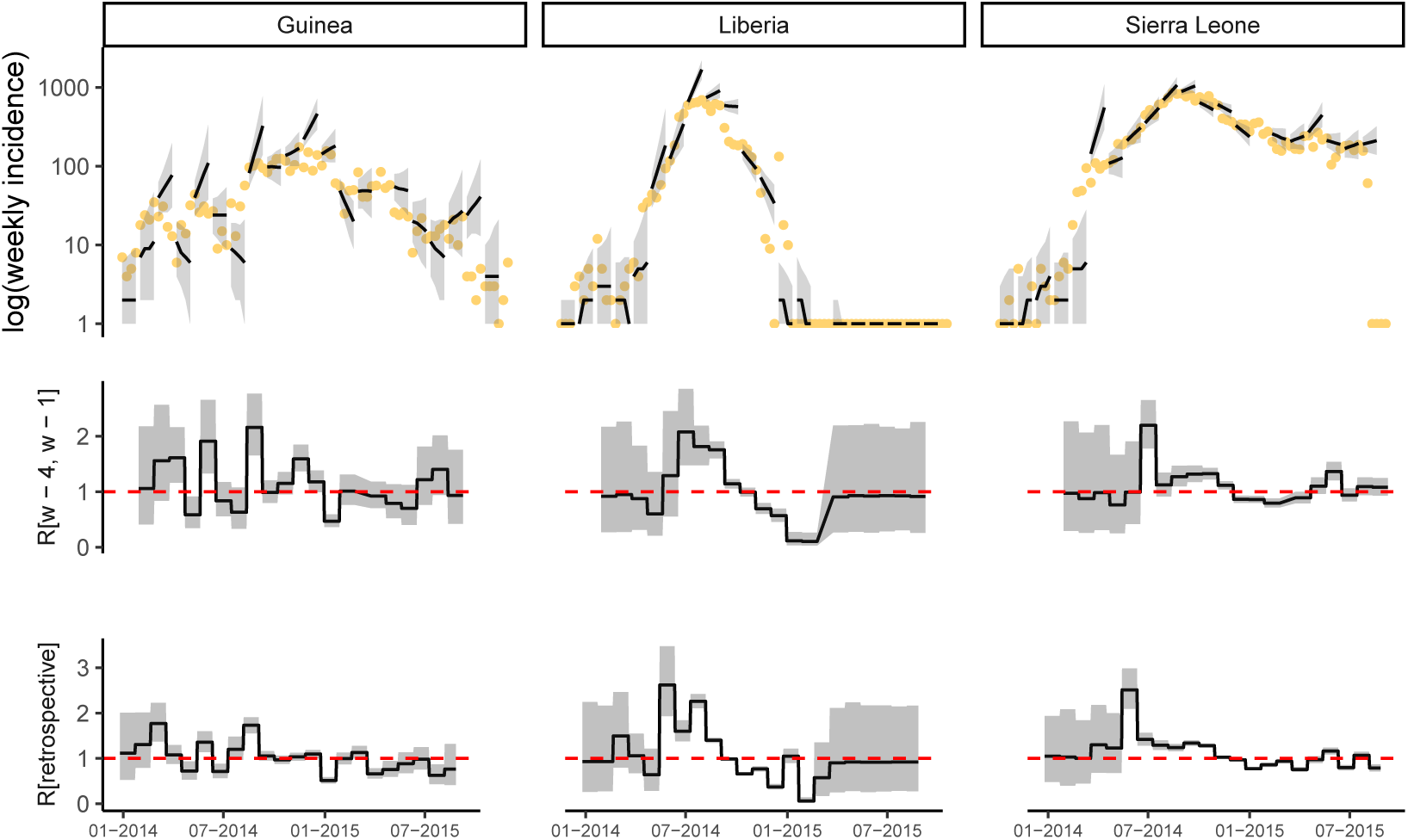
Observed and predicted incidence, and reproduction number estimates from WHO data. The top panel shows the weekly incidence derived from WHO data and the 4 weeks incidence forecast on log scale. The solid dots represent the observed weekly incidence. The projections are made over 4 week windows, based on the reproduction number estimated in the previous 4 weeks. The middle figure in each panel shows the reproduction number used to make forecasts over each 4 week forecast horizon. The bottom figure shows the effective reproduction number estimated retrospectively using the full dataset up to the length of one calibration window before the end. In each case, the solid black line is the median estimate and the shaded region represents the 95% Credible Interval. The red horizontal dashed line indicates the *R*_*t*_ = 1 threshold. Results are shown for the three mainly affected countries although the analysis was done jointly using data for all countries in Africa.

##### 4.2.2 Forecast horizon of 6 weeks

**Fig SI 27:**
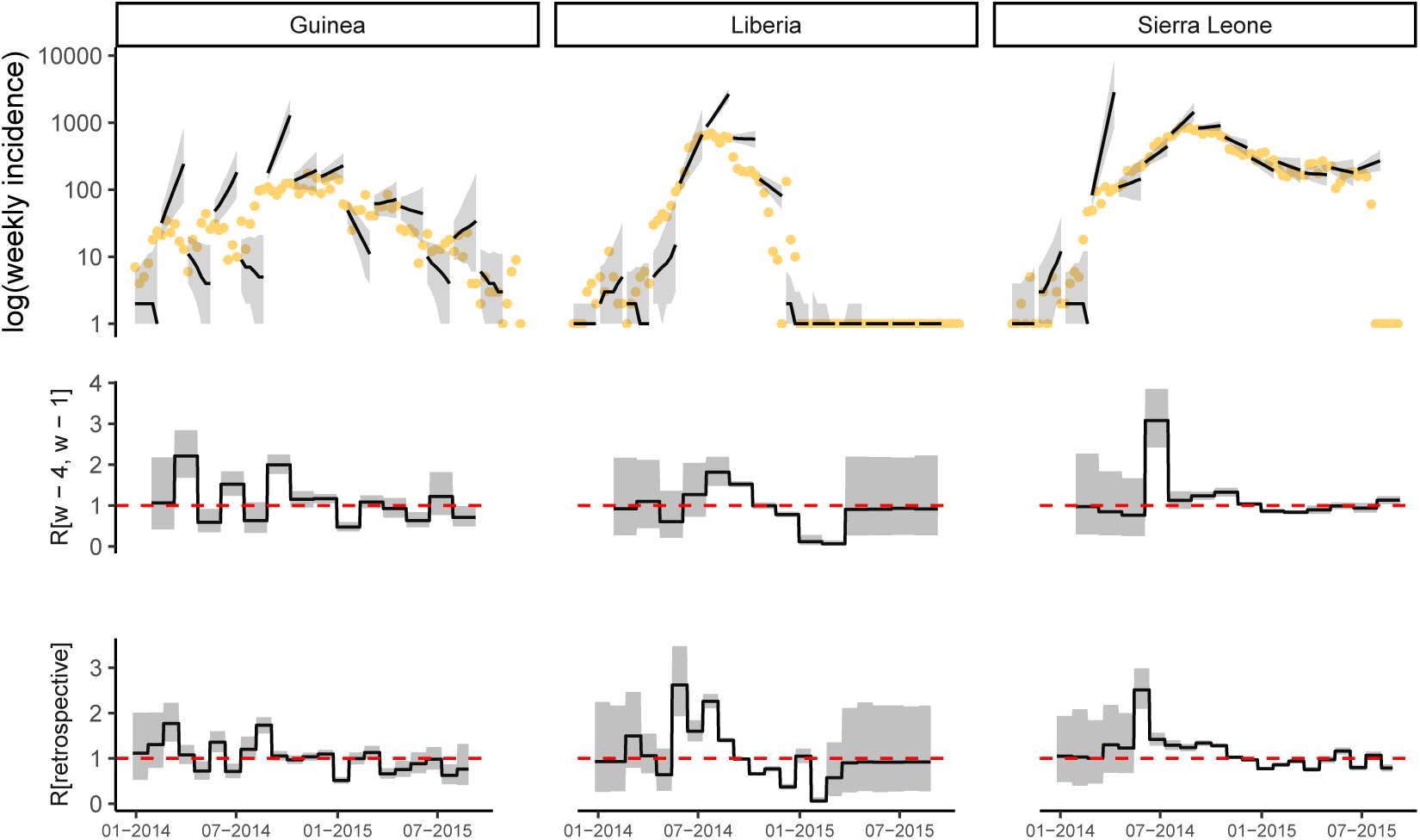
Observed and predicted incidence, and reproduction number estimates from WHO data. The top panel shows the weekly incidence derived from WHO data and the 6 weeks incidence forecast on log scale. The solid dots represent the observed weekly incidence. The projections are made over 6 week windows, based on the reproduction number estimated in the previous 4 weeks. The middle figure in each panel shows the reproduction number used to make forecasts over each 6 week forecast horizon. The bottom figure shows the effective reproduction number estimated retrospectively using the full dataset up to the length of one calibration window before the end. In each case, the solid black line is the median estimate and the shaded region represents the 95% Credible Interval. The red horizontal dashed line indicates the *R*_*t*_ = 1 threshold. Results are shown for the three mainly affected countries although the analysis was done jointly using data for all countries in Africa.

##### 4.2.3 Forecast horizon of 8 weeks

**Fig SI 28:**
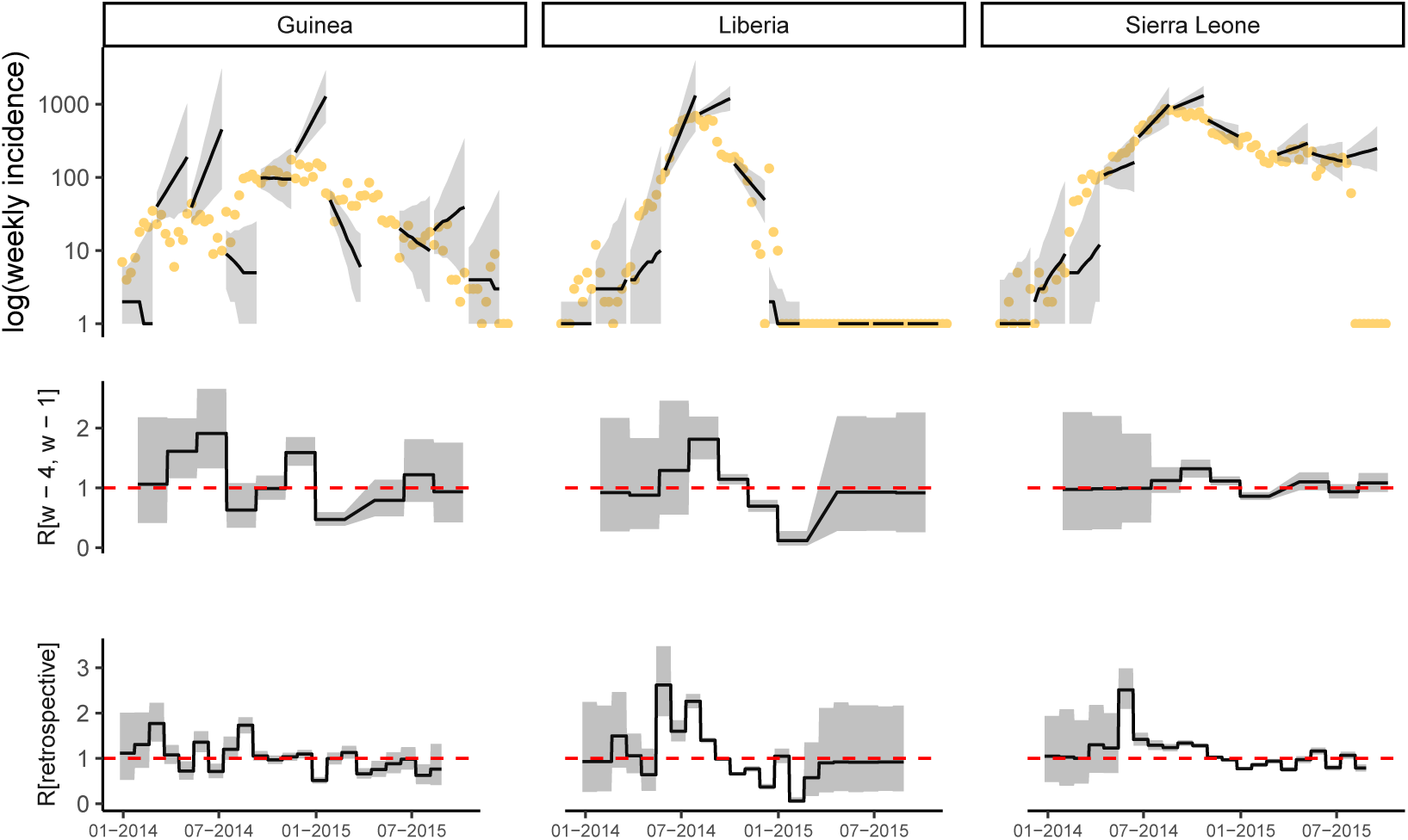
Observed and predicted incidence, and reproduction number estimates from WHO data. The top panel shows the weekly incidence derived from WHO data and the 8 weeks incidence forecast on log scale. The solid dots represent the observed weekly incidence. The projections are made over 8 week windows, based on the reproduction number estimated in the previous 4 weeks. The middle figure in each panel shows the reproduction number used to make forecasts over each 8 week forecast horizon. The bottom figure shows the effective reproduction number estimated retrospectively using the full dataset up to the length of one calibration window before the end. In each case, the solid black line is the median estimate and the shaded region represents the 95% Credible Interval. The red horizontal dashed line indicates the *R*_*t*_ = 1 threshold. Results are shown for the three mainly affected countries although the analysis was done jointly using data for all countries in Africa.

#### 4.3 Calibration window of 6 weeks

##### 4.3.1 Forecast horizon of 4 weeks

**Fig SI 29:**
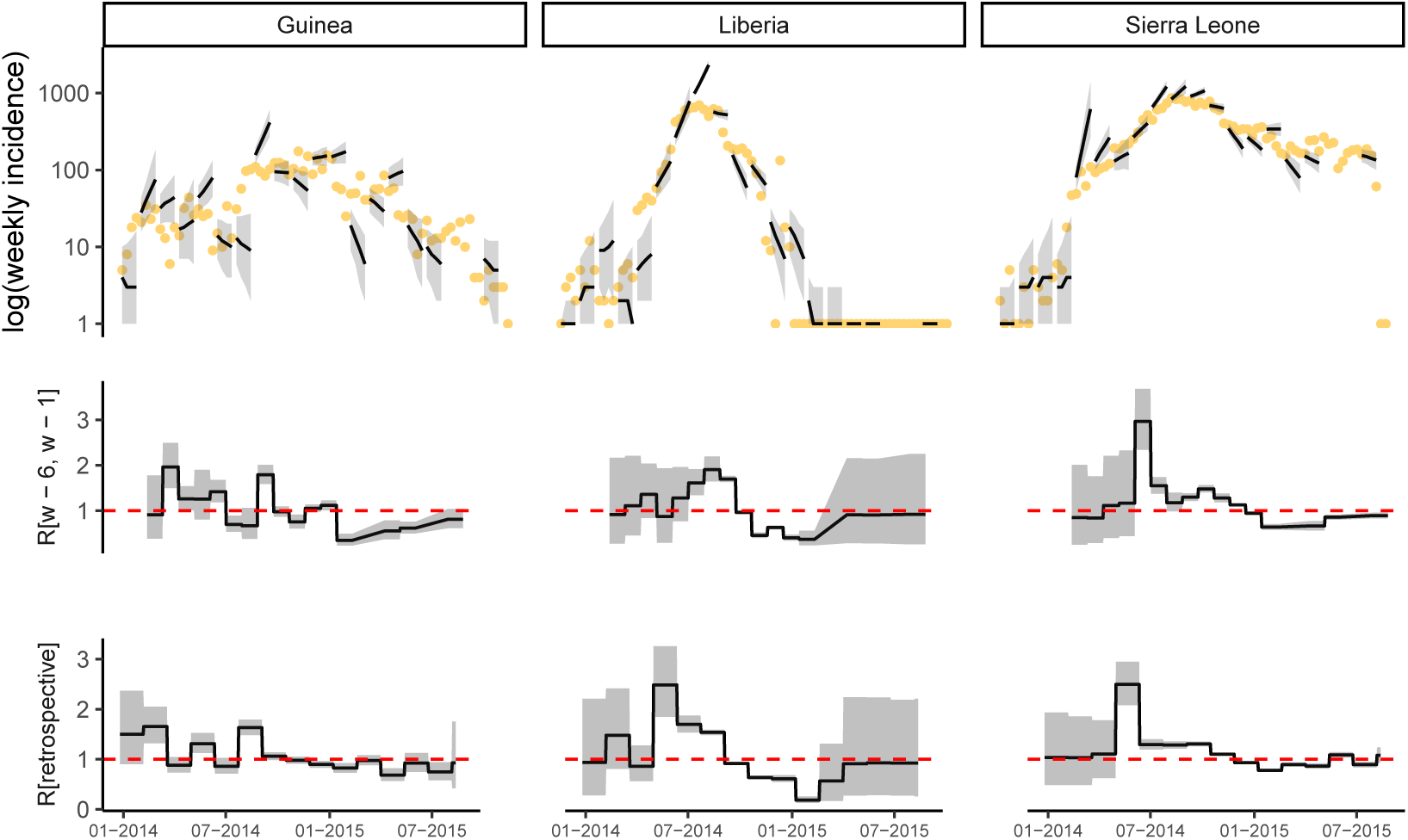
Observed and predicted incidence, and reproduction number estimates from WHO data. The top panel shows the weekly incidence derived from WHO data and the 4 weeks incidence forecast on log scale. The solid dots represent the observed weekly incidence. The projections are made over 4 week windows, based on the reproduction number estimated in the previous 6 weeks. The middle figure in each panel shows the reproduction number used to make forecasts over each 4 week forecast horizon. The bottom figure shows the effective reproduction number estimated retrospectively using the full dataset up to the length of one calibration window before the end. In each case, the solid black line is the median estimate and the shaded region represents the 95% Credible Interval. The red horizontal dashed line indicates the *R*_*t*_ = 1 threshold. Results are shown for the three mainly affected countries although the analysis was done jointly using data for all countries in Africa.

##### 4.3.2 Forecast horizon of 6 weeks

**Fig SI 30:**
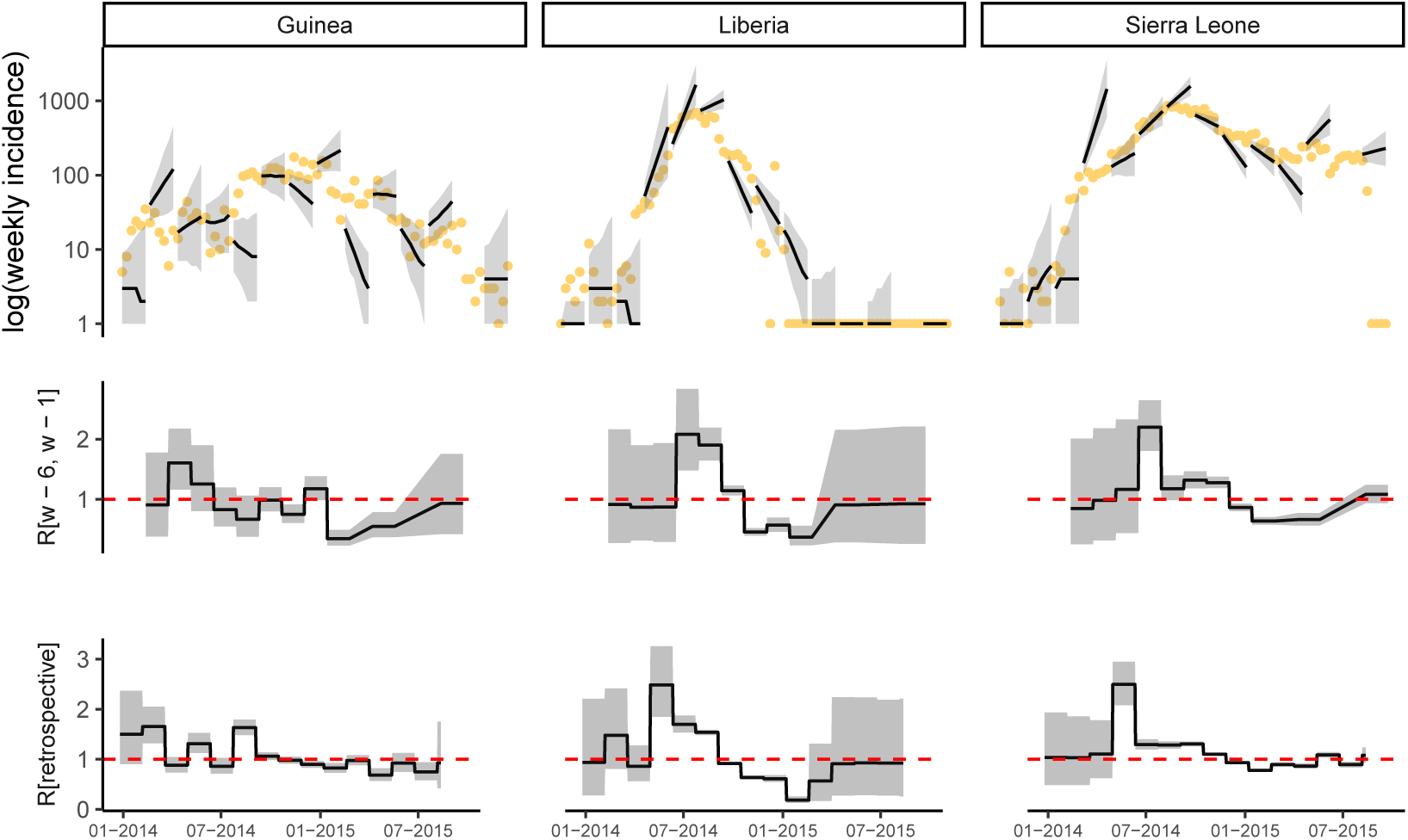
Observed and predicted incidence, and reproduction number estimates from WHO data. The top panel shows the weekly incidence derived from WHO data and the 6 weeks incidence forecast on log scale. The solid dots represent the observed weekly incidence. The projections are made over 6 week windows, based on the reproduction number estimated in the previous 6 weeks. The middle figure in each panel shows the reproduction number used to make forecasts over each 6 week forecast horizon. The bottom figure shows the effective reproduction number estimated retrospectively using the full dataset up to the length of one calibration window before the end. In each case, the solid black line is the median estimate and the shaded region represents the 95% Credible Interval. The red horizontal dashed line indicates the *R*_*t*_ = 1 threshold. Results are shown for the three mainly affected countries although the analysis was done jointly using data for all countries in Africa.

##### 4.3.3 Forecast horizon of 8 weeks

**Fig SI 31:**
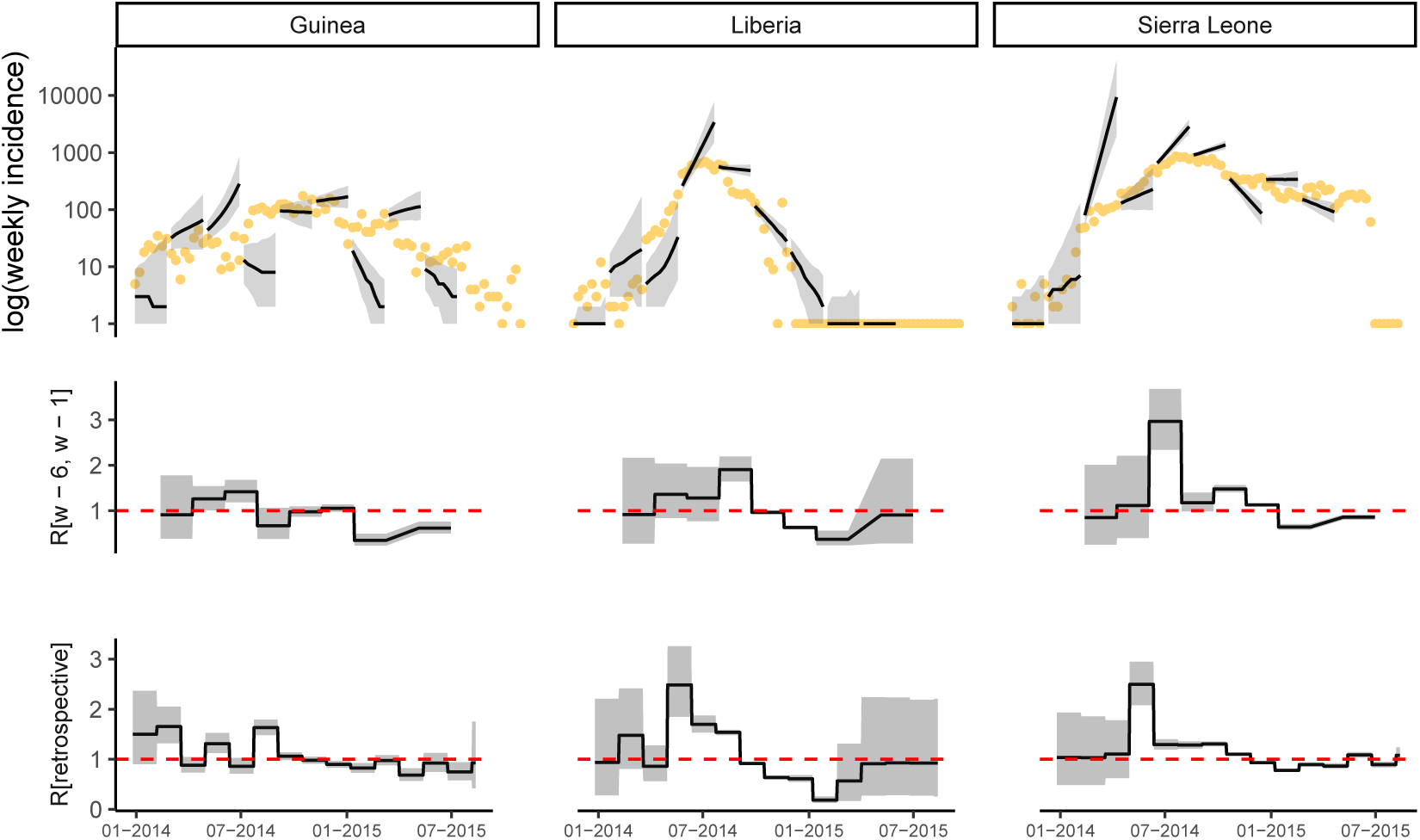
Observed and predicted incidence, and reproduction number estimates from WHO data. The top panel shows the weekly incidence derived from WHO data and the 8 weeks incidence forecast on log scale. The solid dots represent the observed weekly incidence. The projections are made over 8 week windows, based on the reproduction number estimated in the previous 6 weeks. The middle figure in each panel shows the reproduction number used to make forecasts over each 8 week forecast horizon. The bottom figure shows the effective reproduction number estimated retrospectively using the full dataset up to the length of one calibration window before the end. In each case, the solid black line is the median estimate and the shaded region represents the 95% Credible Interval. The red horizontal dashed line indicates the *R*_*t*_ = 1 threshold. Results are shown for the three mainly affected countries although the analysis was done jointly using data for all countries in Africa.

### 5 Impact of calibration window on model performance

Since changing the length of the calibration window modified the model complexity with shorter windows introducing more parameters in the model, we assessed the impact of this length on the performance of the model (Fig 32).

**Fig SI 32:**
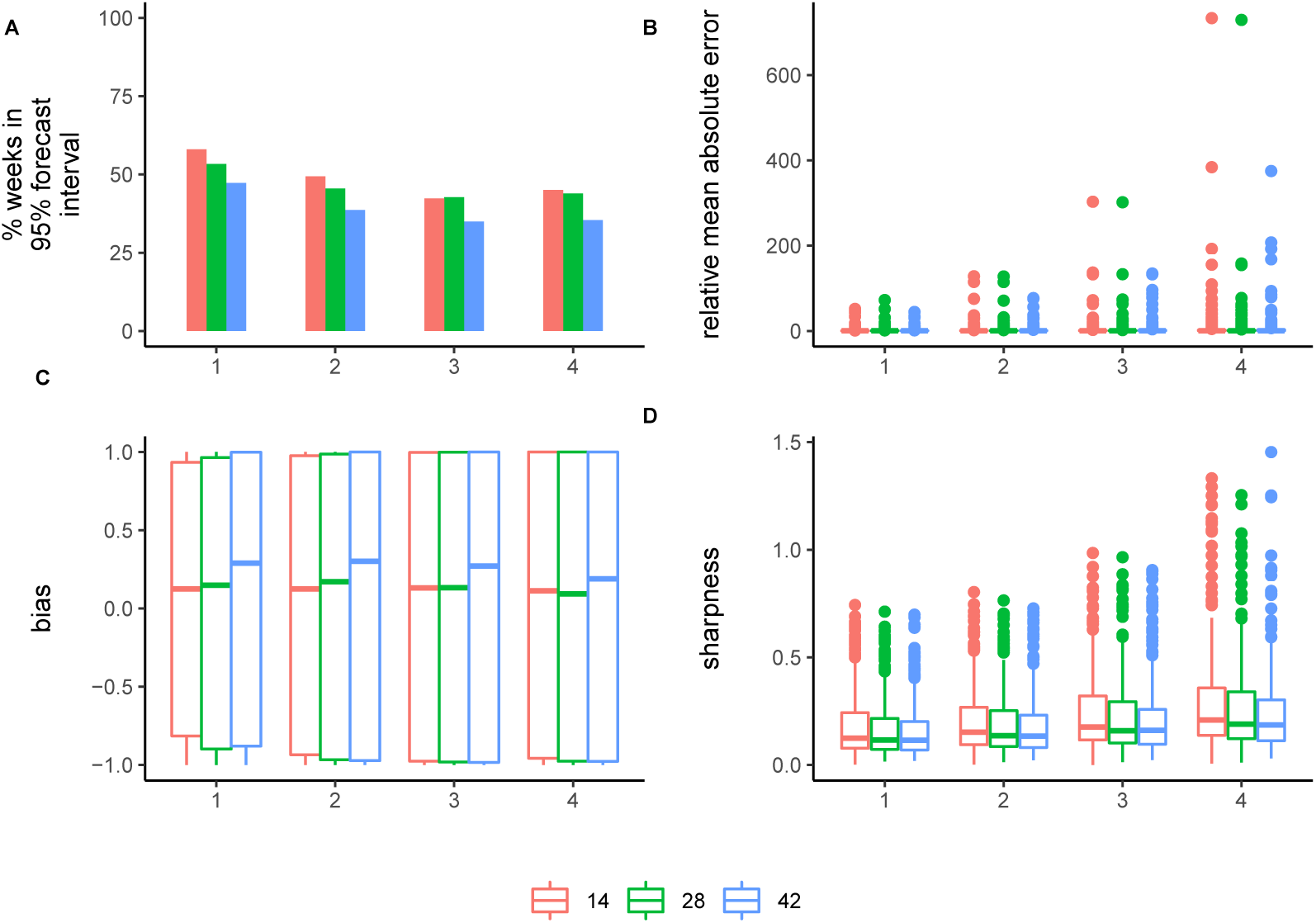
Model performance metrics stratified by the time window used for model calibration and week of projection. The performance metrics are (A) the percentage of weeks for which the 95% forecast interval contained the observed incidence, (B) relative mean absolute error, (C) bias, and (D) sharpness. See Methods for details.

### 6 Impact of datasource on model performance

**Fig SI 33:**
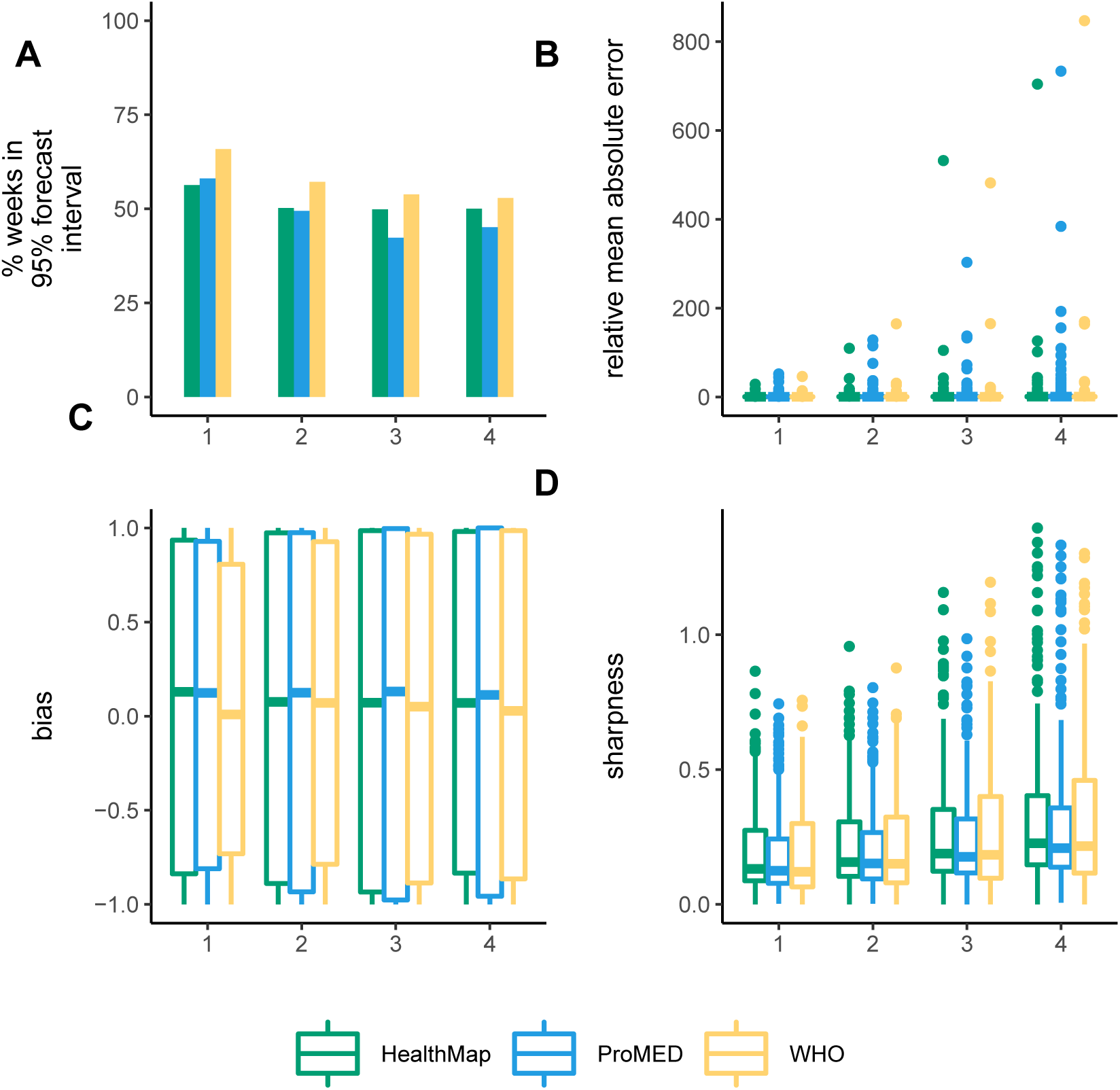
Model performance metrics stratified by datasource ProMED (blue), HealthMap (green), and WHO (yellow) and week of projection. The performance metrics are (A) the percentage of weeks for which the 95% forecast interval contained the observed incidence, (B) relative mean absolute error, (C) bias, and (D) sharpness. See Methods for details.

### 7 Mobility Model Parameters

We estimated the parameters of gravity model - *p*_*stay*_ which is the probability of an infectious case staying within a country, and *γ* which measures the extent to which distance between two locations influences the flow of people between them (Fig 34).

**Fig SI 34:**
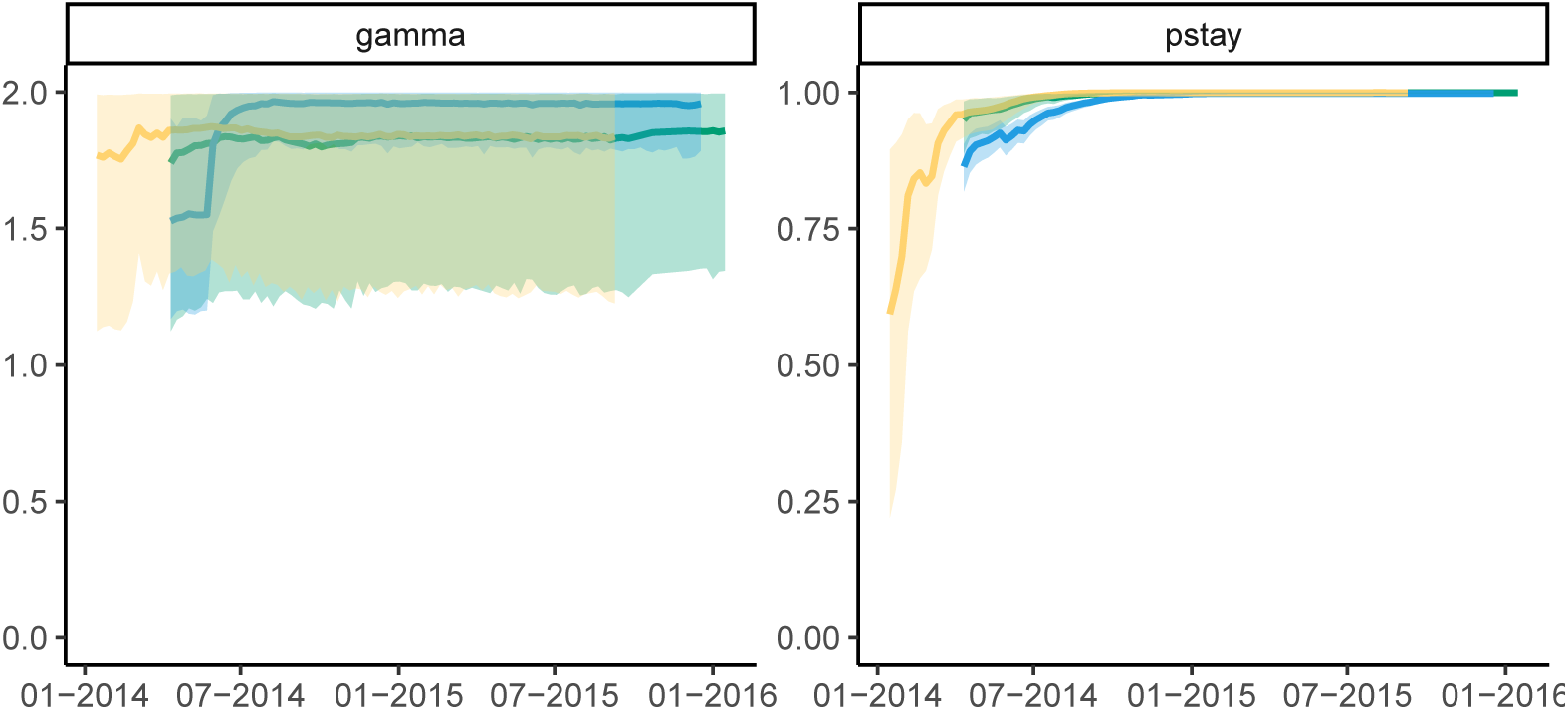
Estimates of mobility model parameters during the epidemic. Population movement was modelled using a gravity model where the flow between locations *i* and *j* is proportional to the product of their populations and inversely population to the distance between them raised to an exponent *γ*. The parameter *γ* thus modulates the influence of distance on the population flow. *p*_*stay*_ represents the probability of an individual to stay in a given location during their infectious period. The solid lines represents the median estimates obtained using WHO (yellow), ProMED (blue) and HealthMap (green) data. The shaded regions represent the 95% CrI.

### 8 Sensitivity Analysis

For the results presented in the main text, we choose an uninformative uniform prior for the parameter *γ* with an upper bound 2. We also fitted the model with a uniform prior for *γ* allowing it to vary from 1 to 10. In this section we present the results from the sensitivity analyses. Fig 34 presents the estimates of the parameters over the course of the epidemic using the alternative priors. As the epidemic progressed, the parameter *p*_*stay*_ assumed larger values suggesting a decreased probability of travel over time (Figures 34, 35). As *p*_*stay*_ assumes large values, the estimated flow is more strongly influenced by *p*_*stay*_ than by *γ*. Furthermore, *p*_*stay*_ is likely to depend on the spatial scale of the model. Our analyses were carried out at the national scale; we expect that *γ* will be more sensitive to *p*_*stay*_ at a finer spatial resolution. Overall, the flow between locations using the parameters estimated using the two alternative priors did not vary much (Fig 36). Figures 37 to 45 present the model forecasts using the alternative priors for *γ* and Figure 46 presents a comparison of model performance metrics using the two priors. Although the analysis was carried out for the three data sources (ProMED, HealthMap and WHO), for brevity we present results using ProMED data only.

**Fig SI 35:**
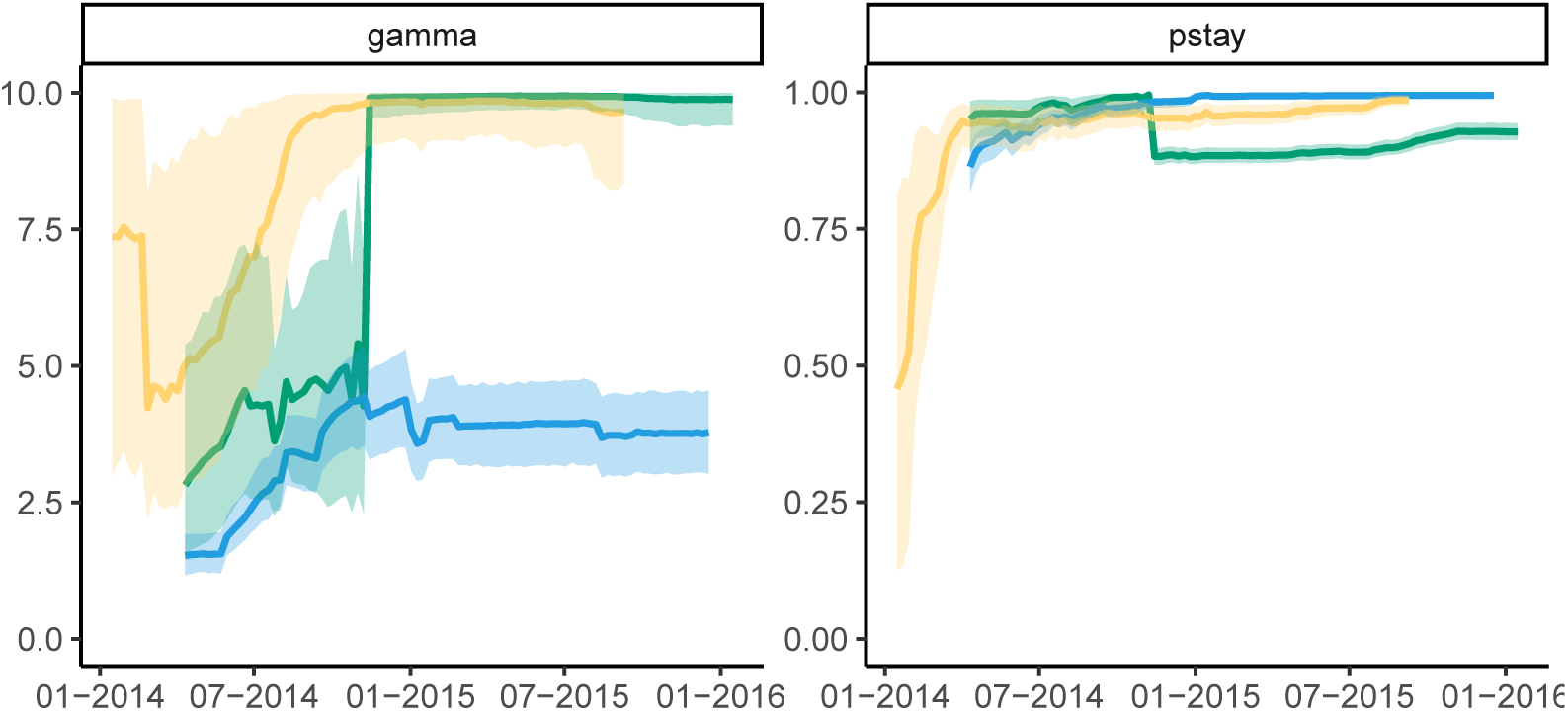
Estimates of mobility model parameters during the epidemic. Population movement was modelled using a gravity model where the flow between locations *i* and *j* is proportional to the product of their populations and inversely population to the distance between them raised to an exponent *γ*. The parameter *γ* thus modulates the influence of distance on the population flow. Here *γ* is allowed to vary between 1 and 10. *p*_*stay*_ represents the probability of an individual to stay in a given location during their infectious period. The solid lines represents the median estimates obtained using ProMED (blue), HealthMap (green) and WHO (yellow) data. The shaded regions represent the 95% CrI.

**Fig SI 36:**
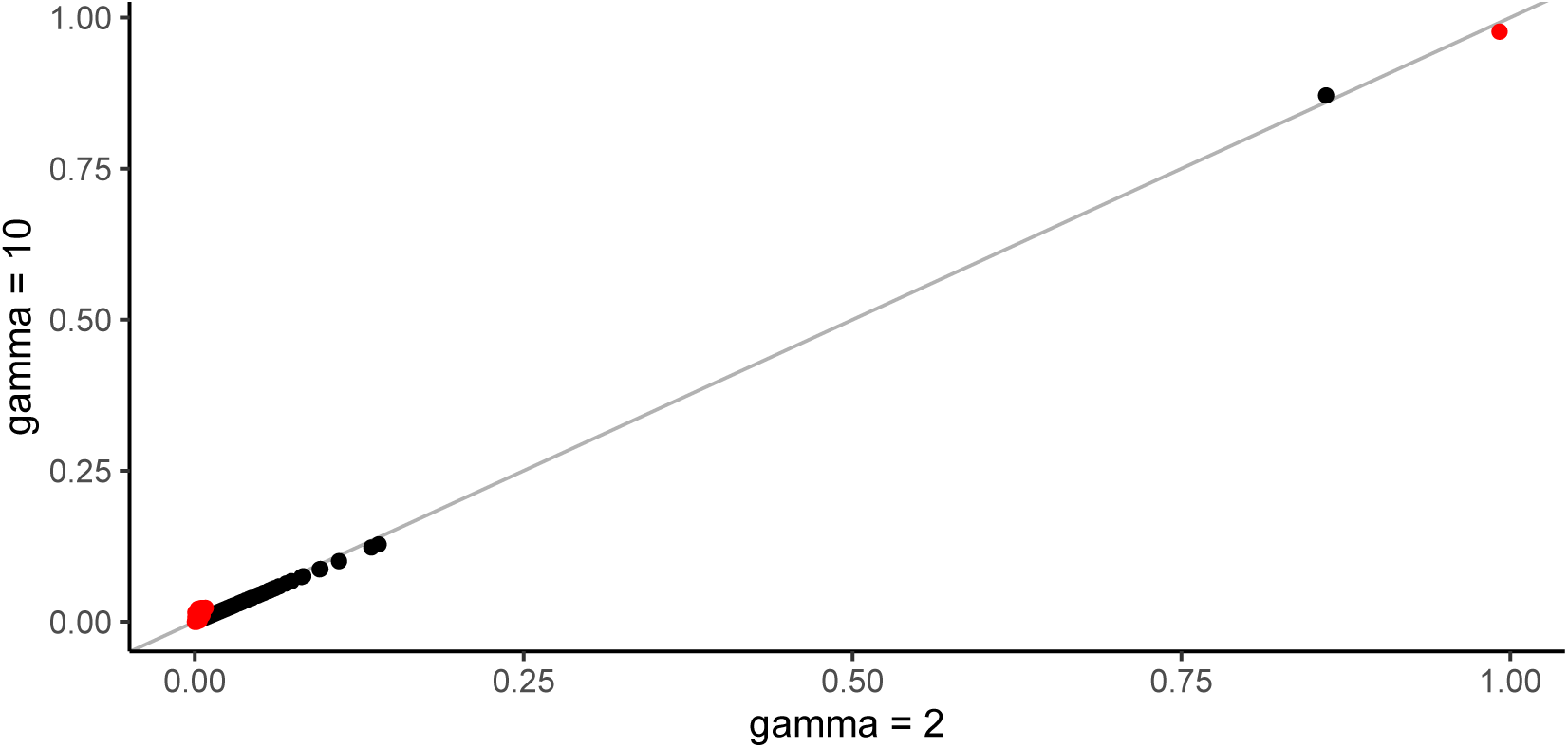
Estimated flows using gravity model. The x-axis shows the flows using a uniform prior for *γ* with upper limit 2 and the y-axis shows the flows using a uniform prior varying from 1 to 10. The black dots show the flows estimated using the first 21 days of incidence data from ProMED. Flows estimated using parameters fitted to the first 210 days of incidence data are shown in red. Results are shown for the model with calibration window set to 14 days.

#### 8.1 Forecasts using ProMED data

##### 8.1.1 Calibration window of 2 weeks

##### 8.1.2 Forecast horizon 4 weeks

**Fig SI 37:**
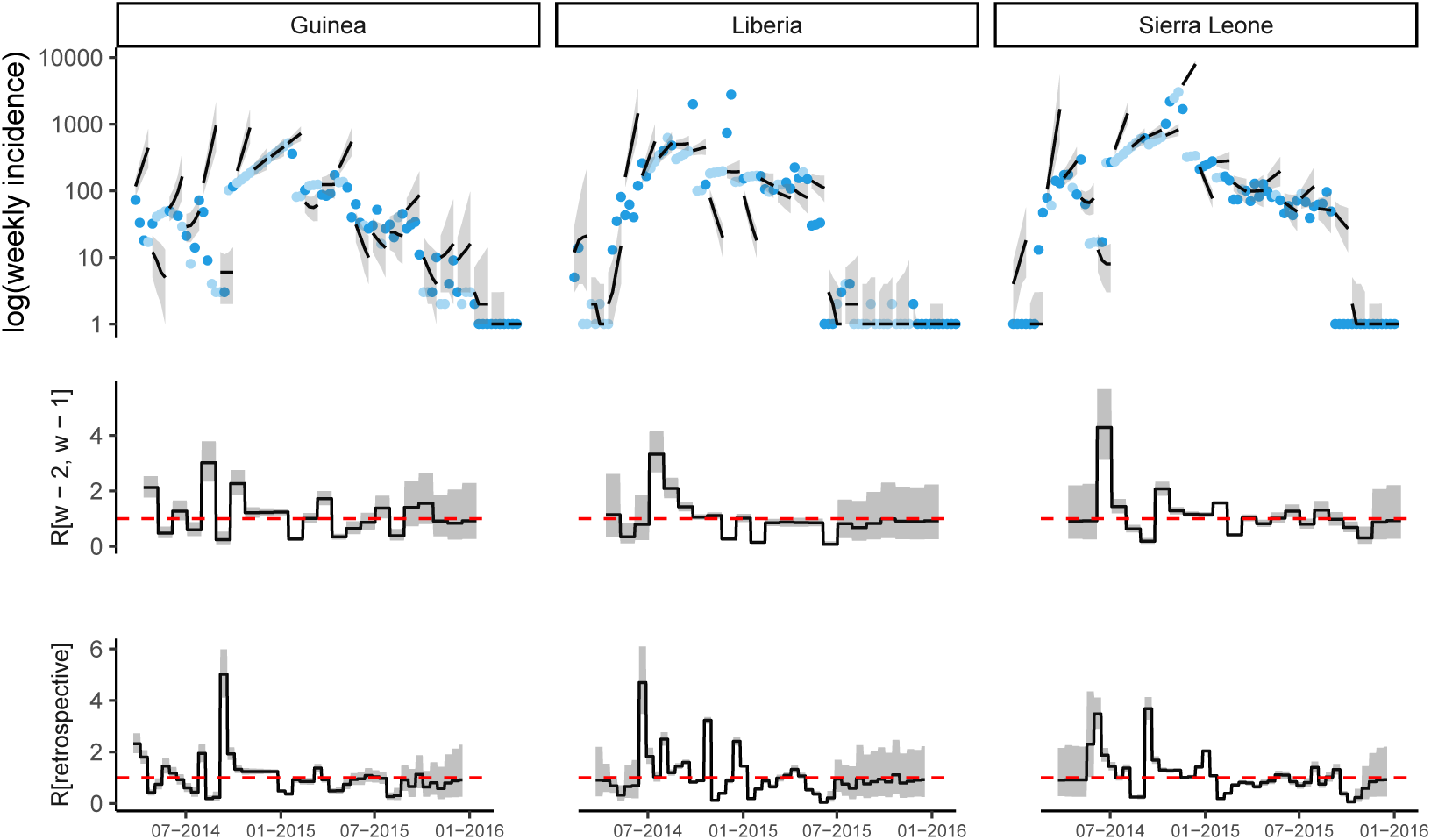
Observed and predicted incidence, and reproduction number estimates from ProMED data. The calibration window is 2 weeks and the forecast horizon is 4 weeks.

##### 8.1.3 Forecast horizon 6 weeks

**Fig SI 38:**
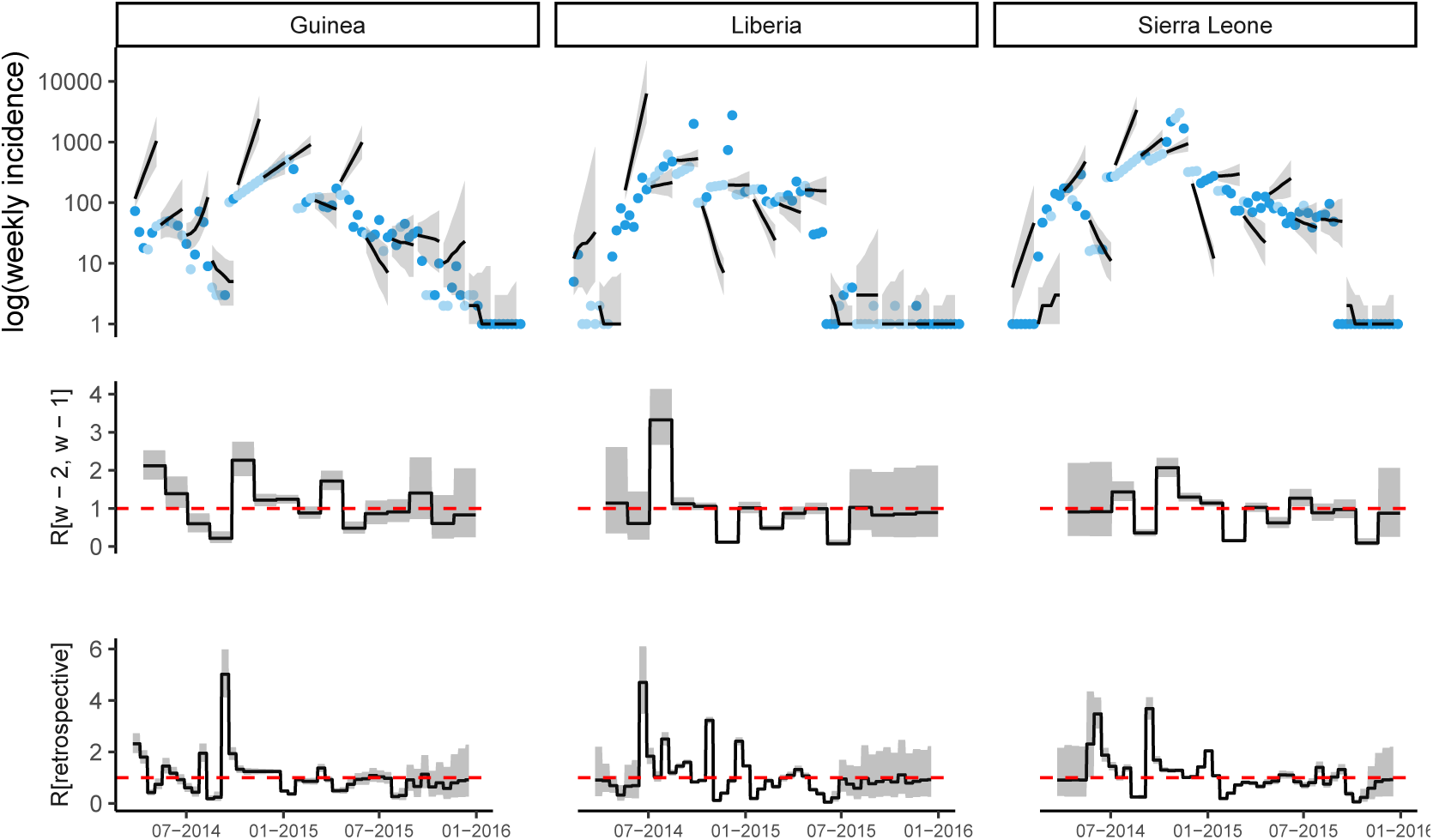
Observed and predicted incidence, and reproduction number estimates from ProMED data.The calibration window is 2 weeks and the forecast horizon is 6 weeks.

##### 8.1.4 Forecast horizon 8 weeks

**Fig SI 39:**
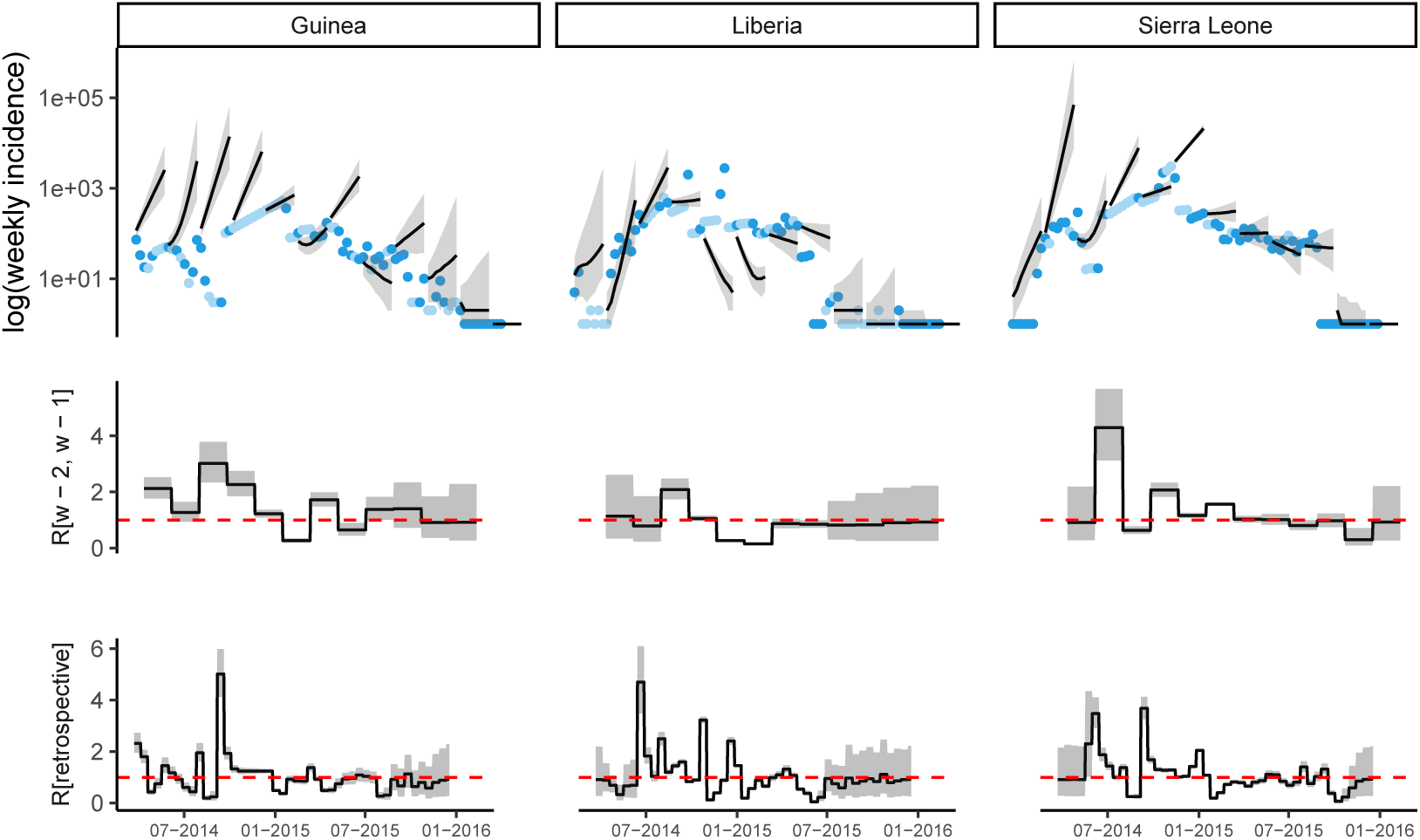
Observed and predicted incidence, and reproduction number estimates from ProMED data.The calibration window is 2 weeks and the forecast horizon is 8 weeks.

##### 8.1.5 Calibration window of 4 weeks

##### 8.1.6 Forecast horizon 4 weeks

**Fig SI 40:**
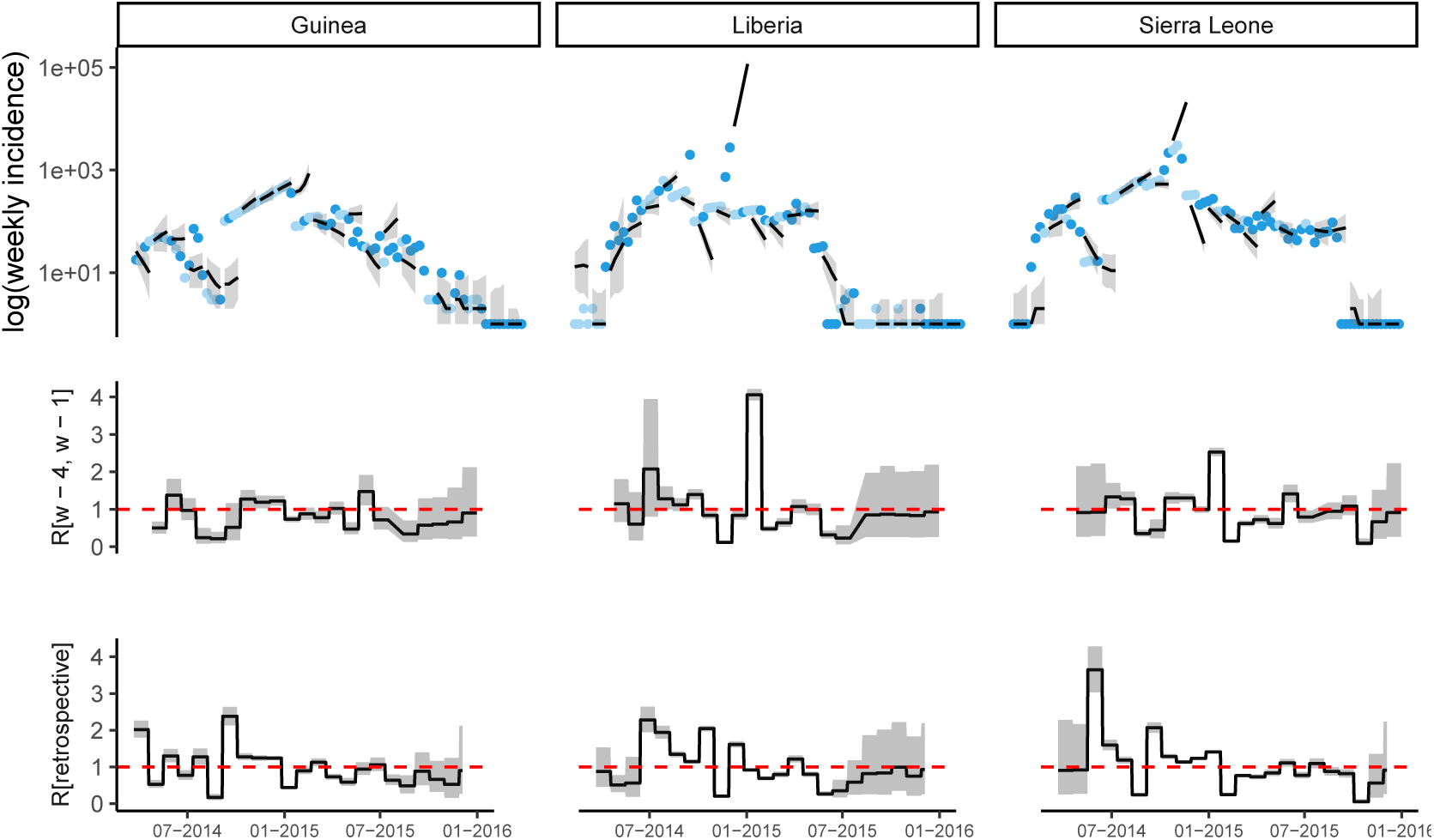
Observed and predicted incidence, and reproduction number estimates from ProMED data.The calibration window is 4 weeks and the forecast horizon is 4 weeks.

##### 8.1.7 Forecast horizon 6 weeks

**Fig SI 41:**
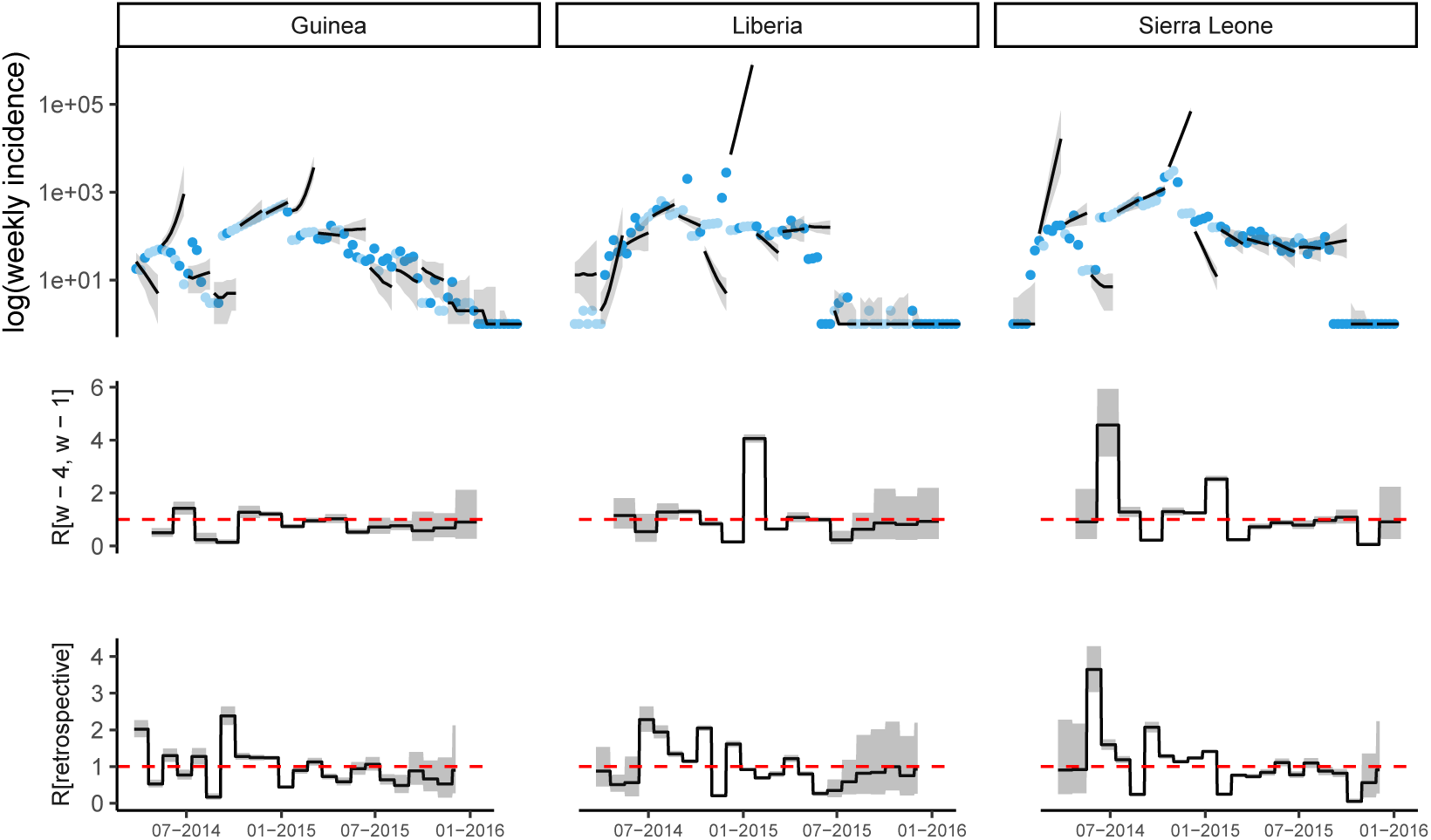
Observed and predicted incidence, and reproduction number estimates from ProMED data.The calibration window is 4 weeks and the forecast horizon is 6 weeks.

##### 8.1.8 Forecast horizon 8 weeks

**Fig SI 42:**
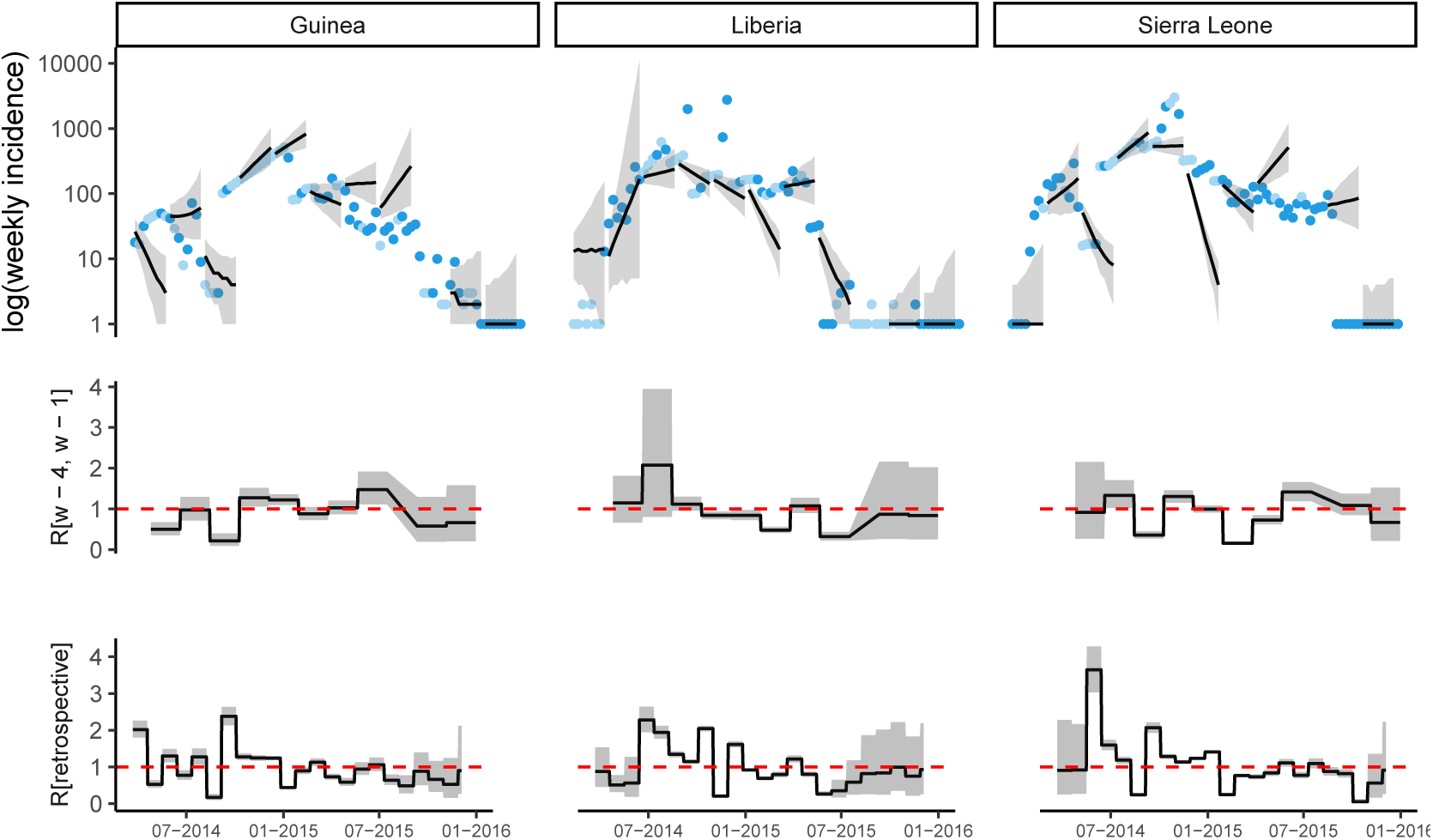
Observed and predicted incidence, and reproduction number estimates from ProMED data.The calibration window is 4 weeks and the forecast horizon is 8 weeks.

##### 8.1.9 Calibration window of 6 weeks

##### 8.1.10 Forecast horizon 4 weeks

**Fig SI 43:**
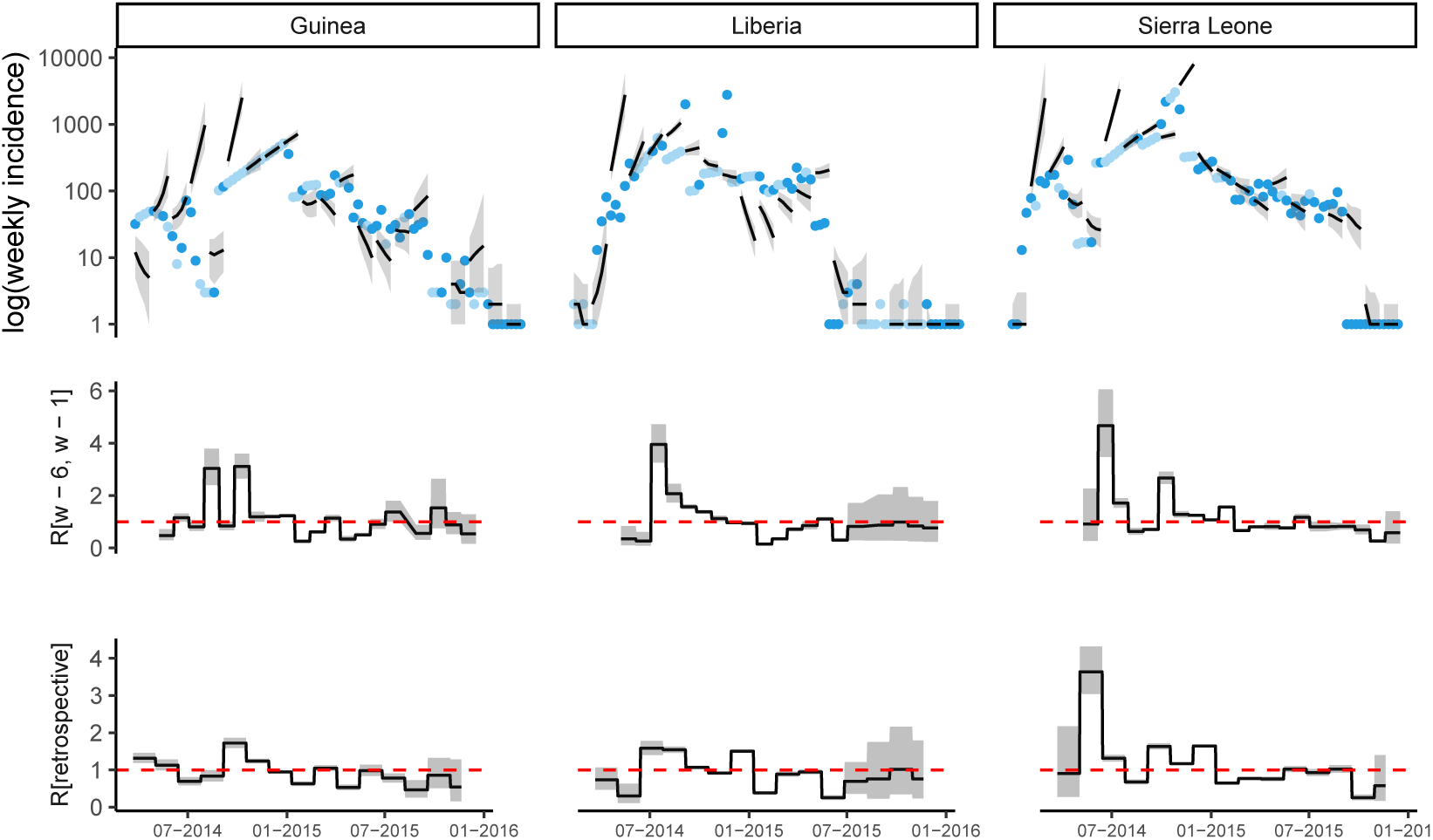
Observed and predicted incidence, and reproduction number estimates from ProMED data.The calibration window is 6 weeks and the forecast horizon is 4 weeks.

##### 8.1.11 Forecast horizon 6 weeks

**Fig SI 44:**
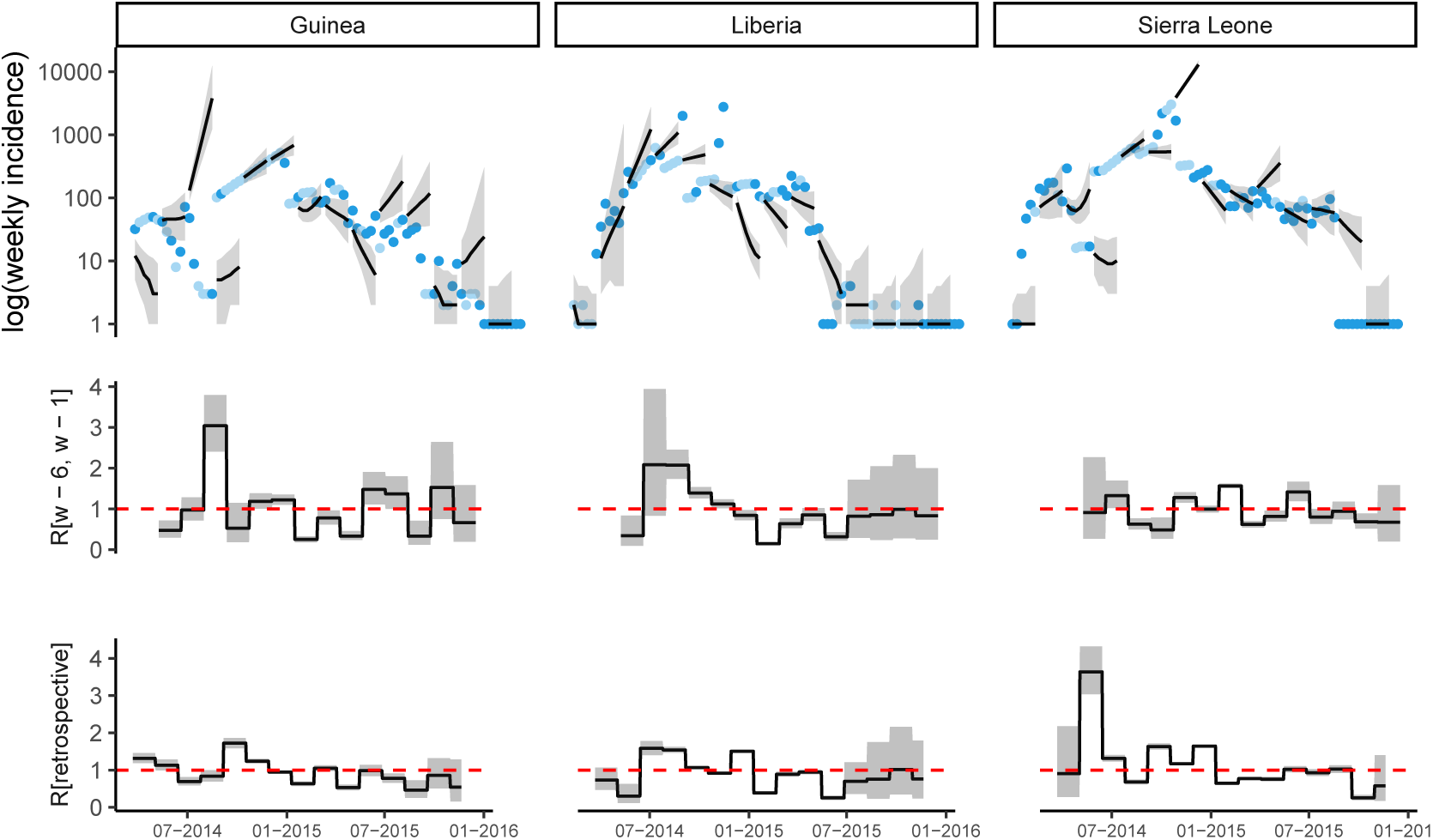
Observed and predicted incidence, and reproduction number estimates from ProMED data.The calibration window is 6 weeks and the forecast horizon is 6 weeks.

##### 8.1.12 Forecast horizon 8 weeks

**Fig SI 45:**
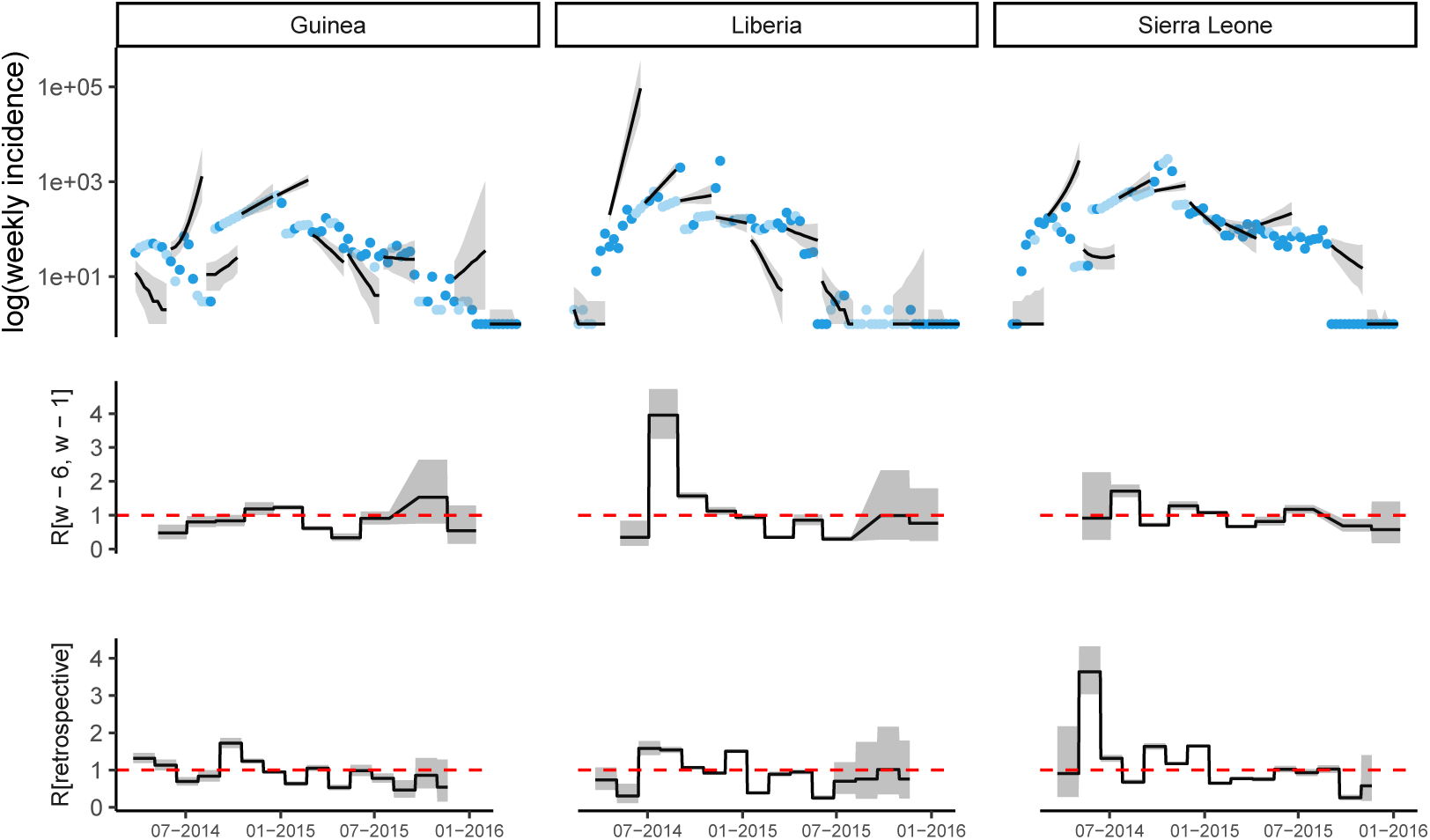
Observed and predicted incidence, and reproduction number estimates from ProMED data.The calibration window is 6 weeks and the forecast horizon is 8 weeks.

#### 8.2 Model performance with alternate priors for mobility model parameter

**Fig SI 46:**
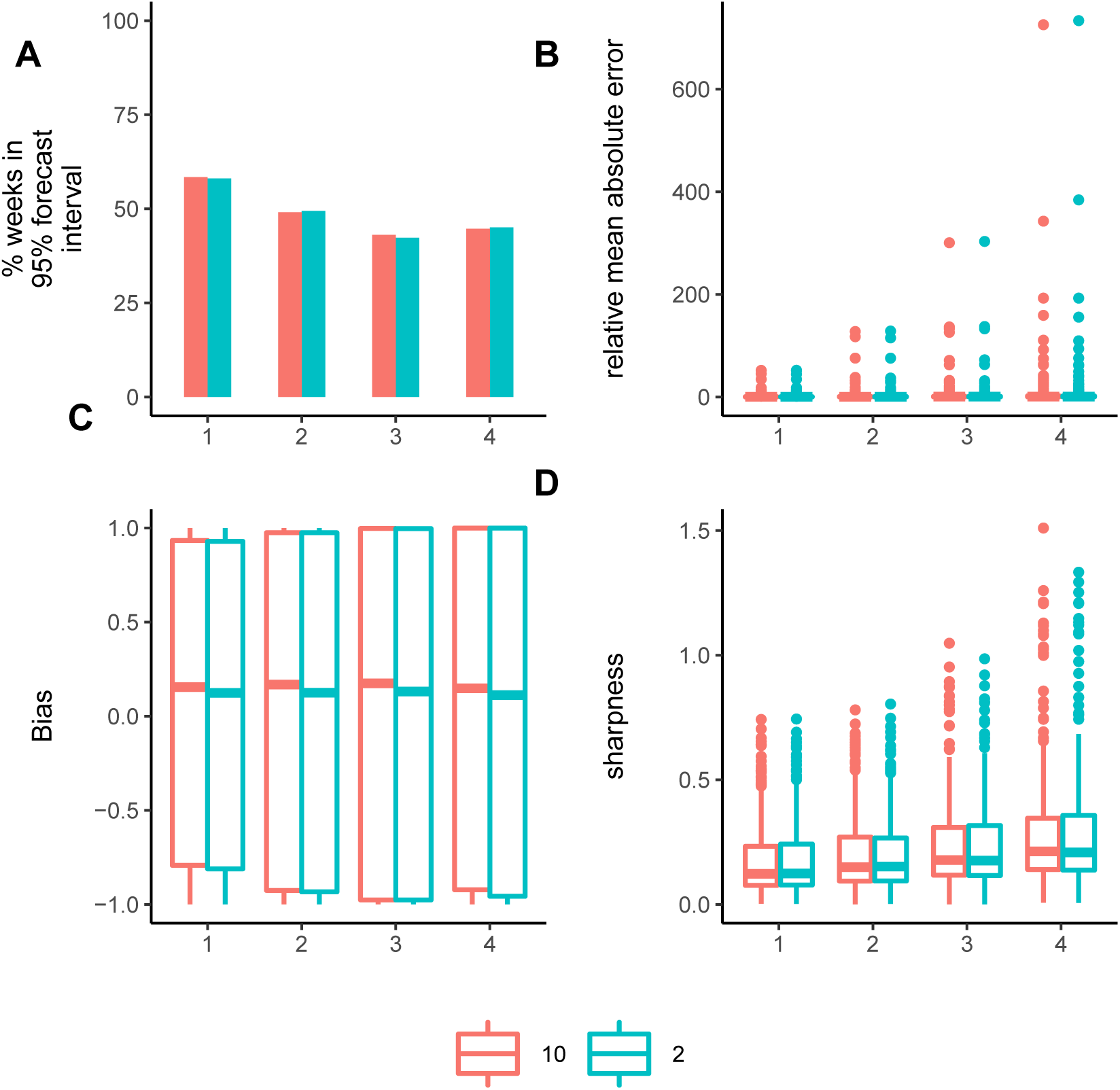
Model performance metrics allowing *γ* to vary up to 2 or 10. The performance metrics (A) the percentage of weeks for which the 95% forecast interval contained the observed incidence, (B) relative mean absolute error, (C) bias and (D) sharpness.

### 9 Risk of spatial spread

In this section, we present additional analyses carried out on predicting the spatial spread of the epidemic. Classification of alerts raised 1, 2, 3, and 4 weeks ahead are shown in Fig 47. We also assessed the sensitivity of the model when the analysis was restricted to countries other than Guinea, Liberia and Sierra Leone (Fig 48). At 93% threshold, the model exhibited high specificity (83.3%) and sensitivity (82.0%) in predicting presence of cases in weeks following a week with no observed cases in each country (Fig 49). In predicting presence of cases in countries with no or low incidence, or in a week following a week in which no cases were observed, the sensitivity improved at higher thresholds with a reasonably low false alert rate (Fig 50). Finally, we find that the model is able to attain a high sensitivity and specificity relatively early in the epidemic using all three data sources (Fig 51). Since WHO data used here were only available for Guinea, Liberia and Sierra Leone, we affixed data from ProMED for all countries other than these three to WHO data for the purpose of classifying alerts.

For a given threshold (*x*^*th*^ percentile of the forecast interval), we defined a True alert for a week where the *x*^*th*^ percentile of the forecast interval and the observed incidence for a country were both greater than 0; false alert for a week where the threshold for a country was greater than 0 but the observed incidence for that country was 0; and missed alert for a week where the threshold for a country was 0 but the observed incidence for that country was greater than 0. True alert rate is the ratio of correctly classified true alerts to the total number of true and missed alerts (i.e., (true alerts)/(true alerts + missed alerts)). False alert rate is similarly the ratio of false alerts to the total number of false alerts and weeks of no alert (where the observed and the threshold incidence are both 0).

**Fig SI 47:**
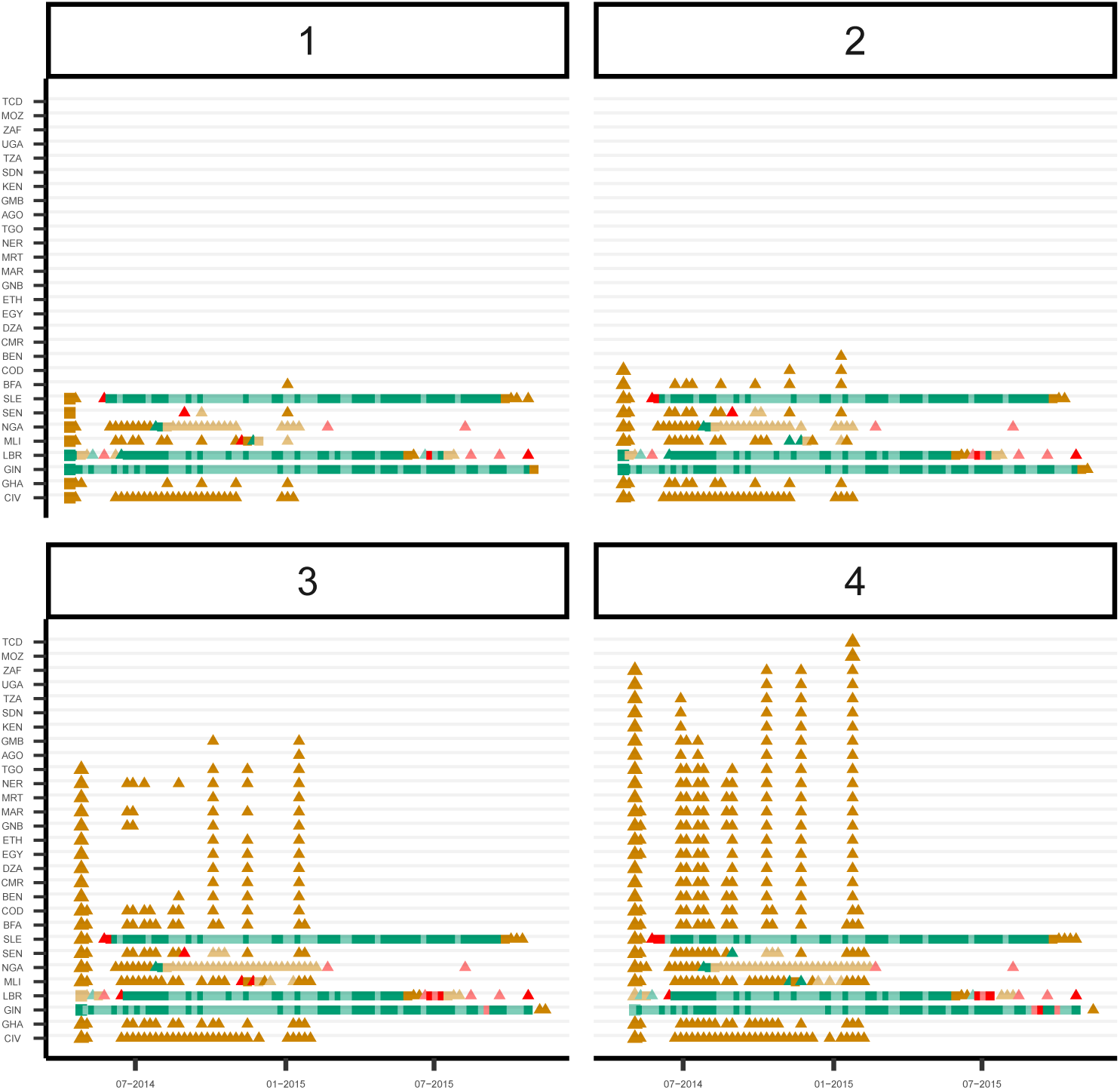
Predicted weekly presence of cases in each country up to 4 weeks ahead. The panels shows the True (green), False (orange) and Missed (red) 1, 2, 3 and 4 week ahead alerts using the 42.5^th^ percentile of the forecast interval as threshold. The figure only shows countries on the African continent for which either the 42.5^th^ percentile of the forecast interval or the observed incidence was greater than 0 at least once. The first alert in each country is shown using larger symbols (square or triangle). Alerts in a country in a week where there were no observed cases in the previous week are shown using hollow triangles. In each case, weeks for which all observed points were imputed are shown in lighter shades. Country codes, shown on the y-axis, are as follows: AGO - Angola, BEN - Benin, BFA - Burkina Faso, CIV – Côte d’Ivoire, CMR - Cameroon, COD - Democratic Republic of Congo, DZA - Algeria, EGY - Egypt, ETH - Ethiopia, GHA - Ghana, GIN - Guinea, GMB - Gambia, GNB - Guinea-Bissau, KEN - Kenya, LBR - Liberia, MAR - Morocco, MLI - Mali, MOZ - Mozambique, MRT - Mauritania, NER - Niger, NGA - Nigeria, SDN - Sudan, SEN - Senegal, SLE - Sierra Leone, SSD - South Sudan, TCD - Chad, TGO - Togo, TUN - Tunisia, TZA - Tanzania, UGA - Uganda, ZAF - South Africa. The alerts are based on forecasts using the ProMED data, a 2-week calibration window and a 4 week forecast horizon.

**Fig SI 48:**
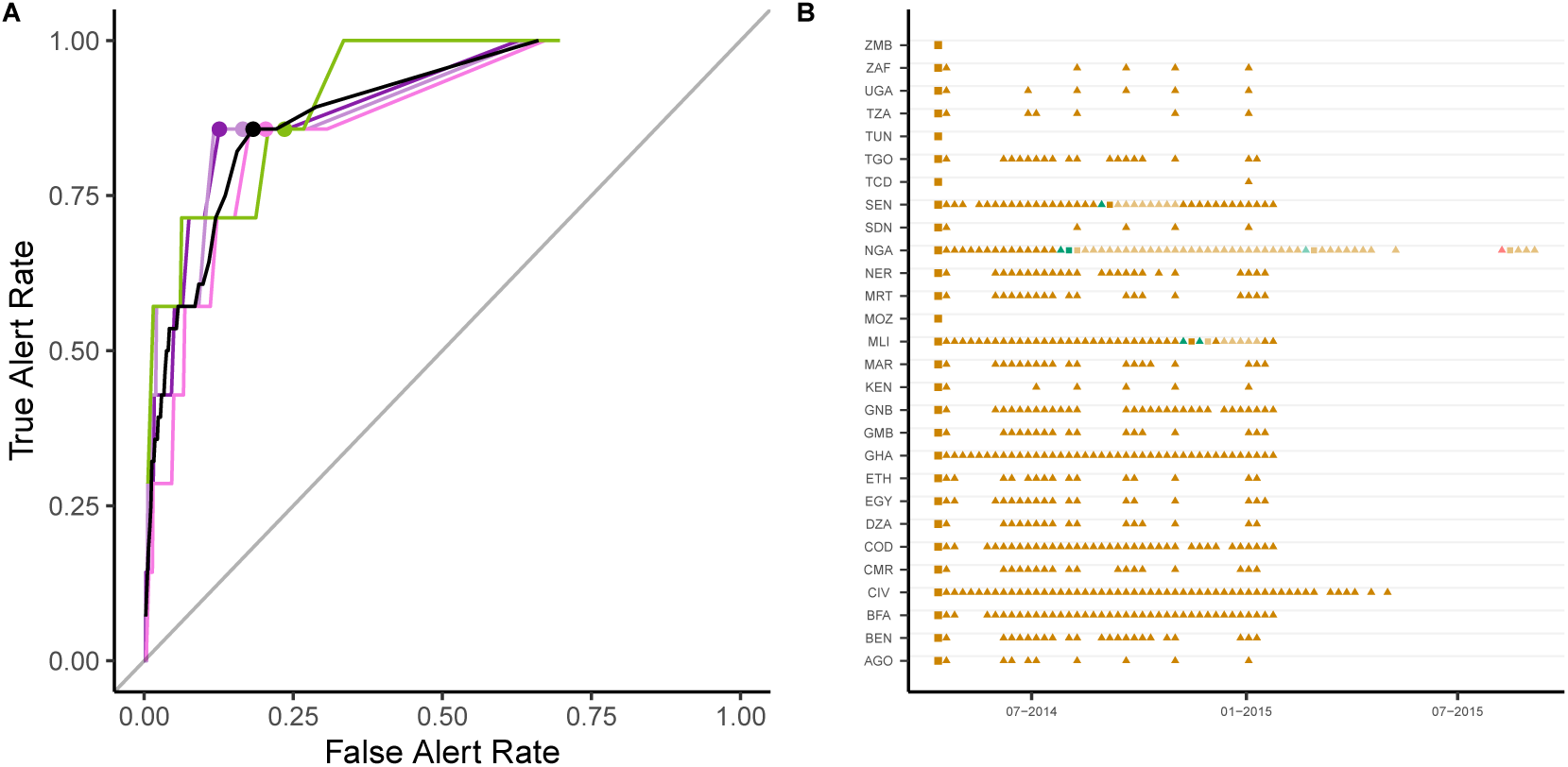
Predicted weekly presence of cases in countries other than the three majorly affected countries (Guinea, Liberia and Sierra Leone). The left panel shows the True and False alert rates using different thresholds for classification for alerts raised 1 (violet), 2 (light violet), 3 (dark pink) and 4 (light green) weeks ahead. The black curve depicts the overall True and False alert rates. On each curve, the dot shows the True and False Alert rates at 92.5% threshold. The right panel shows the True (green), False (orange) and Missed (red) 1 week ahead alerts using the 92.5^th^ percentile of the forecast interval as threshold. The figure only shows countries on the African continent for which either the 92.5^th^ percentile of the predicted incidence or the observed incidence was greater than 0 at least once. The first alert in each country is shown using larger symbols (square or triangle). Alerts in a country in a week where there were no observed cases in the previous week are shown using hollow triangles. In each case, weeks for which all observed points were imputed are shown in lighter shades. Country codes, shown on the y-axis, are as follows: AGO - Angola, BEN - Benin, BFA - Burkina Faso, CIV – Côte d’Ivoire, CMR - Cameroon, COD - Congo - Kinshasa, DZA - Algeria, EGY - Egypt, ETH - Ethiopia, GHA - Ghana, GMB - Gambia, GNB - Guinea-Bissau, KEN - Kenya, MAR - Morocco, MLI - Mali, MRT - Mauritania, NER - Niger, NGA - Nigeria, SDN - Sudan, SEN - Senegal, TGO - Togo, TZA - Tanzania, UGA - Uganda, ZAF - South Africa, MOZ - Mozambique, MWI - Malawi, SSD - South Sudan, TCD - Chad, TUN - Tunisia, BDI - Burundi, CAF - Central African Republic, COG - Congo - Brazzaville, LBY - Libya, MDG - Madagascar, RWA - Rwanda, ZMB - Zambia, ZWE - Zimbabwe, ERI - Eritrea, GAB - Gabon, SOM - Somalia, CPV - Cape Verde, GNQ - Equatorial Guinea, NAM - Namibia. The alerts are based on forecasts using the ProMED data, a 2-week calibration window and a 4 week forecast horizon.

**Fig SI 49:**
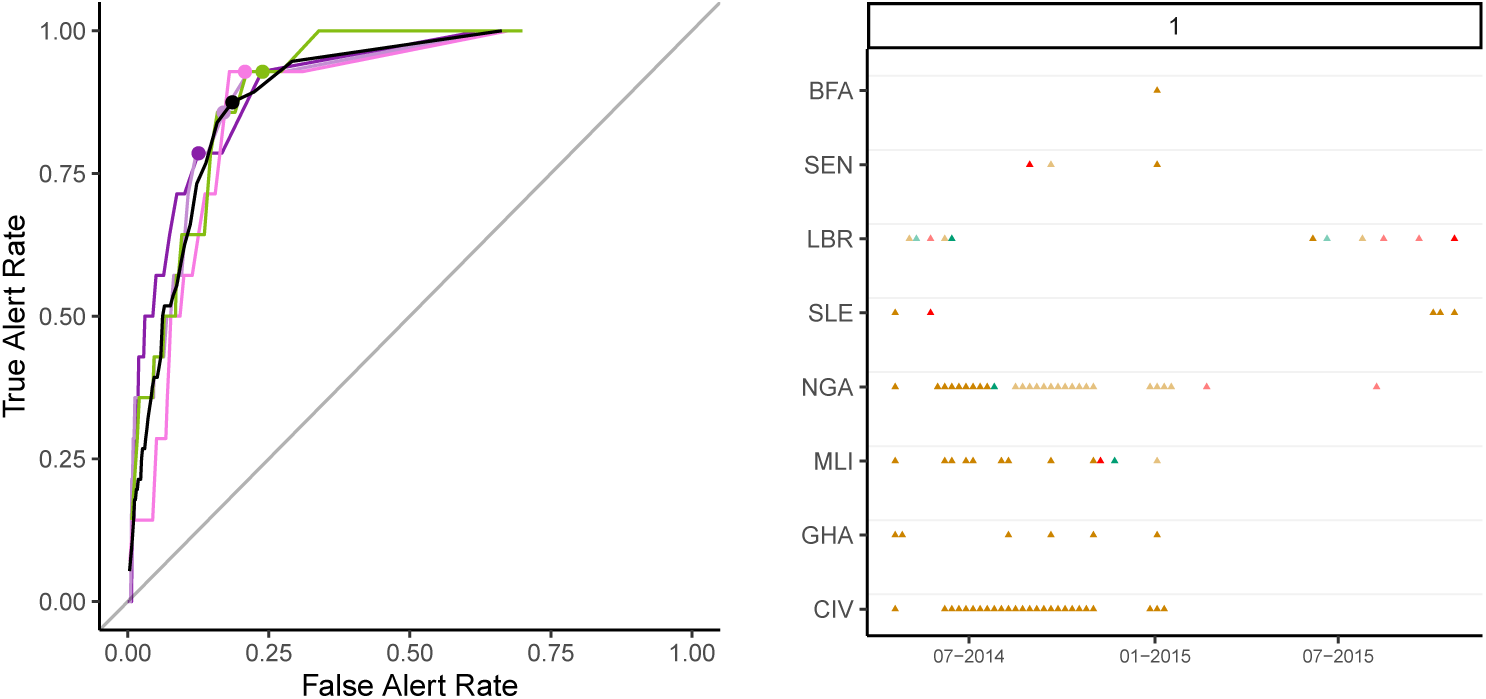
Predicted weekly presence of cases for each country in weeks following a week with no observed cases. The left panel shows the True and False alert rates using different thresholds for classification for alerts raised 1 (violet), 2 (light violet), 3 (dark pink) and 4 (light pink) weeks ahead. The black curve depicts the overall True and False alert rates. On each curve, the dot shows the True and False Alert rates at 93% threshold. For a given threshold (*x*^*th*^ percentile of the forecast interval), we defined a True alert for a week where the *x*^*th*^ percentile of the forecast interval and the observed incidence for a country were both greater than 0; false alert for a week where the threshold for a country was greater than 0 but the observed incidence for that country was 0; and missed alert for a week where the threshold for a country was 0 but the observed incidence for that country was greater than 0. True alert rate is the ratio of correctly classified true alerts to the total number of true and missed alerts (i.e., (true alerts)/(true alerts + missed alerts)). False alert rate is similarly the ratio of false alerts to the total number of false alerts and weeks of no alert (where the observed and the threshold incidence are both 0). The right panel shows the True (green), False (orange) and Missed (red) 1 week ahead alerts using the 93^rd^ percentile of the forecast interval as threshold. The figure only shows countries on the African continent for which either the 93^rd^ percentile of the predicted incidence or the observed incidence was greater than 0 at least once. The first alert in each country is shown using larger symbols (square or triangle). Alerts in a country in a week where there were no observed cases in the previous week are shown using hollow triangles. In each case, weeks for which all observed points were imputed are shown in lighter shades. Country codes, shown on the y-axis, are as follows: BFA - Burkina Faso, CIV - Côte d’Ivoire, GHA - Ghana, GIN - Guinea, LBR - Liberia, MLI - Mali, NGA - Nigeria, SEN - Senegal, SLE - Sierra Leone. The alerts are based on forecasts using the ProMED data, a 2-week calibration window and a 4 week forecast horizon.

**Fig SI 50:**
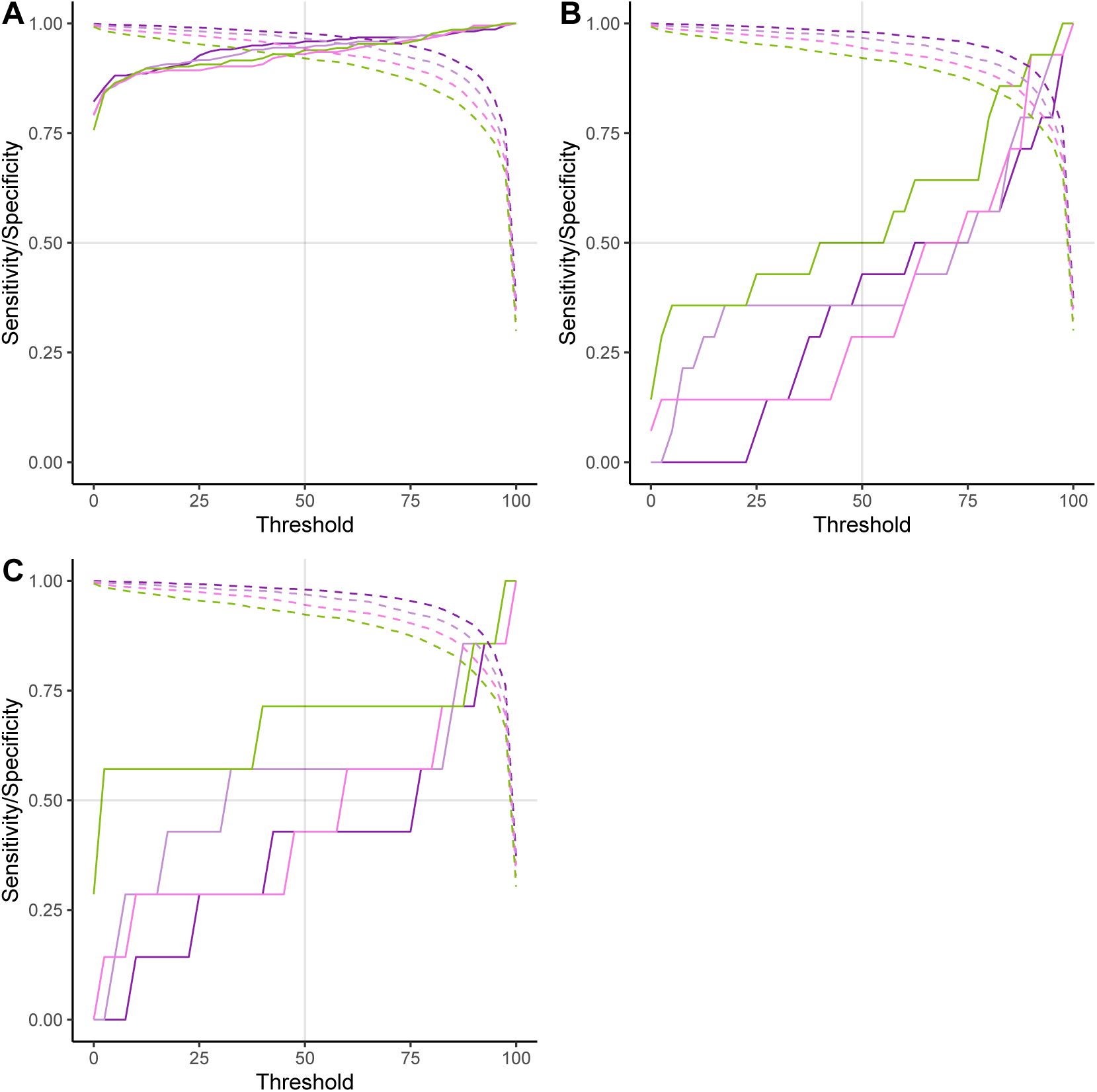
Sensitivity and specificity at various thresholds for (A) all countries in Africa (B) all countries in Africa except Guinea, Liberia and Sierra Leone, and (C) all in Africa in weeks following a week with no observed cases. The solid and dashed lines depict the sensitivity and specificity respectively of 1 (violet), 2 (light violet), 3 (pink) and 4 (light green) week ahead alerts.

**Fig SI 51:**
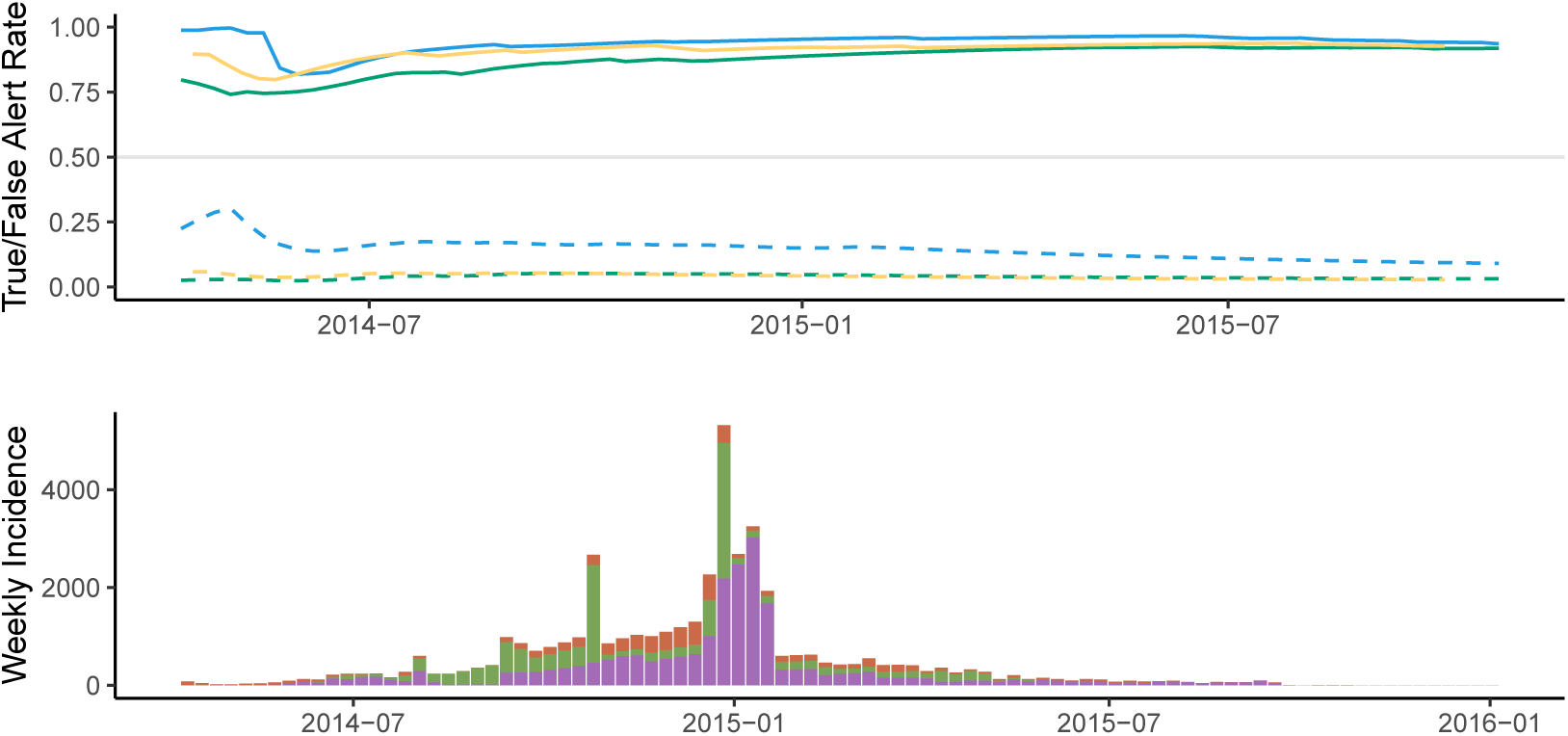
True and False alert rates at 50% threshold using ProMED (blue), HealthMap (green) and WHO (yellow) data over the course of the epidemic. The solid lines show the True Alert rate and the dashed lines show the False alert rate averaged over the 4 weeks forecast horizon. The bottom panel shows the weekly incidence for Guinea (deep orange), Liberia (green) and Sierra Leone (violet) obtained from ProMED data.

## References

[1] Stephen S Morse. Factors in the Emergence of Infectious Diseases.In Plagues and Politics, pages 8–26. Springer, 2001.

[2] Steven T Stoddard, Amy C Morrison, Gonzalo M Vazquez-Prokopec, Valerie Paz Soldan, Tadeusz J Kochel, Uriel Kitron, John P Elder, and Thomas W Scott. The role of human movement in the transmission of vector-borne pathogens. PLoS neglected tropical diseases, 3(7):e481, 2009.

[3] Resolution of the Executive Board of the WHO. Communicable diseases prevention and control: new, emerging, and re-emerging infectious diseases. 1995.

[4] World Health Organisation. Fjactors that contributed to undetected spread of the Ebola virus and impeded rapid containment. https://www.who.int/csr/disease/ebola/one-year-report/factors/en/, 2015. [Accessed 11-Mar-2019].

[5] Moritz UG Kraemer, Nuno R Faria, Robert C Reiner Jr, Nick Golding, Birgit Nikolay, Stephanie Stasse, Michael A Johansson, Henrik Salje, Ousmane Faye, GR William Wint, et al. Spread of yellow fever virus outbreak in angola and the democratic republic of the congo 2015–16: a modelling study. The Lancet infectious diseases, 17(3):330–338, 2017.

[6] Sean Wasserman, Paul Anantharajah Tambyah, and Poh Lian Lim. Yellow fever cases in Asia: primed for an epidemic. International Journal of Infectious Diseases, 48:98–103, 2016.

[7] A Tariq, K Roosa, K Mizumoto, and G Chowell. Assessing reporting delays and the effective reproduction number: The ebola epidemic in drc, may 2018–january 2019. Epidemics, 26:128–133, 2019.

[8] Individuals using the internet (% of population). https://data.worldbank.org/indicator/it.net.user.zs, 2019. [Online, accessed 08-Nov-2019].

[9] Thomas W Grein, KB Kamara, Guénaël Rodier, Aileen J Plant, Patrick Bovier, Michael J Ryan, Takaaki Ohyama, and David L Heymann. Rumors of disease in the global village: outbreak verification. Emerging Infectious Diseases, 6(2):97, 2000.

[10] Aranka Anema, Sheryl Kluberg, Kumanan Wilson, Robert S Hogg, Kamran Khan, Simon I Hay, Andrew J Tatem, and John S Brownstein. Digital surveillance for enhanced detection and response to outbreaks. The Lancet Infectious Diseases, 14(11):1035–1037, 2014.

[11] Stephen S Morse. Public Health Surveillance and Infectious Disease Detection. Biosecurity and Bioterrorism: Biodefense Strategy, Practice, and Science, 10(1):6–16, 2012.

[12] Clark C Freifeld, Kenneth D Mandl, Ben Y Reis, and John S Brownstein. HealthMap: Global Infectious Disease Monitoring through Automated Classification and Visualization of Internet Media Reports. Journal of the American Medical Informatics Association, 15(2):150–157, 2008.

[13] Zika virus Pacific (07): Chile (Easter Island), French Polynesia, 2014.

[14] Nicholas Generous, Geoffrey Fairchild, Alina Deshpande, Sara Y Del Valle, and Reid Priedhorsky. Global disease monitoring and forecasting with Wikipedia. PLoS computational biology, 10(11):e1003892, 2014.

[15] Gabriel J Milinovich, Ricardo J Soares Magalhães, and Wenbiao Hu. Role of big data in the early detection of Ebola and other emerging infectious diseases. The Lancet Global Health, 3(1):e20–e21, 2015.

[16] Gerardo Chowell, Julie M Cleaton, and Cecile Viboud. Elucidating transmission patterns from internet reports: Ebola and Middle East Respiratory Syndrome as case studies. The Journal of Infectious Diseases, 214(Suppl 4):S421–S426, 2016.

[17] WHO Ebola Response Team. Ebola Virus Disease in West Africa — The First 9 Months of the Epidemic and Forward Projections. New England Journal of Medicine, 371(16):1481–1495, 2014.

[18] WHO Ebola Response Team. West African Ebola Epidemic after One Year —- Slowing but Not Yet under Control. The New England Journal of Medicine, 372(6):584, 2015.

[19] Christophe Fraser. Estimating individual and household reproduction numbers in an emerging epidemic. PloS one, 2(8):e758, 2007.

[20] Anne Cori, Neil M Ferguson, Christophe Fraser, and Simon Cauchemez. A New Framework and Software to Estimate Time-Varying Reproduction Numbers During Epidemics. American Journal of Epidemiology, 178(9):1505–1512, 2013.

[21] George Kingsley Zipf. The P_1_ P2/D Hypothesis on the Intercity Movement of Persons. American Sociological Review, 11(6):677–686, 1946.

[22] How Liberia got to zero cases of Ebola. https://www.who.int/features/2015/liberia-ends-ebola/en/, 2015. accessed: 2019-11- 01.

[23] Ebola transmission in Liberia over. Nation enters 90-day intensive surveillance period. https://www.who.int/mediacentre/news/statements/2015/ebola-transmission-over-liberia/en/, 2015. accessed: 2019-11-01.

[24] RN Thompson, JE Stockwin, RD van Gaalen, JA Polonsky, ZN Kamvar, PA Demarsh, E Dahlqwist, S Li, E Miguel, T Jombart, et al. Improved inference of time-varying reproduction numbers during infectious disease outbreaks. Epidemics, page 100356, 2019.

[25] Rosalind M Eggo, Conall H Watson, Anton Camacho, Adam J Kucharski, Sebastian Funk, and W John Edmunds. Duration of ebola virus rna persistence in semen of survivors: population-level estimates and projections. Eurosurveillance, 20(48):30083, 2015.

[26] Stephen K Gire, Augustine Goba, Kristian G Andersen, Rachel SG Sealfon, Daniel J Park, Lansana Kanneh, Simbirie Jalloh, Mambu Momoh, Mohamed Fullah, Gytis Dudas, et al. Genomic surveillance elucidates Ebola virus origin and transmission during the 2014 outbreak. science, 345(6202):1369–1372, 2014.

[27] World Health Organisation. WHO: Ebola Response Roadmap Situation Report 1. https://www.who.int/iris/bitstream/10665/131974/1/roadmapsitrep1_eng.pdf, 2014. [Online; accessed 20-Feb-2019].

[28] ProMED. UNDIAGNOSED VIRAL HEMORRHAGIC FEVER - GUINEA (02): EBOLA CONFIRMED. https://www.promedmail. org/post/2349696, 2014. [Online; accessed 20-Feb-2019].

[29] Max SY Lau, Benjamin Douglas Dalziel, Sebastian Funk, Amanda McClelland, Amanda Tiffany, Steven Riley, C Jessica E Metcalf, and Bryan T Grenfell. Spatial and temporal dynamics of superspreading events in the 2014–2015 West Africa Ebola epidemic. Proceedings of the National Academy of Sciences, 114(9):2337–2342, 2017.

[30] Junerlyn Agua-Agum, Archchun Ariyarajah, Bruce Aylward, Luke Bawo, Pepe Bilivogui, Isobel M Blake, Richard J Brennan, Amy Cawthorne, Eilish Cleary, Peter Clement, et al. Exposure patterns driving Ebola transmission in West Africa: a retrospective observational study. PLoS Medicine, 13(11):e1002170, 2016.

[31] Samuel T Boland, Erin Polich, Allison Connolly, Adam Hoar, Tom Sesay, and Anh-Minh A Tran. Overcoming operational challenges to Ebola case investigation in Sierra Leone. Global Health: Science and Practice, 5(3):456–467, 2017.

[32] Mapping the Risk of International Infectious Disease Spread (MRI- IDS). https://github.com/healthsites/mRIIDS/wiki, 2019. Accessed: 2019-08-19.

[33] Filippo Simini, Marta C González, Amos Maritan, and Albert-László Barabási. A universal model for mobility and migration patterns. Na- ture, 484(7392):96, 2012.

[34] healthsites.io. https://healthsites.io, 2019. accessed: 2019-08-09.

[35] Joseph Maina, Paul O Ouma, Peter M Macharia, Victor A Alegana, Benard Mitto, Ibrahima Socé Fall, Abdisalan M Noor, Robert W Snow, and Emelda A Okiro. A spatial database of health facilities managed by the public health sector in sub Saharan Africa. Scientific data, 6(1):134, 2019.

[36] John G Saw, Mark CK Yang, and Tse Chin Mo. Chebyshev Inequality with Estimated Mean and Variance. The American Statistician, 38(2):130–132, 1984.

[37] Tini Garske, Anne Cori, Archchun Ariyarajah, Isobel M. Blake, Ilaria Dorigatti, Tim Eckmanns, Christophe Fraser, Wes Hinsley, Thibaut Jombart, Harriet L. Mills, Gemma Nedjati-Gilani, Emily Newton, Pierre Nouvellet, Devin Perkins, Steven Riley, Dirk Schumacher, Anita Shah, Maria D. Van Kerkhove, Christopher Dye, Neil M. Ferguson, and Christl A. Donnelly. Heterogeneities in the case fatality ratio in the West African Ebola outbreak 2013–2016. Philosophical Transactions of the Royal Society of London B: Biological Sciences, 372(1721), 2017.

[38] LandScan1 Global Population Database. http://www.ornl.gov/ landscan, 2017.

[39] Tobias Grosche, Franz Rothlauf, and Armin Heinzl. Gravity models for airline passenger volume estimation. Journal of Air Transport Management, 13(4):175 – 183, 2007.

[40] Maria D Van Kerkhove, Ana I Bento, Harriet L Mills, Neil M Ferguson, and Christl A Donnelly. A Review of Epidemiological Parameters from Ebola Outbreaks to Inform Early Public Health Decision-Making. Scientific Data, 2:150019, 2015.

[41] Bob Carpenter, Andrew Gelman, Matthew D Hoffman, Daniel Lee, Ben Goodrich, Michael Betancourt, Marcus Brubaker, Jiqiang Guo, Peter Li, and Allen Riddell. Stan: A probabilistic programming language. Journal of Statistical Software, 76(1), 2017.

[42] Stan Development Team. RStan: the R interface to Stan, 2018. R package version 2.18.2.

[43] Andrew Gelman, Donald B Rubin, et al. Inference from iterative simulation using multiple sequences. Statistical science, 7(4):457–472, 1992.

[44] John F. Geweke. Evaluating the accuracy of sampling-based approaches to the calculation of posterior moments. In A.P. Dawid J.O. Berger, J.M. Bernardo and A.F.M. Smith., editors, Bayesian Statistics 4. Clarendon Press, Oxford, 1992.

[45] Xavier Fernández i Marín. ggmcmc: Analysis of MCMC samples and Bayesian inference. Journal of Statistical Software, 70(9):1–20, 2016.

[46] Chris Tofallis. A better measure of relative prediction accuracy for model selection and model estimation. Journal of the Operational Research Society, 66(8):1352–1362, 2015.

[47] Sebastian Funk, Anton Camacho, Adam J Kucharski, Rachel Lowe, Rosalind M Eggo, and W John Edmunds. Assessing the performance of real-time epidemic forecasts. bioRxiv, page 177451, 2017.

